# DEVELOPMENT OF ORGANOID BASED MODEL TO STUDY IMMUNE-NEURAL INTERACTIONS IN HUMAN CNS DISEASES

**DOI:** 10.64898/2026.08.14.26360461

**Authors:** Joanna Kocot, Sahar H. Pradhan, Dragan Maric, Peter Kosa, Clayton Winkler, Cihan Oguz, Timothy G. Myers, Gustaf Wigerblad, Justin Lack, Cathryn Haigh, Karin Peterson, Bibiana Bielekova

**Author notes:** Correspondence (B.B.).

## Abstract

Modeling neural-immune interactions in neurodegenerative and immune-mediated central nervous system (CNS) diseases requires human 3D models that capture cellular diversity and long-term tissue maturation. Here, we present an enhanced human induced pluripotent stem cell (hiPSC)-derived cerebral organoid (CO) platform optimized to mitigate core hypoxia for over 200 days. Timed pro-myelinating cues established organized neuronal layering and progressive axonal myelination through day 140, while vascular fusion yielded assembloids incorporating endothelial structures and microglia. Extended culture (>500–750 days) spontaneously reproduced hallmark features of human CNS aging, including cellular senescence signatures, neuroaxonal loss, hypomyelination, and the autonomous emergence of a neurotoxic astrocyte transcriptional profile in the complete absence of microglia or immune cells. Co-culture with autologous activated peripheral blood mononuclear cells (PBMC) resulted in transient immune infiltration and a pronounced type II interferon response across CNS lineages. High-plex spatial transcriptomics revealed that immune cell infiltration was associated with oligodendrocyte loss and in aged organoids also with downregulated oligodendrocyte myelin gene transcription. While not fully reproducing adult tissue stoichiometry, this platform enables longitudinal modeling of neural-immune crosstalk in age-related and neuroinflammatory CNS disorders.

## INTRODUCTION

The development of effective treatments for chronic polygenic diseases affecting the central nervous system (CNS) has been exceptionally slow. A major obstacle is the lack of disease models that accurately replicate the complex and evolving biology of the disease interacting with and modifying the human CNS.

Human induced pluripotent stem cell (hiPSC)-derived cerebral organoids (COs) have revolutionized the study of neuronal biology by providing three-dimensional *in vitro* cultures that successfully recapitulate neuronal differentiation during embryonic and post-natal development. However, such COs failed to fully reflect the cellular composition and stoichiometry of the human CNS. While novel protocols have reported the formation of choroid plexus-like structures^1^, current COs generally lack oligodendrocytes (and thus myelinated axons), a stable microglial population, and most mesenchymal cells, including epithelial cells (i.e., ependymal and choroid plexus epithelium) and vasculature-associated cells (vascular endothelial cells [VEC], lymphatic endothelial cells [LEC], pericytes, fibroblasts and smooth muscle cells [SMC]).

Individual studies tried to address separately generation of each missing CNS cell type:

1. <u>Oligodendrocytes and myelination:</u> Human pre-myelinating oligodendrocytes differentiate from oligodendrocyte precursor cells (OPCs) in the late gestational period (i.e., gestational weeks (qw) 30-40). Most myelination occurs postnatally, likely driven by increasing activity of neuronal circuits, such motoric/sensory integration and emergence of cognition. As current CO protocols fail to reproduce this emergent property, novel guided protocols induced oligodendrocyte differentiation by adding pro-myelinating signals.^2–6^ The development of myelinating oligodendrocytes was achieved through either the timed administration of growth factor and hormone cocktails (including e.g., FGF, PDGF, NT3, IGF-1, HGF, and T3)^2–5^ or the overexpression of transcription factors SOX10 and OLIG2^6^, which promote oligodendrocyte differentiation and maturation. While successfully generating oligodendrocytes, these protocols are nowhere close to approximating post-developmental white matter where oligodendrocyte density represents 75-78% of cells^7^ and all large diameter axons are fully myelinated.
2. <u>Microglia:</u> Microglia are CNS innate immune cells that originate from erythromyeloid progenitors in the embryonic yolk sac and colonize the human brain between gw 4-24.^8^ Microglia comprise 5–15% of the total brain cells with variable densities and transcriptional patterns across CNS regions^9^. Microglia-containing COs were generated either by inducing the spontaneous formation of microglia^10,11^ or by co-culturing the COs with microglial progenitors or mature microglia.^9,12,13^ However, current protocols do not produce COs with a stable population of microglial cells. In protocols that depend on spontaneous differentiation, the proportion and distribution of microglia within the organoids tend to be highly inconsistent due to the random and unpredictable nature of unguided differentiation.^10,11^ When microglia are introduced through co-culture, their integration into the COs is often incomplete, leading to uneven distribution and suboptimal interactions with neurons.^14^ *In vivo*, microglia interact with astrocytes and oligodendrocytes, which significantly influence their activation state and phagocytic function. However, the existing protocols for microglia-containing COs either do not produce any or only minimal populations of these essential glial cell types. This limitation restricts the critical cellular interactions necessary for proper microglial development and function.
3. <u>CNS epithelial cells:</u> In the CNS, epithelial cells play important roles in the formation of the blood brain (BBB) and blood-cerebrospinal fluid (CSF) barriers, in CSF dynamics, and immune regulation.^15,16^ Choroid plexus epithelial cells (ChPlEp) form tight junctions that create the blood-CSF barrier. They are also responsible for secreting CSF.^16^ Ependymal cells line the brain’s ventricles and the central canal of the spinal cord. They are essential for CSF homeostasis and dynamics. Unlike ChPlEp, ependymal cells do not have tight junctions, resulting in a diffusive barrier between CSF and the brain’s interstitial fluid.^15^ The COs generated by standard protocols do not contain the choroid plexus. On the other hand, choroid plexus organoids effectively model both the epithelial and stromal components of the choroid plexus and produce CSF-like fluid; however, they lack the mature neurons.^17^ COs cultured using the air-liquid interface (ALICO) method can sometimes develop ChPlEp structures.^1^
4. <u>Vasculature-associated mesenchymal cells:</u> Vasculature-associated mesenchymal cells, particularly pericytes and SMC, play a crucial role in the formation and maintenance of the BBB, facilitating vascular repair, and regulating immune responses after CNS injuries. Pericytes regulate BBB permeability, angiogenesis, cerebral blood flow, neuroinflammation, and stem cell activity. Their dysfunction leads to BBB breakdown, allowing the infiltration of immune cells and neurotoxins. This can result in excessive inflammation and secondary neuronal damage.^18^ SMC control cerebrovascular dynamics. After an injury, SMC dysfunction can cause vascular constriction, ischemia, and hypoxia, which exacerbate the effects of CNS injuries. Growing evidence suggests that the pathophysiology of SMC plays a critical role in the complex processes involved in neurodegeneration.^19^ To tackle the challenge of vascularization, COs were co-cultured with vascular or mesodermal spheroids. When fused with COs, the resulting organoids/assembloids exhibited neuronal progenitors, neurons, VEC and microglia. However, they lacked astrocytes and oligodendrocytes, which typically emerge at later developmental stages.^20^

We found no published studies that have attempted to integrate the aforementioned strategies to generate COs that approximate the cellular composition of the postnatal human CNS. This gap limits the utility of COs for modeling chronic polygenic CNS diseases, which typically induce functional changes across all CNS cell types, particularly as the disease progresses to chronicity. Thus, addressing these shortcomings is critical for developing models that accurately reflect disease pathology and facilitate the discovery of effective treatments.

Furthermore, CNS diseases do not progress in isolation from the rest of the body. Immune cells are actively recruited to the injured CNS, and their quantity, phenotype, and activation status play a pivotal role in determining whether CNS undergoes recovery or further destruction. The interaction between immune cells and CNS mesenchymal cells also governs the compartmentalization of immune responses within the CNS, determining the chronicity of intrathecal inflammation. Apart from recent papers describing glioma-immune interactions^21^, we found no human 3-dimentional (3D) multicellular culture models studying the crosstalk between CNS cells and autologous (blood-derived) immune cells under physiological or disease conditions. Developing such models is essential for understanding the immunopathogenic mechanisms underlying chronic CNS diseases and for translating this knowledge into therapeutic strategies that modulate immune responses to promote healing rather than injury.

The objective of this study was to evaluate the integration (and optimization) of the aforementioned strategies to develop 3D COs that approximate the cellular composition of the postnatal human CNS. We also aimed to assess the spontaneous evolution of these complex COs over extended culture periods to determine whether COs could be adopted to study age-related neurodegeneration and to explore the feasibility of studying autologous neural-immune interactions. Establishing such models is a prerequisite for developing next-generation models of diverse chronic CNS diseases.

## RESULTS

### Induction of oligodendrocyte precursor cells differentiation between days 50 and 70 leads to progressively greater myelination as organoids mature between days 98 and 140

We generated the COs following the protocol by Lancaster et al.^22^, with some modifications. The long-term culture of COs often leads to necrosis of the organoid core. ^1,22^ As development of necrotic core would release danger-associated molecular patterns (DAMPs) which are immunogenic and thus may affect neural-immune interactions, we asked whether decreasing the number of hiPSCs used to create the embryoid body from 9,000 to 5,000 and 2,500 would decrease necrotic core without affecting neuronal development. Indeed, these smaller organoids did not develop visible necrotic core up to day 540 (Figure S1 and S10). Although small cavity-like structures were occasionally observed in multicolor fluorescence microscopy sections at day 98 (Figure S2), the surrounding nuclei showed no signs of pyknosis (Figure S2A). Furthermore, these regions were bordered by neurons displaying normal morphology and preserved marker expression. Specifically, microtubule-associated protein 2 (MAP2) and class III β-tubulin (TUBB3) outlined intact cell bodies, neurofilament light (NF-L) staining was sparse and punctate (Figure S2C,D). Astrocytes with SOX2+ nuclei were sparse and primarily proliferative (Ki67+) (compare Figure S2E and S2F); their intracytoplasmic expression of glial fibrillary acidic protein (GFAP) was modest and comparable to vimentin (VIM) levels (Figure S2E). Finally, we observed weak, punctate cytoplasmic HIF-1α staining (Figure S2B), consistent with normoxic tissue. Under normoxia, HIF-1α is continuously translated and targeted for degradation within endosomal/autophagosomal compartments, maintaining a minimal baseline pool. This contrasts with pathological hypoxia, which triggers HIF-1α stabilization and nuclear translocation to drive an adaptive transcriptional response. Consequently, our protocol modifications successfully mitigate inner core hypoxia and visible necrotic core formation (latter up to d540), indicating that the minor cavities observed on day 98 represent processing artifacts.

We found that decreasing the initial number of hiPSC did not alter neurogenesis, as d40 COs contained rosettes of proliferating Ki67^+^SOX2^+^ neuroglial precursors (see Figure S3B for Ki67 and Figure S3F for SOX2). The COs also contained three distinct populations of NEUN^+^ post-mitotic neurons (Figure S3A): TBR1^+^ deep layer projection neurons (in adult CNS located mostly in cortical layer 6, with corticothalamic projections) (Figure S3C); CTIP2^+^ cortical projection neurons that include corticospinal motor neurons (layer 5, projecting to spinal cord) (Figure S3D); and SATB2^+^ callosal projection neurons (layers 2/3, 5, projecting to contralateral cortex via corpus callosum) (Figure S3C).

TBR1⁺ and CTIP2⁺ neurons were found to co-localize in d40 organoids (compare Figure S3C for TBR1 and Figure S3D for CTIP2), while SATB2⁺ neurons tended to cluster separately (Figure S3C). We also observed development of OLIG2^+^ OPCs (Figure S3D), but the population was very rare. Neuronal layers showed diffuse MAP2 staining, more focal TUBB3 staining in deep layers, and very weak NFL staining in superficial layers (Figure S3E).

To develop myelinated COs, we adopted protocol that induces oligodendrocyte maturation and myelination in human cortical spheroids^3^ and performed these myelination-inducing steps between d50-70 as described in Methods.

At d98, we saw development of clearly differentiated neuronal layers with the least prevalent TBR1-positive neurons at the organoid edge, SATB2-positive neurons underneath and CTIP2-positive corticospinal motor neurons representing the deepest layer of post-mitotic NEUN-positive neurons (Figure 1A). MAP2 (Figure 1B, and tubulin, not shown) were diffusely staining neuropil around neuronal DAPI-positive nuclei. NFL stain was much more prominent than at d40 and strongly enriched at outer organoid edge (Figure 1C). The directionality of these NFL-stained axons was parallel to the organoid edge. In contrast to d40, we also observed clusters of NFL-positive axons deeper in organoid core (arrow in Figure 1C). These deeper NFL^+^ axons had larger diameter than superficial axons and had perpendicular directionality to organoid edge. Compared to d40 organoids the OLIG2^+^ glial cells were more expanded, distributed in all neuronal layers but also in the deeper parts of COs, however, their population remained sparse. Many OLIG2^+^ cells were proliferating based on positive Ki67^+^ staining (Figure 1D, magnified insert). We also observed myelin proteins, MBP (and PLP, not shown) completely overlapping with NFL (Figure 1E), indicating myelination of axons, including afore-mentioned large deep axons projecting perpendicularly to organoid edge (arrow in Figure 1E). Finally, we saw GFAP staining of astrocytic projections (Figure 1F) overlapping with layers of neuronal bodies but also extending through axonal layer to the organoid surface. In contrast, we saw no AQP1-positive astrocytes (data not shown).

**Figure 1.**
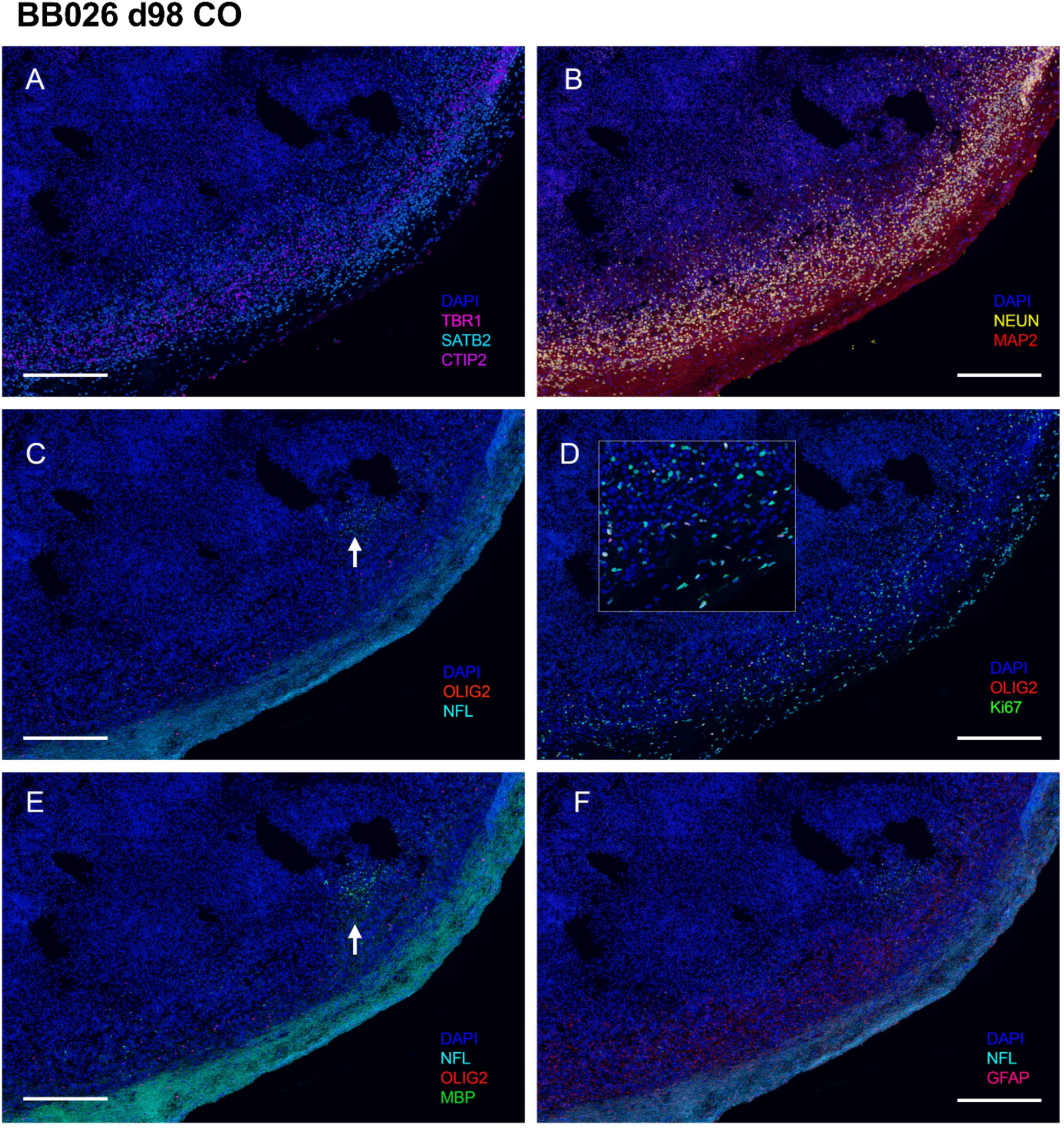
Distinct organization of mature neural layers comprising fully differentiated neurons, corticospinal motor neurons, astrocytes, and early myelinating axons in cerebral organoid at d98. A. Immunostaining of TBR1⁺ (pink), SATB2⁺ (blue), and CTIP2⁺ (purple) neurons in cryosection from 98-day-old organoid. Nuclei are stained with DAPI. *Scale bar, 300 µm*. B. Immunostaining of NEUN⁺ (yellow) post-mitotic neurons and MAP2⁺ (red) neuronal dendrites in cryosection from 98-day-old organoid. Nuclei are stained with DAPI. *Scale bar, 300 µm*. C. Immunostaining of NFL⁺ (cyan) neuronal axons and OLIG2⁺ (red) cells in cryosection from 98-day-old organoid. Nuclei are stained with DAPI. *Scale bar, 300 µm*. D. Immunostaining of OLIG2⁺ (red) cells and Ki67⁺ (green) proliferating cells in cryosection from 98-day-old organoid. Nuclei are stained with DAPI. *Scale bar, 300 µm*. E. Immunostaining of MBP⁺ (myelin basic protein; green) cells co-localizing with NFL⁺ (cyan) axons in cryosection from 98-day-old organoid. Nuclei are stained with DAPI. *Scale bar, 300 µm*. F. Immunostaining of GFAP⁺ (red) astrocytes and NFL⁺ (cyan) axons in cryosection from 98-day-old organoid. Nuclei are stained with DAPI. *Scale bar, 300 µm*. Representative immunofluorescence image of one 98-day-old cerebral organoid (donor BB026).

At d140, the neuronal layers changed: TBR1-positive neurons were mostly at the organoid center and aggregated in clusters; CTIP2-positive neurons were least prevalent, were located occasionally at organoid center and in clusters at organoid edge but stained faintly, while SATB2-positive neurons were majority of NEUN-positive post-mitotic neurons and clustered between superficial CTIP2-positive and deep TBR1-positive neurons and out towards NFL-positively stained axons (Figure 2A). Tubulin staining was widespread throughout the COs, while MAP2 expression increased progressively toward the outer regions (Figure 2B). NFL distribution at d140 resembled that of d98 organoids - organized parallel to the organoid edge, forming a barrier-like structure while also appearing in clusters of large-diameter axons in deeper regions. However, the NFL-positive layer was markedly thicker at d140 (Figure 2A,D; Figure S4A,B). OLIG2⁺ cells, representing both oligodendrocyte precursor cells (OPCs) and oligodendrocytes, were more abundant at d140 and evenly distributed throughout the organoid (Figure 2C; Figure S4C,D). Similarly, myelin proteins, both MBP and PLP, were detected across the full depth of the organoid, with the most intense staining in the outer layer, corresponding to the upper third (Figure 2C,D). This region also showed co-localization with NFL-positive axons (Figure S4C–E; donor BB028). We observed an analogous distribution of myelin proteins in a second donor, where a high-magnification view of the organoid edge revealed elongated OLIG2+ oligodendroglial nuclei arranged in linear ‘trains’ running parallel to NF-L+ axons, resembling interfascicular oligodendrocytes in human white matter tracts (Figure S5A-D; donor BB026).

**Figure 2.**
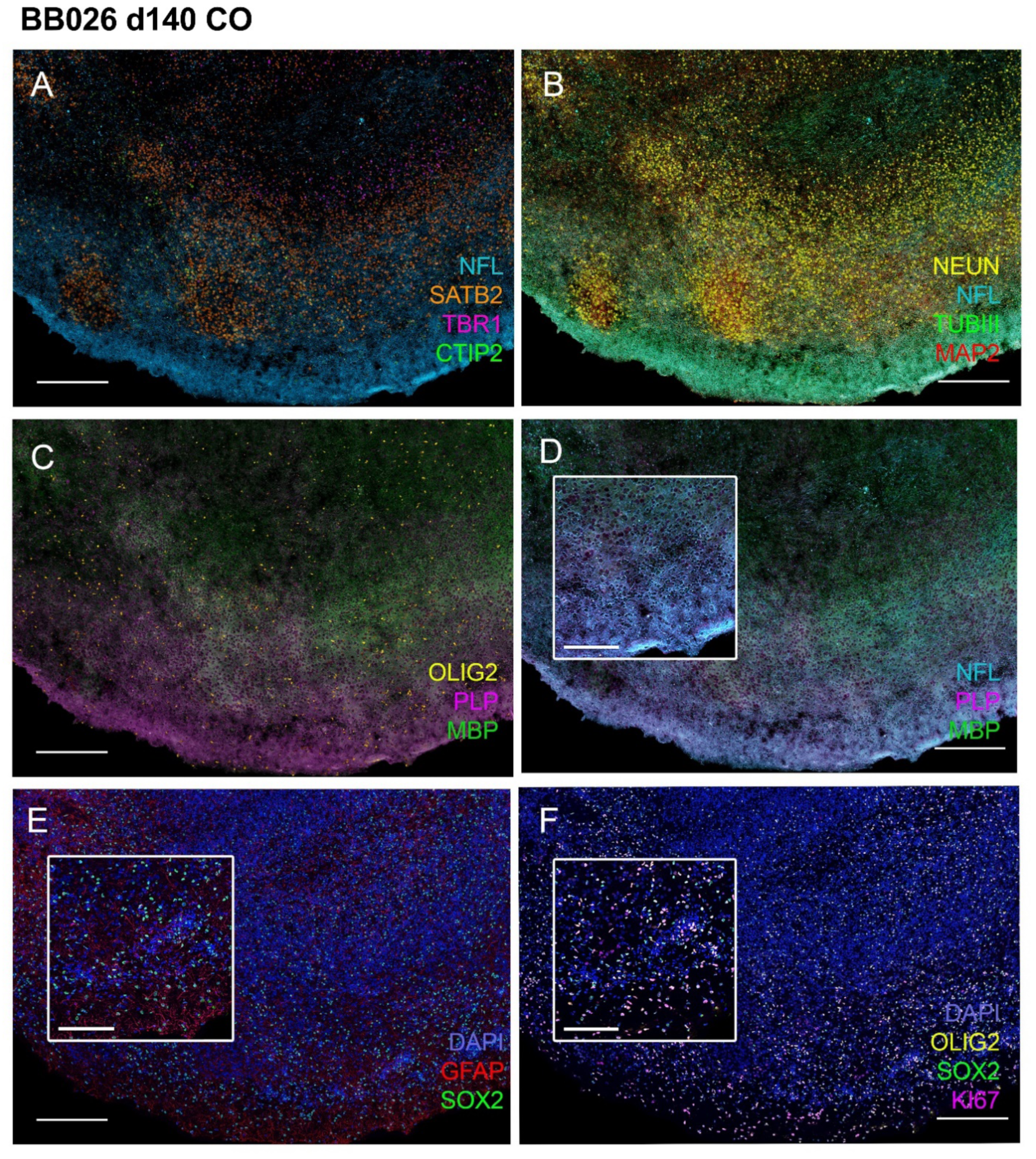
Distinct organization of mature neural layers comprising fully differentiated neurons, astrocytes, oligodendrocytes, and myelinated axons in cerebral organoid at d140. A. Immunostaining of NFL⁺ (cyan), SATB2^+^ (orange), TBR1^+^ (magenta), and CTIP2+ (green) cells in cryosection from 140-day-old organoid. *Scale bar, 300 µm*. B. Immunostaining of NEUN^+^ (yellow), NFL⁺ (cyan), MAP2⁺ (red), and TUBB3⁺ (green) cells in cryosection from 140-day-old organoid. *Scale bar, 300 µm*. C. Immunostaining of OLIG2⁺ (yellow), PLP^+^ (magenta) and MBP^+^ (green) cells in cryosection from 140-day-old organoid. *Scale bar, 300 µm*. D. Immunostaining of NFL⁺ axons (cyan) co-localized with PLP^+^ (magenta) and MBP+ (green) cells in cryosection from 140-day-old organoid. *Scale bar, 300 µm.* Magnified inset shows NFL⁺ axons co-localized with PLP⁺ regions. *Inset scale bar, 50 µm*. E. Immunostaining of GFAP⁺ (red) and SOX2⁺ (green) cells in cryosection from 140-day-old organoid. Nuclei are stained with DAPI (blue). *Scale bar, 300 µm. Inset scale bar, 50 µm*. F. Immunostaining of OLIG2⁺ (yellow) and SOX2⁺ (green), and Ki67⁺ (magenta) cells. Nuclei are stained with DAPI (blue). *Scale bar, 300 µm.* Inset highlights co-localization of OLIG2^+^, SOX2^+^, and Ki67^+^ cells. *Inset scale bar, 50 µm*. Representative immunofluorescence image of two 140-day-old cerebral organoids (donor BB026 & BB028).

SOX2⁺ cells overlapped with GFAP staining, which was diffusely present throughout the organoid but most prominent near the outer edge (Figure 2E; Figure S4E for SOX2 and S4F for GFAP). Ki67⁺ staining indicated that SOX2⁺ cells, along with a subset of OLIG2⁺ cells, were actively proliferating at d140 (Figure 2F).

Immunofluorescence quantification confirmed the structural maturation of cerebral organoids by day 140 (Figure S6). Compared with day 98, the MAP2-positive area increased, consistent with progressive neuronal maturation, while NFL-positive axonal area approximately doubled. Myelination markers exhibited the most pronounced changes, with MBP- and PLP-positive areas increasing markedly and reaching their highest levels at day 140, indicating oligodendrocyte maturation and myelin formation. In contrast, the Olig2-positive area increased only modestly, suggesting that the expansion of mature myelin-forming oligodendrocytes exceeded the increase in the overall oligodendrocyte lineage population. GFAP-positive astrocytic area also increased relative to day 98, reflecting continued astrocyte maturation during long-term culture.

Overall, adding myelination-inducing steps resulted in COs that continued to myelinate from d70 till at least d140 and had diverse population of post-mitotic neurons that were predominant cell type. Oligodendrocytes were prominent and astrocytes were mostly located in deeper organoid core without excessive GFAP abundance.

### Human CO microenvironment does not support homing, maturation and maintenance of microglia

To introduce microglia into the COs, we adopted the MIGRATE protocol developed by Fattorelli et al.^23^ We generated CX3CR1^+^/CD14^+^ microglial progenitors (MGPs)^24^, and co-cultured them (2×10^5^ cells per organoid) with the d30 COs to mimic the *in vivo* migration of primitive macrophages from the yolk sac into developing CNS tissue. Live imaging showed that, although MGPs attached to COs and interacted with CNS cells, their incorporation was minimal and uneven (Figure S7A-D). Furthermore, no TMEM119^+^ or Iba1^+^ microglial cells were observed in COs at 3 or 6 months after co-culture, as determined by immunohistochemistry (data not shown).

Thus, the current protocols for CO development not only did not yield mesenchymal cells, but also lacked the essential microenvironment to support homing, maturation and stability of exogenously generated autologous microglial precursors.

### Assembloids of COs with vascular organoids provide focal microenvironment that supports microglial development and incorporation into CNS tissue

Blood vessels are a well-known conduit for microglial colonization into the CNS, and it is possible that associated mesenchymal cells provide necessary microenvironment for microglial development, maturation and long-term CNS integration. To test this hypothesis we generated assembloids of early (d7) COs fused with autologous hiPSC-derived vascular organoids (Figure 3A) using a modified protocol from Sun et al.^20^ Within 18 days, distinct blood vessel-like structures lined with PECAM/CD31^+^ VEC or LEC had successfully infiltrated into the COs (Figure 3B-C). By d45, the fusion resulted in an engraftment of CD31^+^ vessel-like structures and Iba1^+^ microglial cells into the DCX^+^ neuronal tissue (Figure 3 D-F, Figure S8A,B). However, the desired full integration of neuroectodermal and mesenchymal tissues represented only the focal and limited part of the assembloid.

**Figure 3.**
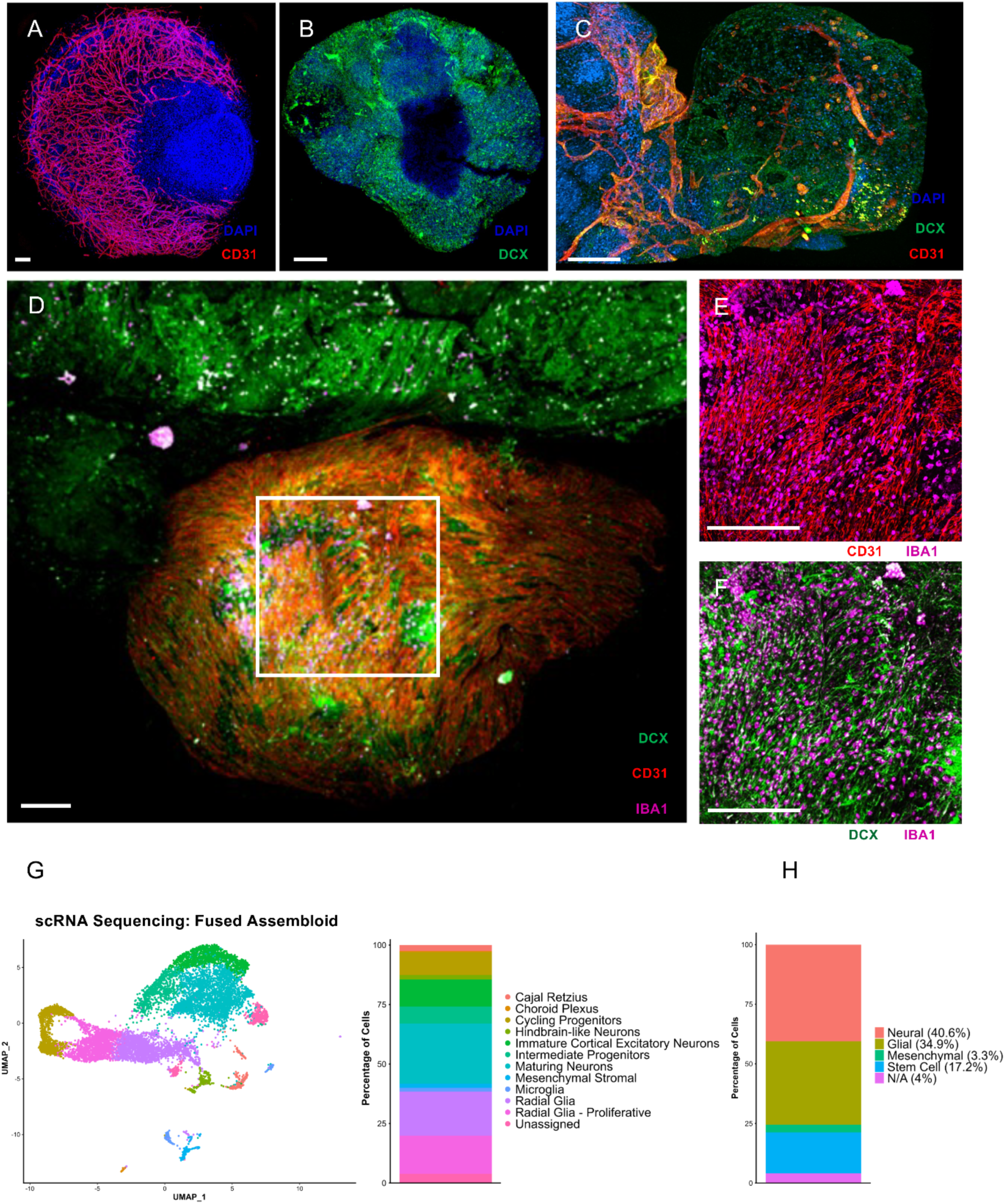
Fusion of vessel and cerebral organoids to generate vascularized, microglia-containing cerebral assembloids. A. Immunostaining of CD31⁺ (red) endothelial cells in the whole-mount vessel organoid at day 45. Nuclei are stained with DAPI (blue). *Scale bar, 200 µm*. B. Immunostaining of DCX⁺ (green) immature neurons in the whole-mount early-stage cerebral organoid at day 25. Nuclei are stained with DAPI (blue). Scale bar, *200 µm*. C. Immunostaining of CD31⁺(red) endothelial cells and DCX⁺ (green) immature neurons in the whole-mount assembloid at day18 post-fusion. Nuclei are stained with DAPI (blue). *Scale bar, 200 µm*. D. Immunostaining of CD31⁺ (red) endothelial cells, Iba1^+^ (magenta) microglia, DCX⁺ (green) immature neurons in the whole-mount assembloid at d45. *Scale bar, 500 µm.* Artifacts outside the organoid were removed. E. Magnified region from (D) highlighting Iba1⁺ (magenta) cells associated with CD31⁺ (red) vessel-like structures. *Scale bar, 500 µm*. F. Magnified region from (D) showing Iba1⁺ (magenta) cells incorporated within DCX⁺ (green) neural tissue. *Scale bar, 500 µm*. G. uMAP of scRNA-seq data from day 60 assembloid (n=2) showing cells colored by cell population, alongside a stacked bar plot indicating the proportion of each cluster. H. Distribution of major cell populations in day 60 cerebral assembloids based on scRNA-seq analysis of two independent assembloids (n = 2). Neural lineage cells comprised 40.6% of the total population, glial cells 34.9%, stem/progenitor cells 17.2%, and mesenchymal cells approximately 3.3%. A-F: Representative images of four cerebral–vessel assembloids.

To further assess stability and stoichiometry of the tissue integration, at d60 we performed single-cell RNA sequencing (scRNA-seq) of fused assembloid (Figure 3 G-H), identifying that approximately 40.6% of cells were of neural lineage, 34.9% glial and only approximately 3.3% of total cells were mesenchymal cells (i.e. fibroblasts, pericytes, VEC, SMC and microglia). In addition, approximately 17.2% of cells were classified as stem cells, whose lineage identity could not be definitively assigned at this stage. Transcriptomic analysis showed that the microglial population expressed canonical microglial markers, including TMEM119, P2RY12, HEXB, and OLFML3, while simultaneously exhibiting a pronounced Type I (i.e., IFN-α/IFN-β) interferon-response signature characterized by expression of IFITM3, ISG15, IFI6, and related interferon-stimulated genes, indicating that these cells were in an activated rather than a homeostatic state.

Thus, assembloid generation led to better integration and prolonged persistence of activated microglia in COs in comparison to co-culturing COs with microglial progenitors. Nevertheless, the full integration of tissues of neuroectodermal and mesenchymal origin was not achieved and small proportion of mesenchymal cells observed by scRNA-seq at d60 suggests weak long-term stability of mesenchymal cells in CO assembloids.

Furthermore, CO–vascular assembloids demonstrated limited structural stability during long-term culture. Approximately 90 days after fusion, we observed progressive architectural changes, cyst formation, and a loss of structural integrity. These findings indicate that CO–vascular assembloids should be generated at developmentally appropriate time points tailored to the pathological process under investigation and used within an approximately 90-day experimental window.

### Prolonged *in vitro* culture leads to CO degeneration characterized by absent myelin, paucity of neurons and overwhelming reactive astrogliosis at organoid edge

As most human neurodegenerative diseases are age-related, aging represents a major risk factor for neurodegeneration. Therefore, we asked whether extended *in vitro* culture of fully matured myelinated COs recapitulates some aspects of age-related neurodegeneration, such as neuro-axonal loss, hypomyelination and reactive astrogliosis. We assessed COs age over 500 days by limited multiplex fluorescence microscopy (performed at d540).

In contrast to the organized, parallel NFL staining observed at the CO edge of myelinated d140 organoids (Figure 4A for donor BB026), this pattern was absent at d540 (Figure 4B for donor BB026 and higher magnification Figure S9 for BB011). Instead, NFL staining was localized only to deeper layers of the organoid, where long axons were predominantly oriented perpendicular to the edge (Figure S9C and S10). A prominent feature of aged COs was diffuse GFAP staining, which was particularly dense at the organoid rim (compare Figure 4C [d140] with Figure 4D [d540]; Figure S9B and S10B), effectively replacing the myelination layers visible at d98-d140. While we hypothesized that aged COs might become hypomyelinated, fluorescence microscopy revealed an almost complete loss of myelin proteins (MBP and PLP) by day 540 (Figure 4E *vs* Figure 4F). Despite this, OLIG2⁺ oligodendrocytes or oligodendrocyte precursor cells (OPCs) were still present at both the edge and the center of the COs (Figure 4E,F; Figure S10C).

**Figure 4.**
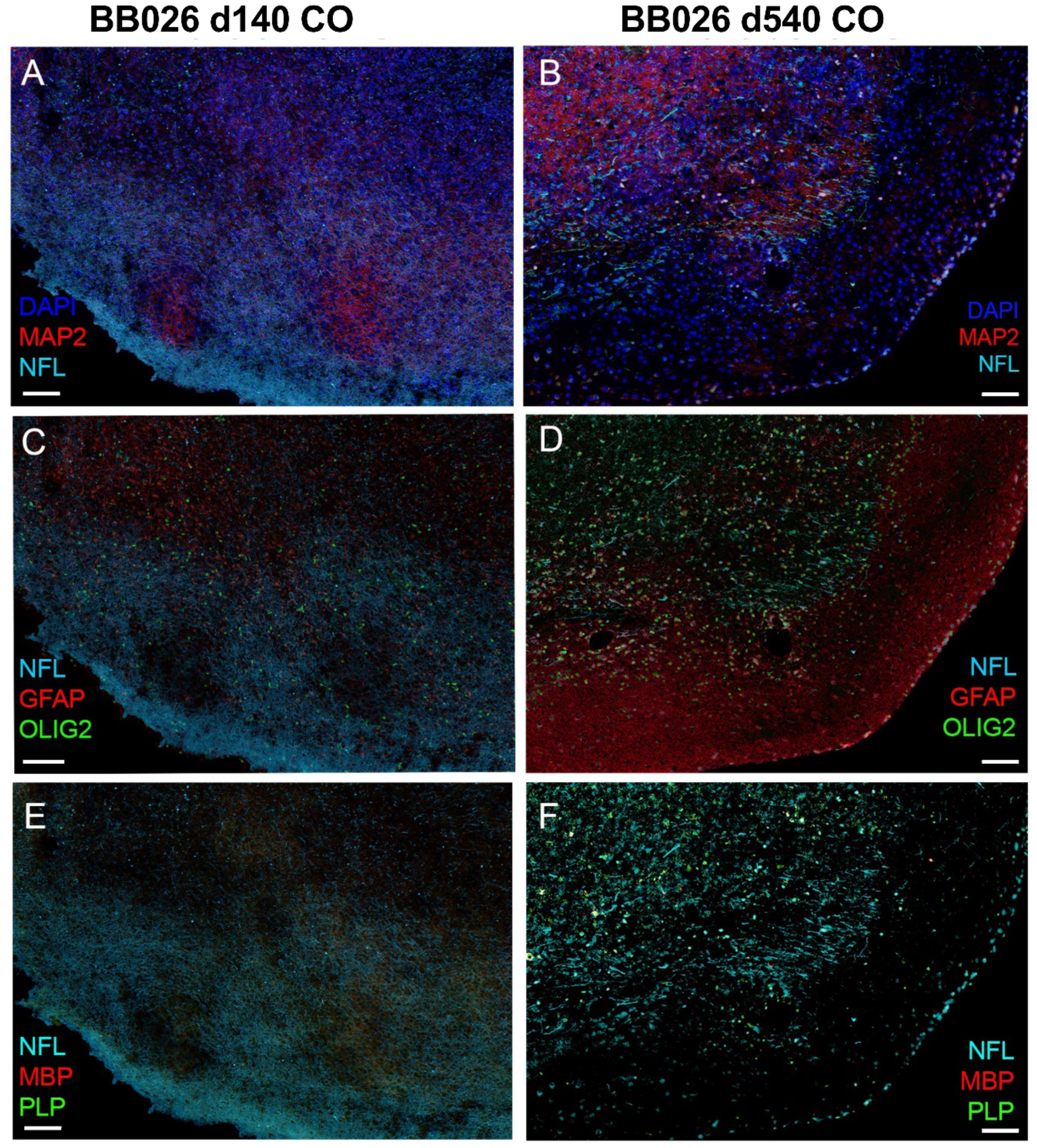
Age-related neurodegeneration in cerebral organoids from d140 to d540. A. Immunostaining of NFL⁺ (cyan) and MAP2⁺ (red) cells in cryosection from 140-day-old organoidNuclei are stained with DAPI. *Scale bar, 100 µm*. B. Immunostaining of NFL⁺ (cyan) and MAP2⁺ (red) cells in cryosection from 540-day-old organoid. Nuclei are stained with DAPI. *Scale bar, 100 µm*. C. Immunostaining of NFL+ (cyan), GFAP+ (red) and OLIG2+ (green) cells in cryosection from 140-day-old organoid. Nuclei are stained with DAPI. *Scale bar, 100 µm*. D. Immunostaining of NFL+ (cyan), GFAP+ (red) and OLIG2+ (green) cells in cryosection from 540-day-old organoid. Nuclei are stained with DAPI. *Scale bar, 100 µm*. E. Immunostaining of NFL+ (cyan), MBP+ (red) and PLP+ (green) cells in cryosection from 140-day-old organoid. Nuclei are stained with DAPI. *Scale bar, 100 µm*. F. Immunostaining of NFL+ (cyan), MBP+ (red) and PLP+ (green) cells in cryosection from 540-day-old organoid. Nuclei are stained with DAPI. *Scale bar, 100 µm*. Representative image of two 140-day-old cerebral organoids (donor BB026 & BB028) and three 540-day-old-organoids (donor BB11, BB026 & BB028).

Quantitative analysis of immunofluorescence staining confirmed the histological observations, demonstrating a loss of detectable MBP- and PLP-positive area and a marked increase in GFAP-positive area in day 540 organoids (Figure S6).

Thus, during long-term culture, COs derived from both healthy volunteers and MS patient consistently display structural hallmarks of aging (i.e., hypomyelination, severe neuroaxonal loss, and a predominance of reactive astrocytes forming a dense GFAP+ rim at the outer edge), demonstrating that these alterations reflect a generalized cellular aging process rather than a disease-specific phenotype.

### Immune cells interact with and incorporate into superficial layers of COs

CNS infiltration by immune cells has been described in most neurodegenerative diseases. However, studying what role these infiltrating immune cells play in disease process and how they interact with CNS cells has been exceptionally difficult in humans. Therefore, we wanted to explore how autologous resting and activated immune cells (i.e., peripheral blood mononuclear cells, PBMC) interacted with and migrated into COs.

To optimize the conditions and duration for co-culturing autologous PBMC with COs, we employed live imaging techniques. We co-cultured cerebral organoids (derived from healthy donors BB026 and BB028) at day 240 with 2 x 10^6^ resting or polyclonally activated PBMC. Within 24h, the autologous PBMC interacted with the superficial layers of the COs. Live imaging confirmed that both resting and activated PBMC successfully engaged with the superficial layers during this timeframe. However, resting PBMC primarily bounced off the edges of the organoids (data not shown), while activated PBMC incorporated into the superficial layers of the organoid structure (Figure S11, Video2).

We repeated autologous PBMC-CO co-culture experiments using 540-day-old organoids derived from three donors: one MS patient (BB011) and two healthy individuals (BB026 and BB028). After 24 hours of co-culture, the MS patient incorporated 443 CD3^+^ T cells, 125 CD20^+^ B cells, and 91 CD68^+^ monocytes/macrophages into a 10 µm section of the organoid (Figure 5A, Figure S12). In comparison, the COs of equivalent size from the healthy donors incorporated on average 108 ± 24.04 CD3^+^ T cells, 49.5 ± 7.70 CD20^+^ B cells, and 29.5 ± 7.78 CD68^+^ monocytes/macrophages (Figure 5B,C).

**Figure 5.**
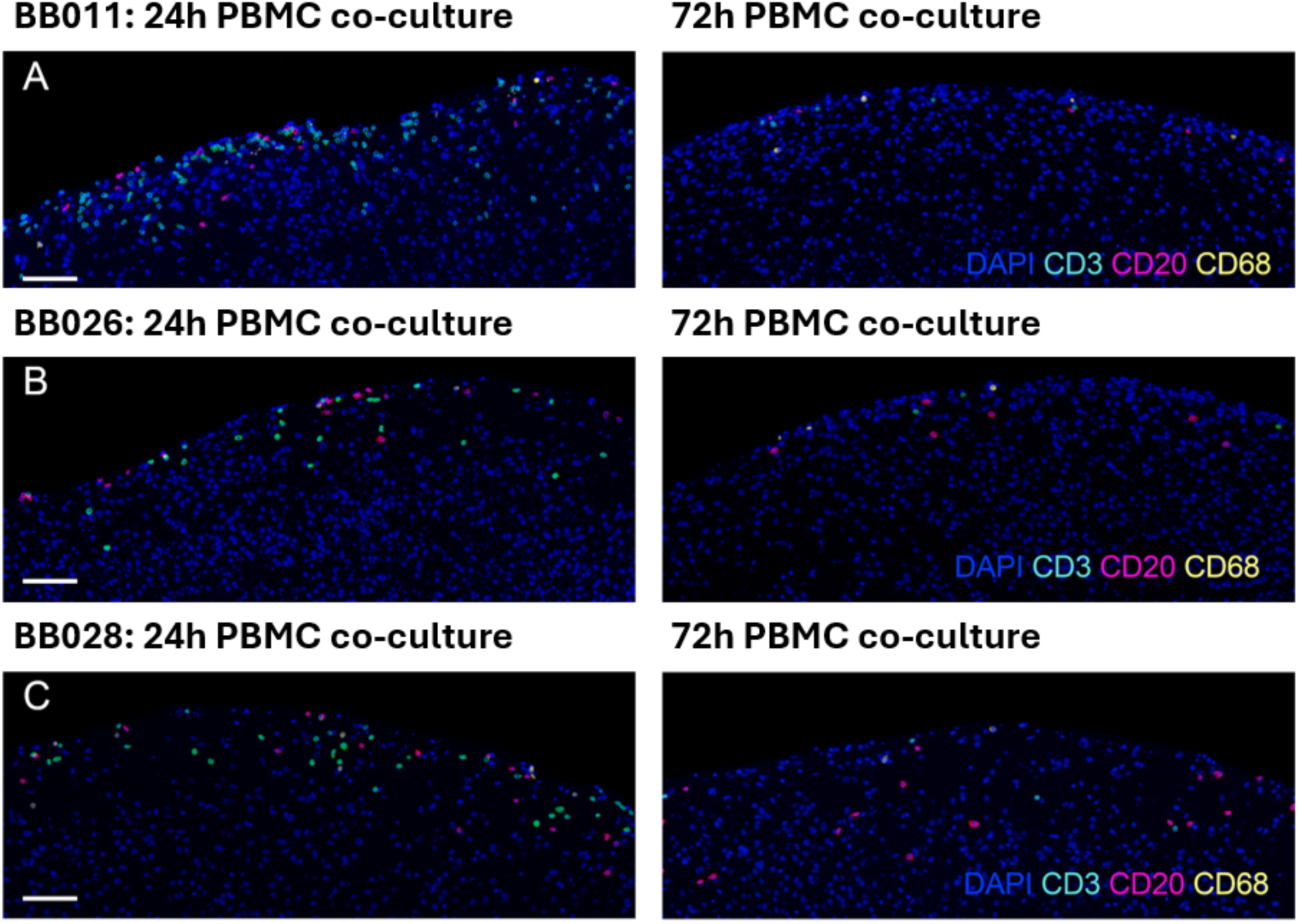
Immune cell infiltration into cerebral organoids following 24- and 72-hour co-culture with autologous activated PBMC. A. Immune cell incorporation into MS donor-derived CO (BB011) 24h (LEFT) and 72h (RIGHT) after co-culture with activated PBMC. Immune cell composition was determined via CD3+ (cyan) T-cells, CD20+ (magenta) B-cells, and CD68+ (yellow) macrophage. *Scale bar, 100 µm*. B. Immune cell incorporation into healthy donor-derived CO (BB026) 24h (LEFT) and 72h (RIGHT) after co-culture with activated PBMC. Immune cell composition was determined via CD3+ (cyan) T-cells, CD20+ (magenta) B-cells, and CD68+ (yellow) macrophage. *Scale bar, 100 µm*. C. Immune cell incorporation into healthy donor-derived CO (BB028) 24h (LEFT) and 72h (RIGHT) after co-culture with activated PBMC. Immune cell composition was determined via CD3+ (cyan) T-cells, CD20+ (magenta) B-cells, and CD68+ (yellow) macrophage. *Scale bar, 100 µm*. One cerebral organoid per donor was analyzed by immunofluorescence at each time point.

Surprisingly, after 72 hours of co-culture, the number of incorporated PBMC significantly decreased for all three donors. For the MS patient, we observed 46 CD3^+^ T cells, 14 CD20^+^ cells, and 11 CD68^+^ cells (Figure 5A). In contrast, the healthy donors had average counts of 27 ± 15.55 CD3^+^ T cells, 42 ± 33.94 CD20^+^ cells, and 15.5 ± 3.53 CD68^+^ cells (Figure 5 B,C).

We conclude that all immune cells readily interact with autologous COs, but (polyclonally) activated immune cells have clear edge in infiltrating into superficial layers of organoid tissue. Nevertheless, we observed that CO infiltration by immune cells was transient, and it decreased robustly after 72h of co-culture in all 3 donors.

The structural and behavioral congruency of the immune cells across all donors suggests that this infiltration reflects a baseline physiological interaction between peripheral immune cells and the CNS. We therefore caution against over-interpreting baseline differences in absolute cell numbers between individual donors as an MS-specific trait, as these characterization experiments were not designed to elucidate disease mechanisms.

### Activated PBMC establish a pro-inflammatory cytokine environment in CO co-cultures

To characterize the inflammatory environment associated with organoid–immune cell interactions, cytokine concentrations were measured in conditioned medium from organoids cultured alone, organoids co-cultured with activated autologous PBMC, and activated PBMC cultured alone (Figure S13; donors BB026, BB028, and BB007). Organoids cultured alone secreted little or no detectable GM-CSF, IL-1β, IL-6, IL-10, INF-γ or TNF-α, whereas activated PBMC produced high concentrations of these inflammatory cytokines. Cytokine concentrations in co-culture supernatants closely resembled those of activated PBMC alone, indicating that activated PBMC were the principal source of inflammatory cytokines in the co-culture. In contrast, VEGF-A concentrations remained relatively high in organoid cultures and co-cultures but were low or undetectable in activated PBMC alone, indicating that VEGF-A was primarily produced by the organoids. Together, these findings demonstrate that activated PBMC establish a pro-inflammatory milieu while the organoids maintain production of tissue-derived trophic factors, thereby creating an experimental environment that models inflammatory insults to human neural tissue.

### Single-cell spatial transcriptomics confirms and expands observations that COs in long-term culture (d750) display structural and transcriptomic changes that closely overlap with the aging human brain phenotype

To validate and extend our immunofluorescence findings, we performed high-content spatial transcriptomics analysis using the Xenium Prime 5K Human Pan Tissue & Pathways Panel, supplemented with 100 custom CNS lineage-enriched genes for a total of 5,100 targets (Supplementary Data 1). To maintain continuity with previous experiments, day 750 (d750) COs from healthy donors BB026 and BB028 were analyzed. Day 200 (d200) COs were generated from a newly included multiple sclerosis (MS) patient donor (BB007).

This cross-sectional design had two primary objectives. First, we sought to determine whether longitudinal features of CO maturation and myelination could be cross-sectionally reproduced by comparing young (d200) and aged (d750) organoids from distinct batches. Second, because extended culture introduces some variability in organoid size and necrotic core development, we used the multiple organoids (i.e., biological replicates) available from donor BB007 to formally evaluate within-subject cellular and transcriptional heterogeneity. This characterization establishes a baseline for statistical powering of future organoid-based assays.

Cellular and transcriptional compositions across four distinct d200 COs generated from donor BB007 were highly reproducible (Figure S14). This high within-subject reproducibility contrasted sharply with the compositional and transcriptional differences observed between young (d200) and aged (d750) organoids (Figure 6). Consistent with our longitudinal findings, aged organoids exhibited expanded astrocyte and radial glia populations alongside severely contracted proportions of progenitors/OPCs, mature neurons, and oligodendrocytes (Figure 6A upper panels and 7A,B left panels).

**Figure 6.**
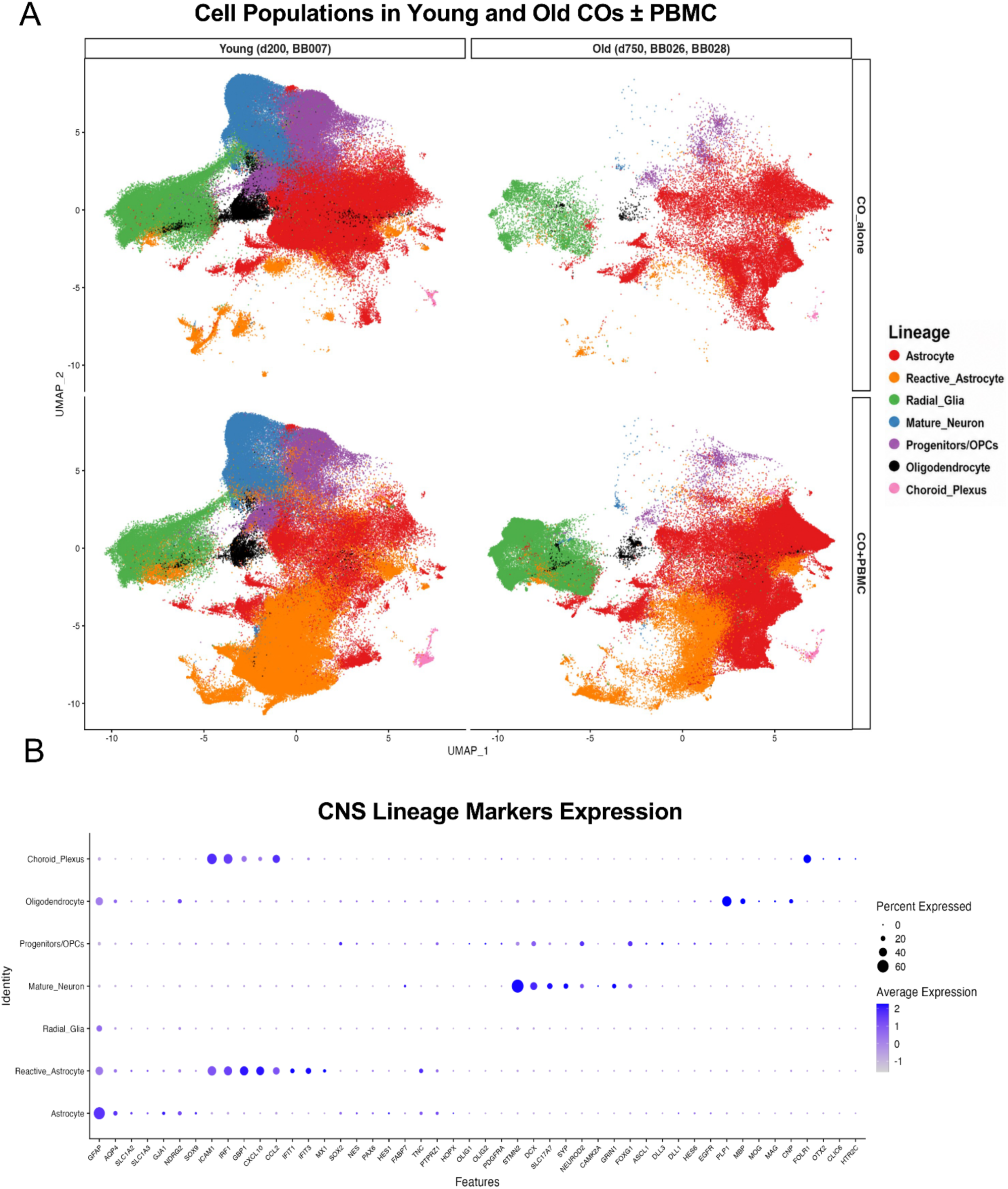
Distribution of major CNS cell populations in young and aged cerebral organoids cultured alone or following co-culture with activated PBMC. A. UMAP visualization of integrated single-cell transcriptomes from cerebral organoids cultured alone (CO alone) or co-cultured with activated PBMC (CO + ActPBMC). Young organoids (day 200; donor BB007) included n = 4 organoids per condition (CO alone, n = 4; CO + ActPBMC, n = 4). Aged organoids (day 750; donors BB026 and BB028) included n = 2 organoids for the CO alone condition (one organoid from each donor) and n = 4 organoids for the CO + ActPBMC condition (two organoids from each donor). Consequently, the integrated CO + ActPBMC dataset contains a greater total number of cells than the CO alone dataset for the aged organoids. Young organoids are shown in the left column, and aged organoids are shown in the right column. The top row represents organoids cultured alone, and the bottom row represents organoids following ActPBMC co-culture. Cells are colored according to their annotated major CNS cell lineages. Cell identities were assigned based on canonical marker gene expression as described in the Methods. B. Dot plot showing the expression of representative marker genes used to annotate the major CNS cell lineages. Dot size indicates the percentage of cells expressing each gene, and color intensity represents the average normalized expression level.

**Figure 7.**
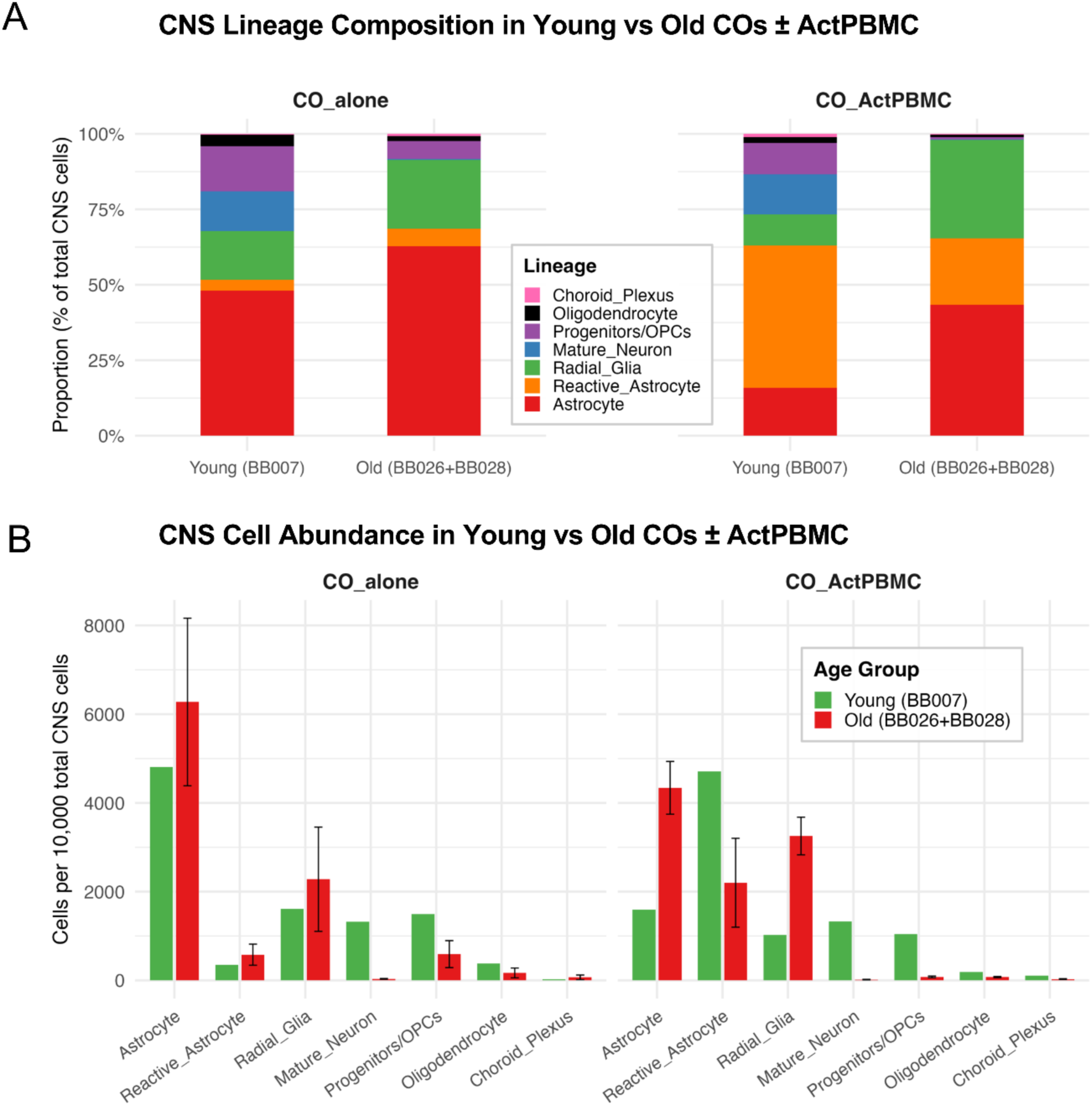
Remodeling of CNS cell composition during cerebral organoid aging. A. Relative proportions of major CNS cell populations in young (BB007) and aged (BB026 and BB028 combined) cerebral organoids cultured alone (CO alone; BB007, n = 4 organoids; BB026 and BB028, n = 1 organoid per donor (2 organoids total)) or following co-culture with activated PBMC (CO + ActPBMC; BB007, n = 4 organoids; BB026 and BB028, n = 2 per donor (4 organoids total)). Cell proportions are expressed as the percentage of the total CNS cell population. B. Absolute abundance of each CNS cell lineage normalized to organoid size (cells per 10,000 total CNS cells) in young and aged organoids under each culture condition. Bars represent mean ± standard error (SE). CNS cell populations were annotated based on established lineage-specific transcriptional markers, as described in the Methods.

GFAP+ astrocytes localized to the outer periphery of the organoids and dominated cellular composition of aged organoids (Figure 8A). Consequently, we resolved highly significant, lineage-specific transcriptional differences between young and aged organoids, particularly within astrocytes and reactive astrocytes (all highlighted changes reached p<10^-20^; Figure 8B,C).Top differentially expressed transcripts included markers associated with inflammatory and neurotoxic astrocytes in neurodegenerative diseases, such as *CHIT3L1* (∼13.9-fold enriched) and *SERPINA3* (∼36.8-fold enriched) ^25–28^ (Figure 8A-C). While this neurotoxic phenotype typically emerges in response to microglia-^27^ or PBMC^26^-derived soluble inflammatory mediators, aged CO astrocytes developed this profile in the absolute absence of immune cells, pointing to cell-autonomous or alternative induction mechanisms.

**Figure 8.**
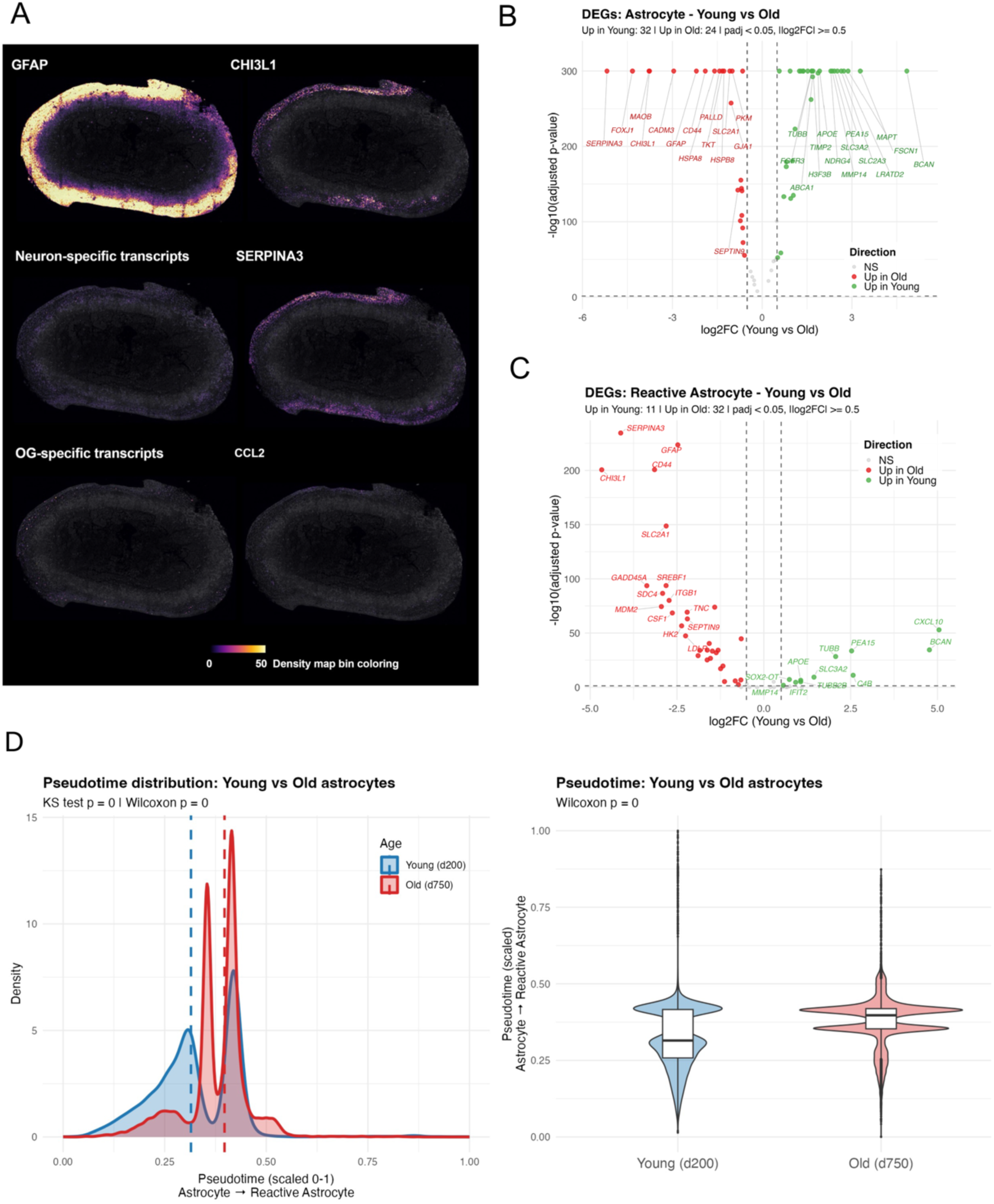
Aged cerebral organoids exhibit astrocyte reactivity and progression toward a reactive transcriptional state. A. Representative Xenium spatial transcriptomic maps of aged (day 750) cerebral organoids showing the spatial distribution of astrocyte (GFAP), reactive astrocyte (CHI3L1 and SERPINA3), neuronal, oligodendrocyte (OG)-associated, and inflammatory (CCL2) transcripts. Signal intensity represents transcript density. B. Differential gene expression analysis of astrocytes from young (BB007, *n* = 4 organoids) and aged (BB026 and BB028, *n* = 2 organoids) cerebral organoids. Positive log₂ fold change indicates higher expression in young organoids, whereas negative log₂ fold change indicates higher expression in aged organoids. Differentially expressed genes were defined as |log₂ fold change| ≥ 0.5 and adjusted *P* value (padj) < 0.05. All highlighted changes reached p<10^-20^. C. Differential gene expression analysis of reactive astrocytes from young (n= 4) and aged (n=2) cerebral organoids. Differentially expressed genes were defined as |log₂ fold change| ≥ 0.5 and adjusted *P* value (padj) < 0.05. All highlighted changes reached p<10^-20^. D. Pseudotime analysis of the astrocyte-to-reactive astrocyte trajectory in young (n=4) and aged (n=2) cerebral organoids. Density distributions (left) and violin plots (right) show pseudotime progression for each group. Statistical significance was assessed using the Kolmogorov–Smirnov test and the Wilcoxon rank-sum test.

The ciliogenesis transcription factor *FOXJ1* was ∼19.7-fold enriched in aged astrocytes, suggesting a shift toward an ependymal-like identity. This was accompanied by upregulation of the primary astrocytic gap junction gene *GJA1* (connexin-43), suggesting enhanced intercellular coupling. Additionally, upregulation of heat shock proteins (*HSPA8*, *HSPB8*) indicated proteostatic stress, while coordinated upregulation of *PKM*, *TKT*, *SLC2A1*, and *PFKFB3* demonstrated a robust glycolytic shift (Warburg-like effect) in aged astrocytes (Figure 8 B,C; Supplementary Data 2).

In contrast, astrocytes in young organoids displayed a healthy, neurotrophic state characterized by high expression of BCAN (brevican; a brain-specific proteoglycan essential for perineuronal nets and synaptic stability), the maturation markers NDRG2 and FGFR3, and the neuroprotective factor NDRG4. Young astrocytes also differed from their aged counterparts by elevated levels of APOE and ABCA1, which mediate cholesterol transport to neurons and oligodendrocytes, as well as QKI (RNA-binding protein critical for myelination). Finally, they also expressed a distinct metabolic transcriptomic profile (FAM107A, GPI, BSG, SLC3A2, ATP5F1A), associated with active mitochondrial ATP generation and lactate shuttling to neurons (Figure 8 B,C and Supplementary Data 2).

Pseudotime analysis confirmed that astrocytes from aged organoids progressed significantly further along the chronological trajectory compared to those from young organoids (Figure 8D, Figure S15B).

Thus, we conclude that resting astrocytes in young organoids exhibit a transcriptome consistent with neurotrophic and myelination-supportive physiological functions. Conversely, aged astrocytes lose these supportive roles as they adopt a stressed and neurotoxic phenotype.

### CNS cells in aged organoids adopt a transcriptionally senescent phenotype

To determine whether the transcriptional differences observed between young and aged astrocytes overlap with a senescent phenotype and extend to other CNS lineages, we evaluated the lineage-specific expression of 50 transcripts associated with the senescence-associated secretory phenotype (SASP; Figure S16. We observed robust upregulation of most SASP-linked transcripts across CNS cell lineages in aged organoids. Notable exceptions included *APOE* and *MMP14*, which were preferentially upregulated in young CNS cells, consistent with their established physiological roles in baseline CNS tissue homeostasis and development.

Thus, we conclude that in addition to reproducing characteristic age-related cellular composition changes of human brain, aged COs also reproduced transcriptional senescence-associated phenotype across all CNS lineages compared to d200 COs.

### Incorporation of autologous immune cells into aged COs is associated with a robust type II interferon signature in CNS cells and a decreased proportion of oligodendrocytes actively transcribing myelin genes

While previous longitudinal experiments demonstrated that both young and aged organoids transiently incorporate immune cells from autologous activated PBMC into outer cellular layers, we sought to generate pilot, hypothesis-generating data to characterize the responses of CNS cells to this immune cell infiltration.

First, when comparing changes in cellular composition of CNS cells in COs cultured alone and COs cultured with activated autologous PBMC, we observed highly congruent expansion of reactive astrocytes and radial glia in both old and young organoids, with associated contraction of resting astrocytes and progenitors/OPCs (Figure 6 A and 7A,B, compare the right panels with the corresponding left panels; Figure 9A). Consistently, across all organoids studied (4 young and 4 aged) astrocytes and progenitors/OPCs exhibited the strongest cellular and transcriptional responses (Figure 9A-C) to activated PBMC. This was characterized by a robust type II IFN-response signature and a phenotypic shift from resting to reactive astrocyte subpopulations. Transcriptionally, reactive astrocytes showed highly congruent changes between aged and young organoids, strongly upregulating classical IFN-γ response genes (*TAP1*, *IRF1*, *GBP1*, *IFIT2*, *WARS*, *APOL2*) and immune activation/adhesion molecules (*ICAM1*, *CCL2*, *CD44;* Figure 9C, Figure S17). Consistent with previous studies demonstrating that activated PBMC release factors that induce a neurotoxic A1 astrocyte phenotype^26^ both young and aged astrocytes upregulated *SERPINA3*, as well as *TCN*, *TAGLN2*, and *ADM*, which participate in injury response and extracellular matrix (ECM) remodeling (Figure 9C, Figure S17).

**Figure 9.**
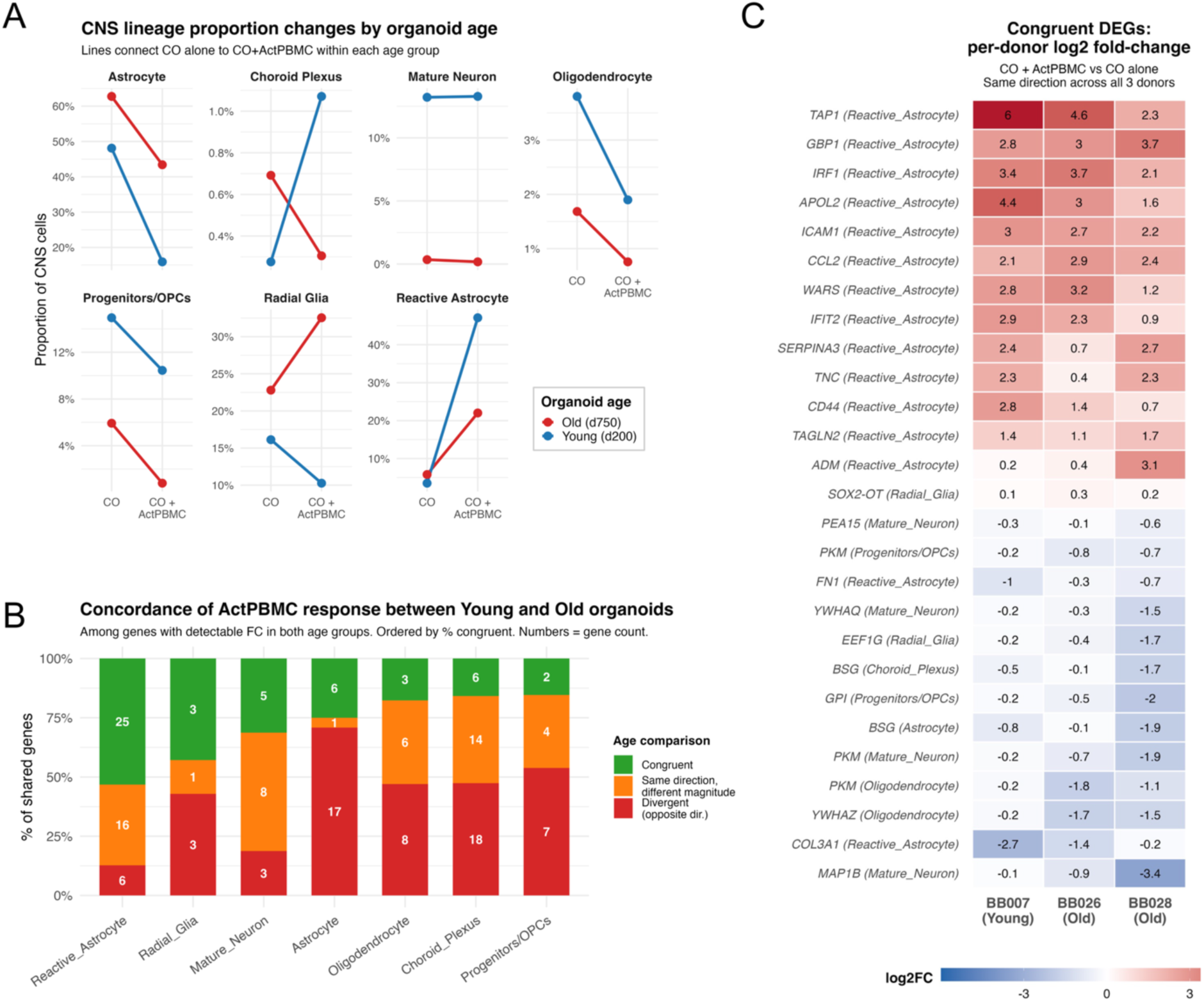
Young and aged cerebral organoids exhibit distinct cellular and transcriptional responses to activated autologous PBMC. A. Changes in the relative abundance of major CNS cell populations following co-culture with activated PBMC (ActPBMC) in young (day 200) and aged (day 750) cerebral organoids. Lines connect matched organoids cultured alone (CO alone) and after co-culture with ActPBMCs (CO + ActPBMC). B. Concordance analysis comparing ActPBMC-induced transcriptional responses between young and aged cerebral organoids across major CNS cell populations. Shared differentially expressed genes were classified as congruent (same direction and similar magnitude), same direction with different magnitude, or divergent (opposite direction). Numbers within bars indicate the number of shared differentially expressed genes. C. Heatmap of representative genes consistently differentially expressed following ActPBMC co-culture across all three donors (BB007, BB026, and BB028). Heatmap values represent per-donor log₂ fold changes for ActPBMC co-culture relative to organoid-only controls. Differentially expressed genes were defined as |log₂ fold change| ≥ 0.5 and adjusted *P* value (padj) < 0.05. Young organoids (day 200; donor BB007) included n = 4 organoids per condition (CO alone, n = 4; CO + ActPBMC, n = 4). Aged organoids (day 750; donors BB026 and BB028) included n = 2 organoids for the CO alone condition (one organoid from each donor) and n = 4 organoids for the CO + ActPBMC condition (two organoids from each donor).

While oligodendrocytes, as proportion of CNS cells also decreased in both young and old COs, their transcriptional responses to activated PBMC differed in age-related manner. This was true also for choroid plexus cells, and progenitors/OPCs (Figure 9B, Figure S18).

We focused our analysis on oligodendrocytes, which selectively in aged organoids (two donors, four organoids) congruently downregulated 21 genes (Figure 10A). Most of these downmodulated genes are explicitly linked to myelination, including structural myelin proteins (*MBP*, *PLP1*, *CNP*), the orphan receptor *GPRC5B* (highly expressed in myelinating oligodendrocytes), *SPP1* (osteopontin, which promotes oligodendrocyte differentiation and remyelination), and *FDFT1* (required for cholesterol biosynthesis). Furthermore, the downregulation of metabolic transcripts (*PKM*, *SLC2A1*, *ATP1A1*), lysosomal membrane proteins (*LAMP1*, *LAMP2*), the translation elongation factor *EEF1G*, and scaffolding proteins *YWHAZ* and *YWHAQ* (which regulate stress signaling and inhibit apoptosis) points to severe metabolic failure and proteostatic stress (Figure 10B and Supplementary Data 2).

**Figure 10.**
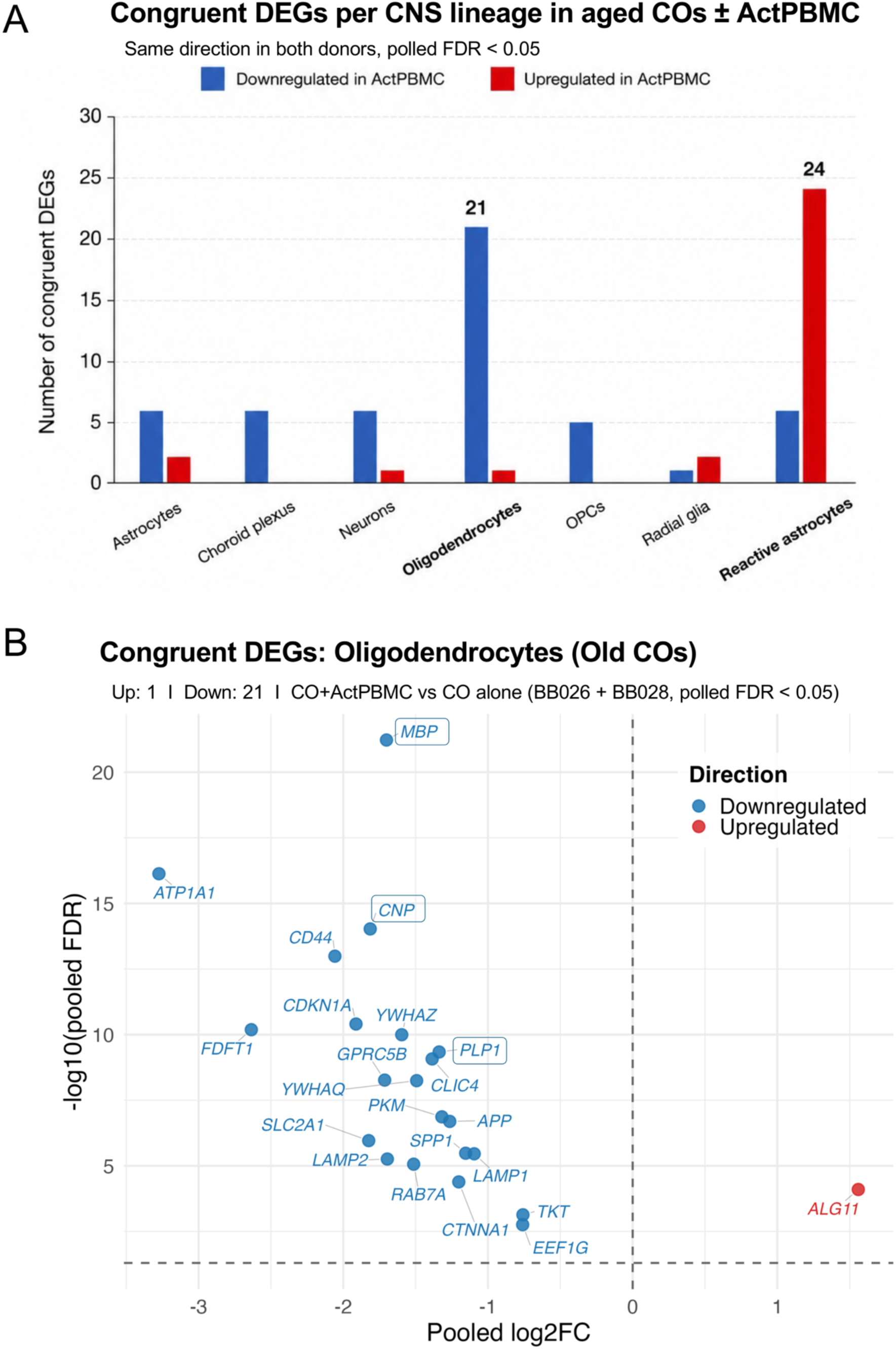
Reactive astrocytes and oligodendrocytes exhibit the strongest conserved transcriptional responses to activated PBMC co-culture in aged cerebral organoids. A. Number of congruent differentially expressed genes (DEGs) in aged organoids (BB026 and BB026) identified in each CNS cell lineage following co-culture with activated PBMC (CO + ActPBMC, n=4) relative to organoids cultured alone (CO alone, n=2). Congruent DEGs were defined as genes that were significantly differentially expressed (pooled FDR < 0.05) and changed in the same direction in both aged donors (BB026 and BB028). Upregulated and downregulated genes are shown separately for each cell lineage. B. Volcano plot of congruent DEGs in oligodendrocytes following ActPBMC co-culture. The x-axis represents pooled log₂ fold change (CO + ActPBMC (n=4) versus CO alone (n=2)), and the y-axis represents −log₁₀(FDR). Differentially expressed genes were defined as |log₂ fold change| ≥ 0.5 and pooled FDR< 0.05. Myelin-associated genes are highlighted.

We also noted that myelin transcripts (predominantly *MBP* > *PLP1*, which were abundant in resting organoids) frequently aligned into extracellular linear arrays, likely representing oligodendrocyte processes (Figure S19-21). Consequently, we developed an automated pipeline to identify these linear structures (see Methods) and compared their abundance, cumulative length, and length distributions between control organoids and those co-cultured with activated PBMC (Figure S21). These data demonstrated a consistent reduction in both the total number and length of these extracellular structures across aged organoids following activated (but not resting) PBMC co-culture.

Notably, while activated PBMC co-culture decreased the overall proportion of oligodendrocytes in young organoids, the surviving oligodendrocytes maintained a distinct transcriptomic profile compared to those in aged organoids post-co-culture (Figure S18). Although a subset of transcripts was congruently downregulated across both cohorts, young oligodendrocytes uniquely demonstrated a significant upregulation of *MBP* and *CNP*.

In conclusion, aged organoids successfully recapitulated key features of age-related neurodegeneration, including a robust loss of neurons and axons, oligodendrocyte depletion, and extensive astrogliosis. Most striking was the autonomous induction of a neurotoxic astrocyte phenotype in the complete absence of autologous immune cells or microglia. Co-culture with autologous activated PBMC triggered strong type II interferon signaling in reactive astrocytes across all cohorts, culminating in oligodendrocyte loss in young organoids and profound oligodendroglial injury/loss in aged organoids. These findings suggest that oligodendrocytes, particularly within an aged microenvironment, may be uniquely susceptible to immune-mediated damage, irrespective of disease status.

## DISCUSSION

While we did not succeed in generating *in vitro* model that faithfully recapitulates the cellular diversity and stoichiometry of fully developed human CNS, we have substantially advanced this ambitious and difficult aim. Specifically, optimizing hiPSC numbers for embryoid body formation robustly delayed the development of tissue hypoxia and the resulting necrotic core, extending time available for modeling neural-immune interactions during CO maturation and aging. We successfully generated myelinated COs and demonstrated that following the induction of oligodendrocyte differentiation, myelination evolves over several months, peaks, and subsequently degenerates during prolonged (> 1-2 year) culture. We also characterize an aging-like CO phenotype where both oligodendrocytes and neurons are progressively replaced by reactive astrocytes. Comparing COs at the peak of neuronal development and myelination (day 140) with those cultured for more than 500 days revealed a paucity of myelinated axons, alongside cell bodies confined to a progressively thinning rim layer. This layer is separated from the culture medium by dense astrocytic processes exhibiting high GFAP expression. CNS cells in these aged organoids transcribe senescence-associated genes. Furthermore, reactive astrocytes assume a previously described “neurotoxic” transcriptional phenotype that develops independently of immune cells or their soluble mediators. Thus, while soluble immune mediators can induce neurotoxic astrocyte phenotype ^26,27^, our data indicate that astrocytes can adopt this state via alternative, thus far undescribed mechanism(s).

Although we can only hypothesize what causes CO “aging”, it may relate to the lack of mesenchymal cells that in healthy CNS mediate both barrier functions and nutrient delivery. In the absence of barrier epithelium (i.e., CNS epithelial and endothelial cells) featuring tight junctions and selective solute transfer systems, and lacking subepithelial layer of mesenchymal phagocytes (i.e., macrophages and microglia), astrocytes appear to assume compensatory, excessive barrier-forming functions. This is supported by both their structural characteristics and transcriptional signatures. However, this phenotypic shift fails to replace the physiological functions normally mediated by CNS mesenchymal cells and may ultimately trigger neurodegeneration ^26,27^, although this interpretation remains speculative.

We also investigated the integration of mesenchymal cells into COs and confirmed that co-culturing COs with hiPSC-derived microglia progenitors leads to weak and only transient incorporation of these innate immune cells. Instead, generating assembloids by combining COs with hiPSC-derived vascular organoids appears more promising; beyond serving as a source of microglia, vascular organoids provide other mesenchymal cells that play essential roles in chronic CNS diseases. However, we did not achieve complete integration between COs and vascular organoids; instead, the vascular tissue formed only isolated islands within the CO-vascular assembloids. Furthermore, our preliminary observations indicate that, like un-assembled COs, these assembloids also deteriorate in prolonged culture (after approximately 90 days). This suggests that COs and assembloids must be utilized within a limited, optimal experimental window tailored to the specific disease under investigation. For example, pediatric conditions such as genetic demyelinating diseases are best studied during the active myelination stage, whereas age-related neurodegenerative diseases should be modeled during the declining phase of CO development.

As a laboratory focused on human neuroimmunological diseases such as multiple sclerosis (MS), we aimed to develop COs as an *in vitro* model of human neuro-immune interactions, which remain exceedingly difficult to study *in vivo*. We investigated the extent to which neuro-immune interactions are influenced by baseline heterogeneity, specifically donor phenotype, technical variations in CO generation, PBMC activation states, and the timing of CO-PBMC co-culture. Accounting for these variables is essential, as future applications of this model must tightly control confounding heterogeneity and interpret findings within the specific constraints of the experimental conditions. Reassuringly, utilizing hiPSCs from multiple donors, generating COs across independent batches, varying the number of seeded progenitors, and initiating co-cultures at different time points relative to PBMC activation and CO development introduced only minor variations in CO size, structural organization and cellular composition. The true biological replicates tested at d200 showed highly uniform distributions of CNS cells in resting organoid and reproducible cellular and transcriptional responses to activated PBMC co-cultures.

Crucially, this intra- and inter-donor heterogeneity did not alter our primary experimental conclusions: polyclonally activated autologous PBMC interact with and infiltrate COs, albeit transiently. The CNS cells recognize this infiltration by displaying strong type II IFN (i.e., IFN-γ)-mediated transcriptional signature. Whether infiltrating immune cells can persist long-term within the organoids and adopt a tissue-resident phenotype remains to be determined. Furthermore, while immune cell infiltration in both young and aged COs decreased proportions of oligodendrocytes among all CNS cells and aged COs further displayed decreased expression of myelin transcripts within surviving oligodendrocytes, we did not directly evaluate formal demyelination on a protein/lipid level. Because we observed lack of myelin proteins by fluorescence microscopy already at d540, and extreme paucity of neurons and NEFL transcripts in >700 days aged COs, it is much more likely that these aged COs were not myelinated and the linear structures of myelin transcripts represent oligodendroglial processes searching for axonal contacts rather than myelinated axons. Future studies will need to explore this phenomenon in detail. We also cannot causally link oligodendrocyte or myelin transcripts loss to a specific immune mechanism, meaning the transcriptomic analyses presented here should be considered strictly hypothesis-generating.

To what degree the *in vitro* conditions tested in these experiments (e.g., immune cell-to-organoid stoichiometry, or the type and duration of immune cell activation) reproduce what CNS cells encounter in human disease remains unclear. Nevertheless, alternative models of neuroinflammatory disease are similarly, if not more, artificial. These include actively induced experimental autoimmune encephalomyelitis (EAE) using myelin epitopes emulsified in complete Freund’s adjuvant augmented by pertussis toxin injection, adoptively transferred EAE where unphysiological quantities of *in vitro*-expanded myelin-specific T cells are transferred, or transgenic animals where the vast majority of T cells (and occasionally B cells) recognize a single myelin epitope.

EAE models induce pathogenic T cells by breaking immune tolerance, e.g., by providing strong pathogen associated molecular patterns, such as complete Freund’s adjuvant, alongside T cell activating signals. But tolerance mechanisms seems to be preserved in people with MS (pwMS), where pathogenic role of autoreactive T cells was only proven under iatrogenic condition when clinical trial of altered peptide ligand inadvertently expanded MBP83-98-specific CD4+ T cells >1000 fold.^29^ However, this expansion occurred exclusively in pwMS harboring MHC-II alleles that bind MBP83-98-too weakly to trigger their deletion during central tolerance. Thus, both EAE and MS data indicate that inducing sterile, T cell-mediated autoimmunity against normal CNS tissue is exceptionally difficult.

While we cannot entirely exclude the possibility that oligodendrocyte injury in our CO–immune co-cultures was driven by antigen-specific recognition by CO-infiltrating T cells, we consider this highly unlikely. First, we observed a loss of MBP+/PLP1+ oligodendrocytes in CO–PBMC co-cultures derived from healthy donors who lacked any evidence of CNS pathology. Second, oligodendrocyte injury and the transcriptional inhibition of myelination occurred within 24 hours of co-culture. In the absence of prior CNS injury, any autoreactive T cells present within the peripheral repertoire of these healthy donors would almost certainly exhibit a naive phenotype. Within a 24-hour sterile co-culture window, it is virtually impossible for naive autoreactive T cells to become fully activated, differentiate, and directly kill oligodendrocytes or secrete the high levels of cytokines required to destroy them indirectly.

Instead, we consider it far more likely that the observed decrease in MBP+/PLP1+ oligodendrocytes resulted from a selective vulnerability of oligodendrocytes to an unexpectedly robust induction of interferon signaling and innate immune pathways across most CNS cells upon co-culture with autologous PBMC. Notably, PBMC co-culture experiments using day 200 organoids reproduced this IFN-γ signaling response of reactive astrocytes, demonstrated approximately 30% loss of oligodendrocytes, but intact myelin transcripts in remaining oligodendrocytes. This difference suggests that the oligodendrocyte response varies depending on organoid age. Consequently, this observation strengthens the conclusion that aged COs can serve as a valuable model for recapitulating key features of brain aging, provided future studies utilize enough donors to encompass population variance.

### Limitations of the study

The primary technical limitations of this study include the current absence of a stable microglia population and the lack of long-term integration of autologous vessel organoids within the developing assembloids. Furthermore, the model lacks directional axonal growth and exhibits limited overall axonal length. *In vivo*, axonal directionality and length are driven and sustained by active electrical signaling circuits, such as motor or cortical pathways. The absence of patterned, experience-dependent neuronal activity in this in vitro system likely impacts long-term neuronal health and myelination.

We also observed unconstrained growth and a reactive, neurotoxic phenotype in astroglial cells, which formed a dense network of GFAP+ processes at the organoid periphery, potentially due to the absence of structural epithelial barriers. This reactive network may contribute to the involution of myelinated axons observed at the outer organoid boundary between days 98 and 140. Furthermore, because assembloid generation was restricted to early stages of CO development, we could not evaluate structural integration at later time points. The transcriptional and structural features of astrocytes in aged COs suggest that neurodegeneration may be partly driven by the lack of a functional CNS barrier and nutrient delivery network, which subsequently triggers reactive astrogliosis. Testing this hypothesis will require generating assembloids at later stages of CO maturation (e.g., days 80–120, during peak maturation and myelination) and optimizing their long-term culture survival.

The PBMC co-culture experiments aimed to determine whether autologous immune cells could infiltrate deep CO layers and interact directly with parenchymal CNS cells to study neural-immune interactions. To evaluate this, we used a robust activation protocol (PMA/ionomycin plus LPS for 4 hours) that simultaneously stimulates all immune cell types within the PBMC population. While this uniform activation is supra-physiological, it is widely used to model human immune-mediated pathologies. Crucially, introducing a 72-hour post-activation delay and washing step prior to co-culture (i.e., donor BB028) yielded identical cellular and transcriptomic alterations. This confirms that the CNS cells were not reacting directly to residual PMA/ionomycin or LPS, but rather to infiltration by the activated autologous immune cells. Nonetheless, these co-culture experiments and the hypothesis-generating spatial transcriptomics data are intended to demonstrate model feasibility and should not be over-interpreted as mechanistic or disease-specific conclusions. Future studies must tailor the immune activation signals to replicate the specific etiology of the disease under investigation.

Because this study focused on model development and optimization, we prioritized donors with sufficient cryopreserved PBMC volumes to complete the full experimental matrix. Although we generated and evaluated over 300 organoids across conditions, the limited donor cohort size precludes definitive disease-specific conclusions. Instead, the high reproducibility of the age-related changes and autologous neuroimmune interactions across all four donors highlights generalizable biological mechanisms.

In conclusion, despite these inherent constraints, myelinated human hiPSC-derived COs successfully recapitulate critical features of CNS aging and offer a powerful platform for investigating human-specific neuroimmune interactions. While animal models provide indispensable systemic *in vivo* architecture, they often fail to replicate human-specific immunological responses and cell-type-specific vulnerabilities. Consequently, these human-derived organoid systems serve as a highly complementary *in vitro* counterpart to *in vivo* animal modeling, bridging a critical translational gap to advance our understanding of human neurodegenerative and neuroinflammatory diseases.

## METHODS

### Human induced pluripotent stem cells (hiPSCs)

The study was approved by the NIH Institutional Review Board. Written informed consent was obtained from the study participants. In this study, we used human induced pluripotent stem cells (hiPSCs) reprogrammed from PBMC obtained from two MS patients (BB007 & BB011) and two healthy controls (BB026 & BB028) who are a part of the natural history protocol “Comprehensive Multimodal Analysis of Neuroimmunological Diseases of the Central Nervous System” (Clinicaltrials.gov identifier NCT00794352) conducted by the Neuroimmunological Diseases Section (NDS) at NIH/NIAID. The hiPSCs were generated at the New York Stem Cell Foundation (NYSCF) under institutional review board-approved protocols. All hiPSC lines were generated using the NYSCF Global Stem Cell Array®, a fully automated reprogramming process that minimizes line-to-line variability.^30^ Each hiPSC line is accompanied by a Certificate of Analysis (CoA), which includes the results of tests for sterility, mycoplasma, karyotyping, identity, and pluripotency.

### Culturing of human induced pluripotent stem cells

After thawing, the hiPSCs were expanded on Matrigel-coated dishes in mTeSR1 medium (StemCell Technologies, #85850) supplemented with 10 μM ROCK Inhibitor (StemCell Technologies, #72304) for the first 24 hours. The hiPSCs were routinely passaged using ReLeSR™ Passaging Reagent (StemCell Technologies, #100-0483) and then cultured on Matrigel-coated dishes in mTeSR1 medium without ROCK inhibitor. Upon proper confluency and morphology, hiPSCs were dissociated into single cell suspension using Accutase (StemCell Technology, #07920), counted and reseeded at the desired density to generate organoids.

### Generation of human cerebral organoids containing myelinating oligodendrocytes

Based on prior protocol established by Lancaster et al.^1,22^, cerebral organoids were generated using the STEMdiff Cerebral Organoid Kit (StemCell Technologies, #8570) from four donors (BB007, BB011, BB026 and BB028). Approximately 90 cerebral organoids were generated from each donor. The manufacturer’s protocol was modified to promote the generation of myelinating oligodendrocytes. Briefly, hiPSCs were dissociated into single cell suspension with Accutase and plated in a 96-well U-bottom plate in Organoid Formation Medium to induce embryoid body formation and germ layer differentiation (Day 0–5), at a density of 2,500 cells/well or 5,000 cells/well. On day 5, the medium was changed to Induction Medium to promote the induction of the neural ectoderm. On day 7, the organoids were embedded in Matrigel droplets (Corning, #354234) and cultured in Expansion Medium to stimulate the growth of neuroepithelial buds. The organoids were then transferred to Maturation Medium and placed on the orbital shaker in the 5% CO2 incubator at 37°C for maturation on day 10.

To pattern oligodendrocyte progenitor cells (OPCs) and differentiate them into myelinating oligodendrocytes (OG), a prior protocol from Madhaven et al.^3^ was adopted. Starting on day 50, organoids were stimulated with 10 ng/mL PDGF-AA (R&D Systems, #221-AA-050) and 10 ng/mL IGF-1 (R&D Systems, #291-G1–200) for 10 days, followed by treatment with 40 ng/mL 3,3’,5-triiodothyronine (T3, Sigma, #ST2877) for next 10 days. PDGF-AA and IGF-1 are potent mitogens that promote the proliferation and survival of OPC, while T3 mediates maturation of OPCs to mature oligodendrocytes throughout nuclear receptors. From this point, the organoids were maintained in Maturation Medium (50% [v/v] DMEM/F12 and 50% [v/v] Neurobasal medium containing 1X N2 supplement, 250 μL/L insulin, 1xGlutaMAX, 0.5xMEM-NEAA, 1% [v/v] penicillin-streptomycin, 0.4 mM ascorbic acid, 7 μL/L β-mercaptoethanol, 1xB27 supplement with vitamin A and 1g/L of sodium bicarbonate^1,22^) with every-other-day media changes until the completion of the experiment.

### Microglia progenitors’ differentiation and their co-culture with cerebral organoids

In parallel, hiPSCs were differentiated from three donors (BB011, BB026 & BB028) into microglial progenitors following the MIGRATE protocol from Fattorelli et al.^23^ Differentiation was initiated using mTeSR medium supplemented with 50 ng/mL human BMP4, 50 ng/mL human VEGF, and 20 ng/mL human SCF. From days 4 to 10, cells were stimulated with 50 ng/mL human SCF, 50 ng/mL human M-CSF, 50 ng/mL human IL-3, 50 ng/mL human FLT3, and 5 ng/mL human TPO in complete X-VIVO 15 medium (2 mM GlutaMAX, 100 U/mL Antibiotic-Antimycotic, and 0.055 mM 2-mercaptoethanol). On day 11, stimulatory agents in the media were adjusted to 50 ng/mL human FLT3 and 50 ng/mL human M-CSF, with the addition of 25 ng/mL human GM-CSF until day 18. Double positive, CD14 and CX3CR microglial progenitor cells were identified via FlowCytometry and co-cultured with 30 days cerebral organoids at a density of 2×10^5^cells/organoid. Organoids were later harvested for live imaging or immunohistochemistry.

### Generation of human vessel organoids

Adopting protocol by Sun et al.^20^ to generate human blood vessel organoids, hiPSCs were resuspended in mTeSR1 medium with 10 μM Y27632 and seeded at a density of 9,000 cells per well in a lipidure coated V bottom 96 well plate. On day 2, the aggregates were cultured in mesodermal induction medium (APEL2 (StemCell Technologies, #5270) supplemented with 6 μM CHIR9902). Media was replaced on day 4 with endothelial induction medium composed of APEL2 supplemented with 50 ng/mL VEGF, 25 ng/mL BMP4, and 10 ng/mL bFGF. From days 7 to 11, the medium was replenished with MV2 medium (PromoCell, #C-22022) containing 50 ng/mL VEGF to promote endothelial cell maturation. On day 12, the final day of differentiation, vessel organoids were embedded in Matrigel and cultured in Maturation Medium supplemented with 20 ng/mL VEGF.

### Fusion of cerebral and vessel organoids

To generate the fused cerebral organoids, 7-day-old cerebral organoid (BB007) and 12-day-old vessel organoid were embedded together in Matrigel droplet.^20^ The fused organoids were maintained on a shaker at 85 rpm maintained in Maturation Medium supplemented with 20 ng/mL VEGF. Approximately 60 assembloids were generated across three independent fusion experiments. One or two representative assembloids from each batch were selected for immunofluorescence staining to confirm successful fusion.

### Co-culture of cerebral organoids with autologous PBMC

To further characterize neural-immune interactions, autologous peripheral blood mononuclear cells (PBMC) were co-cultured with cerebral organoids. PBMC were thawed in RPMI1640 medium (Gibco, # 11875085) supplemented with 1% [v/v] penicillin-streptomycin and 10% [v/v] FBS. Cells were rested for 1 hour at 37 °C in 5% CO2 prior to 10 ng/mL PMA, 1 µg/mL ionomycin, and 1 µg/mL LPS stimulation for 4 h. The activated PBMC were either used directly for co-culture experiment (donors BB007, BB011, BB026) or allowed to rest for 72 hours before co-culturing them with organoid (donor BB028). Duration of stimulation was determined based on preliminary experiment. Dead cells and debris were removed using the Dead Cell Removal Kit (Miltenyi Biotec, #130-090-101) to achieve a final viability of 98–100%. Resting or activated PBMC were co-cultured at 2×10^6^ cells per organoid in Maturation Medium for 24 or 72 hours.

Next, the organoids were used for downstream experiments which included live imaging (BB011, BB026, BB028), multiplexed immunofluorescence imaging (BB011, BB026, BB028) and Xenium Spatial transcriptomics (BB007, BB026 and BB028).

### Cytokine measurements

Conditioned media were collected after 24 h from cerebral organoids cultured alone, cerebral organoids co-cultured with activated autologous PBMC, and activated PBMC cultured alone. Samples were centrifuged to remove cellular debris (250xg, 5 min, RT), aliquoted, and stored at −80°C until analysis. Concentrations of GM-CSF, IL-1β, IL-6, IL-10, TNF-α, and VEGF-A were quantified using the U-PLEX Custom Biomarker (hu) kit (#K15067M-2, Meso Scale Discovery, Rockville, MD, USA) according to the manufacturer’s instructions. IFN-γ concentrations were measured separately using the U-PLEX Human IFN-gamma Assay (#K151TTK-2, Meso Scale Discovery) following the manufacturer’s protocol. Prior to analysis, all samples were diluted 1:2 with the appropriate MSD assay diluent. Assays were read on the MESO QuickPlex SQ 120 instrument (Meso Scale Discovery), and cytokine concentrations were calculated using Discovery Workbench software based on the corresponding standard curves and expressed in pg/mL. The measured concentrations are presented after correction for the 1:2 sample dilution.

### Live imaging of cerebral organoids co-cultured with immune cells

Live imaging was performed on cerebral organoids co-cultured with either hiPSC-derived microglial progenitors or autologous PBMC (resting or activated for 4 hours). Briefly, cerebral organoids were incubated in Maturation Medium with 0.1 µM Red Cell Tracker (C34552, ThemoFisher) for 1 hour, then washed three times to remove excess dye. In parallel, immune cells were stained with 0.5 µM Green Tracker (C7025, ThemoFisher) for 15 minutes in RPMI medium containing 10% FBS, washed three times with complete medium, and resuspended in Maturation Medium supplemented with 20% FBS. hiPSC-derived microglial progenitors (2 x 10^5^ cells/organoid) or autologous PBMC (2 x 10^6^ cells/organoid) were incubated for 1-hour with organoid in the 5% CO2 incubator at 37°C. Post-incubation, organoids were transferred to 14 mm glass-bottom microwell dishes (MatTek). A preheated solution of 2% agarose in Maturation Medium was cooled to 37°C and immediately poured over the organoid and immune cells. Upon agarose polymerization, complete Maturation Medium supplemented with 20% FBS was added to the dishes. The co-cultures were incubated at 37°C in a humidified environment for an additional two hours. Imaging was performed over 24-hour period using a Leica SP8 DIVE (Deep In Vivo Explorer) inverted confocal microscope, which featured five channels—four HyD detectors and one PMT fusion DIVE detector—as well as a motorized stage and a 37°C incubation chamber provided by the NIH Division of Scientific Equipment and Instrumentation Services. The microscope was configured for four-dimensional analysis (x, y, z, and time) to monitor cell segregation and migration. Excitation was achieved with a 405 nm diode laser, a 488 nm Argon laser, a 561 nm DPSS laser, and HeNe lasers for 594 nm and 633 nm, all set to minimal power (0.05–1%). Z-stack images covering 100–250 µm were captured over time. Additionally, mosaic images of the entire organoids were created by acquiring multiple Z-stacks in Navigator mode to cover the full structure, and these were assembled into a tiled image using LAS X software. Final image processing was carried out with both the Leica Application Suite (LAS X) and Imaris (Bitplane).

### Immunofluorescence

For whole-mount staining, vessel organoids and assembloids were fixed in 4% PFA at 4°C overnight. After fixation, they were washed three times with PBS and then incubated in 0.5% Triton X-100 at RT (room temperature) for 1 hour. Following this, the organoids were blocked with 5% donkey tissue in 0.1% Triton X-100 at RT for 1 hour. Organoids were then incubated with primary antibodies at 4°C for over 72 hours, followed by washing with PBS (3x, 10 min). Next, they were incubated with secondary antibodies at 4°C for another 72 hours. The stained organoids were washed three more times with PBS before being subjected to confocal imaging. Detailed information about the primary and secondary antibodies can be found in Key Resources Table. The images were acquired using a Leica SP8 WLL FLIM microscope, and then analyzed using Imaris software for processing and analysis.

### Multiplexed immunofluorescence imaging

Large-scale highly multiplexed immunofluorescence imaging was performed as previously described by Maric et al.^31^ at NINDS Flow Cytometry and Imaging Core Facility (FCICF).

Organoids were fixed in 4% paraformaldehyde (PFA) at 4°C overnight. Next, they were dehydrated sequentially - first in a 20% sucrose solution and then in a 30% sucrose solution, with each step performed overnight. Finally, the organoids were embedded in optimal cutting temperature (OCT) medium and sectioned into 10-μm-thick slices using a Leica cryostat.

Cryosections were retrieved from −80 °C storage and allowed to thaw at RT. The sections were then air-dried overnight to ensure robust adhesion to the microscope slides. All subsequent procedures were performed at RT.

The slides were initially permeabilized by sequential immersion in graded methanol solutions (70%, 80%, and 95%; 2 minutes per step), followed by rehydration and rinsing in distilled water (dH₂O). Antigen retrieval was carried out by completely submerging the sections in 10 mM sodium citrate buffer (pH 6.0) and applying a 2-minute heat-mediated treatment in an 800W microwave (GE model PEM31DFWW) operating at 100% power.

To minimize non-specific binding, sections were incubated for 15 minutes at RT in neat FcR Blocking solution (Innovex Biosciences, NB309) to block endogenous Fc receptors, followed by treatment with Background Buster solution (Innovex Biosciences, NB306).

Primary immunoreactions were performed by iteratively incubating the sections using different cocktails of up to 8 unconjugated, immunocompatible primary antibodies, each used at a final concentration of 1 μg/mL (i.e., a 1:1,000 dilution of a 1 mg/mL stock) in PBS supplemented with 1 mg/mL BSA, for 60 minutes at RT. Following primary antibody incubation, the sections were washed three times in PBS and three times in dH₂O (1 minute per wash). Appropriate spectrally compatible secondary antibodies, each at a final concentration of 1 μg/mL, were then applied. Detailed information about the primary and secondary antibodies can be found in Key Resources Table. After a second series of washes (3× in PBS and 3× in dH₂O), the sections were counterstained with 1 μg/mL DAPI (Thermo-Fisher Scientific) to provide a reference channel for autofocus and pixel-to-pixel registration during subsequent image analyses. Iterative Multiplexing for High-Plex Immunohistochemistry

After completing the first round of up to 10-plex immunostaining and imaging, the bound primary and secondary antibodies were removed by incubating the slides in NewBlot Nitro 5X Stripping buffer (Li-Cor Biosciences) for 5 minutes at RT. This was followed by a 1-minute heat-mediated antigen retrieval in 10 mM Sodium Citrate buffer (pH 6.0) using the same 800W microwave set at 100% power. Each re-staining cycle, beginning with tissue re-blocking using FcR Blocking and Background Buster solutions, was then repeated with a new cocktail panel of primary and secondary antibodies.

Following staining, the sections were cover-slipped using Immu-Mount medium (Thermo-Fisher Scientific, MI). Fluorescence imaging was performed using an Axio Imager.Z2 10-channel scanning microscope (Carl Zeiss, Thornwood, NY) equipped with a ×20, 0.8 NA Plan-Apochromat (Phase-2) non-immersion objective, a 16-bit ORCA-Flash 4.0 sCMOS digital camera (Hamamatsu Photonics, Japan), and a 200W X-Cite 200DC broadspectrum light excitation source (Lumen Dynamics). Ten self-contained excitation/dichroic/emission filter sets (Semrock, Rochester, NY) optimized for the detection of up to 10 spectrally compatible fluorophore combinations (e.g., DAPI, DyLight 405, Alexa Fluor 430, Alexa Fluor 488, Alexa Fluor 546, Alexa Flor 594, Alexa Fluor 647, PerCP, IRDye 680LT and IRDye 800CW) were used for imaging per each slide scanning round, as previously described (Maric et al., 2021). Each labeling reaction was captured sequentially in separate image channels using the ZEN 2 image acquisition software. The acquired 16-bit images (600 × 600 μm tiled fields with 10% overlap) were globally stitched in ZEN 2 and then sequentially processed for (1) pixel-to-pixel registration for all imaging channels acquired across multiple rounds of antibody staining and tissue scanning, (2) illumination correction for vignetting artifacts in each imaging channel, (3) autofluorescence subtraction, and (4) spectral unmixing for any signal bleedthrough in spectrally overlapping channels using an open source python pipeline (https://github.com/RoysamLab/whole_brain_analysis.git) previously published (Maric et al., 2021). Finally, pseudo-colored images were converted to 8-bit BigTIFF files and merged in Adobe Photoshop to generate multi-colored composite images. The unprocessed images were utilized to create the figures with all panels included. Subsequently, the curves tool was applied for linear adjustments across the entire image. Unprocessed images will be provided by the lead contact upon request.

### Immunofluorescence image quantification

Immunofluorescence images were analyzed using FIJI (ImageJ, NIH). The organoid area was defined as the region of interest (ROI). For each marker, a consistent threshold was applied to all images within the same experiment to identify positively stained regions. The marker-positive area was quantified as the percentage of the total ROI area (% area fraction). Identical image acquisition settings and image analysis parameters were used for all samples within each experiment.

### Single Cell RNA Sequencing

Sixty-day-old assembloids (n=2) were dissociated into single-cell suspensions using a modified Miltenyi Neural Dissociation Kit protocol. Assembloids were transferred to a 6-well plate containing pre-warmed (37 °C) Enzyme Mix 1. Samples were incubated at 37 °C on a shaker (85 rpm) for 15 minutes, gently triturated 10 times using a wide-bore pipette, and then incubated for an additional 15 minutes under the same conditions. Enzyme Mix 2 was then added, followed by the same titration and two 10-minute incubations. Dissociated cells were centrifuged at 250 × g for 5 minutes at 4 °C, resuspended in 1 mL HBSS+, and filtered through a pre-wet 70 µm MACS Smart Strainer. The strainer was rinsed with 5 mL HBSS+, and cells were centrifuged again at 250 × g for 10 minutes at room temperature. Finally, cells were resuspended in Fluent Biosciences’ cell suspension buffer containing 40 units of RNase inhibitor, kept on ice, and immediately processed for capture and lysis.

Cell suspension was gently mixed using a wide-bore pipette and 40,000 cells were transferred into the provided PIPs along with 40 units of RNase Inhibitor. The PIPseq v4.0PLUS T20 3’ Single Cell RNA Kit follows a standardized workflow; therefore, all subsequent processing adhered to the manufacturer’s protocol (Doc ID: FB000213-, Revision 8.9). Prior to library preparation, cDNA fragments were assessed for quality using the Agilent TapeStation and quantified with the Qubit High Sensitivity Kit. After quality assessment, subsequent library preparation was processed as recommended by manufacturer.

Prepared duplicate libraries were pooled and sequenced on a NovaSeq X Plus. All samples have sequencing yields of more than 604 million read per sample. The sequencing run was set up as a 54-cycle + 68-cycle asymmetrical run. Demultiplexing was performed using CellRanger (bcl2fastq 2.20), allowing one mismatch.

### Xenium spatial transcriptomics

For Xenium spatial transcriptomics, PBMC were treated as described in the “Co-culture of cerebral organoids with autologous PBMC” section. Briefly, autologous PBMC were stimulated with PMA/Ionomycin and LPS for 4 h. The activated PBMC were either used directly for co-culture experiment (donor BB026 and BB007) or allowed to rest for 72 hours (donor BB028) before co-culturing them with organoid. Resting PBMC were thawed on the day of co-culture and rested for 1-hour. Then, resting or activated PBMC were co-cocultured with cerebral organoid in Maturation Medium for 24 h. For donors BB026 and BB028 the following conditions were included: 1. cerebral organoid alone, 2. cerebral organoid co-cultured with resting PBMC, 3a. cerebral organoid co-cultured with activated PBMC, 3b. cerebral organoid co-cultured with activated PBMC. Note, there were two organoids/donor co-cultured with activated PBMC, for 4 organoids/donor total. To test reproducibility of findings across biological replicates, for donor BB007, we included 4 separate organoids/condition studying only conditions: 1. cerebral organoid alone, and 3. cerebral organoid co-cultured with activated PBMC, resulting in a total of 8 organoids/donor.) Together with the eight organoids from the aged donor experiment, a total of 16 organoids were analyzed by Xenium spatial transcriptomics. After 24 hours of co-culture, the organoids were fixed in 4% PFA at 4°C overnight and used for downstream spatial transcriptomics.

Tissue sample preparation followed the manufacturer’s protocol (“Xenium In Situ Protocols FFPE_Tissue Preparation Guide - CG000578 Rev E”). Five micrometer sections were prepared under RNase-free conditions according to the protocol, mounted on Xenium slides with a 235 mm² positioning area (10.45 mm x 22.45 mm), and stored in slide mailers with desiccant at room temperature. For quality control, subsequent sections were stained with Hematoxylin and Eosin (H&E) as recommended by the manufacturer. On the following day, the FFPE tissue sections were deparaffinized and decrosslinked according to the “Xenium In Situ for FFPE - Deparaffinization & Decrosslinking - CG000580 Rev A” protocol. The slides were then processed immediately following the Xenium Prime Expression with Optional Segmentation User Guide (CG000760 Rev B) using the Xenium Prime 5K Human Pan Tissue and Pathways Assay Kit (10x Genomics, CA, USA; 1000671) along with predesigned Panel (10x Genomics, CA, USA; PN1000766), which targets 100 genes (see Supplementary Table 1).

Briefly, primer hybridization was performed first, followed by RNase treatment and polishing, and overnight probe hybridization. The next day, the samples underwent post-hybridization washing and enzymatic amplification to generate multiple copies of the gene-specific barcode for each RNA target. Finally, cell segmentation and DAPI staining were performed, and data acquisition was performed with the Xenium Analyzer (the instrument software version 3.1.0.0 with analysis version 3.1.0.4.; 10x Genomics, CA, USA; the protocol for the UserGuide: CG000584 Rev E), yielding cell-by-gene and transcript-by-location matrices.

### Processing and analysis of scRNA-seq data

FASTQ file processing: FASTQ files were processed with PIPseeker 3.3.0 software (Fluent BioSciences) using the default parameters. To accommodate for differences between samples and facilitate accurate cell calling, PIPseeker performs cell calling at five different sensitivity levels. With sensitivity_3, the number of cells captured ranges from 11,013 to 12,160 and mean reads per cell ranges from 75,714 to 111,141. Median genes found per cell ranges from 1,846 to 1,937 and the total number of genes detected ranges from 32,180 to 32,600

For single-cell RNA sequencing dataset, quality control was performed by examining the distributions of the total number of unique molecular identifiers (nCount_RNA), the number of detected genes (nFeature_RNA), and the percentage of mitochondrial transcripts (percent.mt) using scatter and violin plots. Sample-specific thresholds were selected to remove low-quality cells, low-complexity cells, putative doublets or multiplets, and stressed or dying cells. The cells were retained if they met the following criteria: nCount_RNA > 800, 500 < nFeature_RNA < 5,000, and percent.mt < 10%. For PBMC samples, all datasets were first jointly visualized to determine a conservative set of filtering thresholds that could be applied consistently across samples.

Cell clustering and annotation: PIPseeker “sensitivity=3” count matrices were used to input into Seurat V4.4.0 (https://doi.org/10.1038/s41587-023-01767-y). Comparable libraries were integrated via CCA using 30 dimensions at default resolution for clustering. Doublets were removed using DoubletFinder package at random. Clusters were manually annotated using canonical markers from Seurat’s’ “FindMarkers” to compute Wilcoxon-based tests with default parameters to generate differential expression.

### Processing and analysis of Xenium spatial tanscriptomic data

The initial processing of Xenium data was done with Xenium Onboard Analyzer (XOA) v3.1.0.4 (10X Genomics, Pleasanton, CA) utilizing the following settings: preparation method (ffpe), cell segmentation (Xenium multi-tissue stain), panel name (hMulti_100g), panel designer version (3.2.0), predesigned panel ID (hAtlas_v1.1), tissue type (human multi), chemistry version (Xenium Prime), number of predesigned target genes (5001) and number of custom target genes (100). As part of the downstream Xenium data analysis, XOA outputs derived from each sample region were loaded into R package Seurat (v5.2.1)^32^ using the LoadXenium function and filtered by the subset function (nCount_Xenium > 10).

Quality control: Genes expressed in fewer than 3 cells and cells expressing fewer than 5 genes were excluded. Cells with ≤10 total transcripts were removed as low-quality. After quality control filtering the 3-donor merged CNS dataset (BB026, BB028 and BB007 combined) comprised 580,144 high-quality cells with 175,472,763 total transcripts, with median transcripts/cell 76 and mean transcripts/cell 302.46. Median genes/cell 67, mean genes/cell 185.86. The d750 organoids had lower median numbers of per cell transcripts/genes, comparable between donors (transcripts/cell: median 41 and 37, mean 319.08 and 323.62; genes/cell median 35 and 31, mean 183.24 and 175.37) compared to d200 organoids (transcripts/cell: median 93, mean 294.14; genes/cell median 81, mean 188.54).

Normalization and integration: All 16 samples were merged into a single Seurat v5 object. Normalization was performed using SCTransform. Principal component analysis (PCA) was computed on the top 50 components. To correct for donor-level batch effects across 3 donors and 2 experiments, Harmony integration was applied on all 50 PCA dimensions, grouping by donor. Uniform Manifold Approximation and Projection (UMAP) was computed from the first 30 Harmony-corrected dimensions.

Unsupervised clustering: Graph-based clustering was performed using shared nearest neighbor (SNN) graphs and the Louvain algorithm at multiple resolutions (0.1–1.5) to assess cluster stability. The final clustering resolution was selected based on biological interpretability: resolution 0.5 was used for the initial full dataset, resolution 0.2 for the CNS subset.

Immune versus CNS cell classification: CNS cells have extensive cellular processes (e.g., axons/dendrites) that contain lineage-specific transcripts (e.g., GFAP in reactive astrocytes, MBP/PLP1 in oligodendrocytes, MAP2, NEFL, NEFM, NEFH in axons) that overlap and intertwine into dense network (e.g., neuropil), contaminating overlapping cells. Because organoid-incorporated immune cells represent a small minority of the total cell population and do not form discrete clusters at standard resolutions, we employed a per-cell marker-based classification strategy rather than cluster-level annotation. For each cell, we computed expression scores from raw counts for the following immune lineage markers: T cells (CD3E, CD3G, plus cytotoxic markers GZMA, GZMB, GZMK, GZMH, PRF1), monocytes (CD14, CD68, CSF1R), and B cells (CD19, MS4A1, CD79A). Simultaneously, we computed a CNS marker score (SOX2, VIM, GFAP, MAP2, TUBB3, MBP, OLIG1, OLIG2). A cell was classified as immune if it met all of the following criteria: (1) expression of at least 2 T cell markers (CD3E/CD3G or cytotoxic markers) OR at least 2 monocyte markers OR at least 2 B cell markers; (2) CNS marker score ≤3 total counts; and (3) the cell originated from a co-culture condition (not CO alone, where no PBMC were added). Cells in CO alone samples were assigned to the CNS compartment regardless of marker expression, as no immune cells were present in these conditions. Immune cells identified by the per-cell classification were subset and independently processed: SCTransform normalization, PCA (30 components), Harmony integration by donor (30 dimensions used for PCA, 20 for UMAP), and Louvain clustering at resolution 0.3. Ten immune cell clusters were annotated based on known lineage markers (Figure S22).

CNS cell type annotation: CNS clusters were annotated into 7 lineages based on manual examination of: average expression z-scores for canonical markers, FindAllMarkers top differentially expressed genes per cluster, percent expression of key lineage markers, and QC metrics (nCount, nFeature) per cluster. CNS cells were annotated using established marker gene panels including progenitor markers (SOX2, NES, PAX6), glial markers (AQP4, OLIG1/2, PDGFRA, MOG, CNP), neuronal markers (SNAP25, NEFL/M/H, SLC17A7, GAD1/2). Process-localized transcripts (GFAP, GJA1, MBP, PLP1) were excluded from the scoring as these genes are transported to cellular processes and their spatial detection in Xenium does not reliably reflect cell body identity. Module scores were calculated per cell using Seurat’s AddModuleScore function, then z-score normalized across clusters to enable fair comparison. Each cluster was assigned the cell type identity corresponding to its highest z-scored module score. Annotations were validated using FindAllMarkers (Wilcoxon rank-sum test, min.pct = 0.2, log₂FC threshold = 0.3, positive markers only). We then merged clusters that mapped to seven major CNS lineage (Figure 6) to avoid un-interpretable subpopulations and rather compare phenotypical changes between young and old organoids using differential gene expression analyses. The seven CNS cell lineages were: 1. Astrocytes (clusters 1, 3, 7, 8, 12, 15, 19, 22, 23): defined by GFAP, AQP4, NDRG2, GJA1, and/or SOX9 expression. Included metabolically active subpopulations (high IGFBP2); 2. Reactive astrocytes (clusters 2, 13, 14, 16, 18, 21): astrocyte background (GFAP+) with superimposed interferon-stimulated gene signature (ICAM1, GBP1, IRF1, CXCL10) and/or NF-κB-driven inflammatory markers (CCL2). These represent organoid astrocytes responding to activated PBMC co-culture. 3. Radial glia (clusters 4, 9, 10, 17, 24, 25, 26): dominated by SOX2-OT expression (64-100% of cells). 4. Mature neuron (cluster 5): STMN2 (63%), DCX (34%), SLC17A7 (24%), SYP (20%). 5. Progenitor/OPC (cluster 6): FOXG1 (19%), DCX (19%), STMN2 (13%), representing cells in an early progenitor state. A dedicated OPC cluster could not be resolved (only 287 cells [0.05%] in the entire dataset co-expressed PDGFRA and OLIG2 and they mostly localized to cluster 6) therefore progenitor/OPC cells were merged. 6. Oligodendrocyte (cluster 11): PLP1 (47%), MBP (22%), CNP (16%). Note: GFAP, MBP, and PLP1 transcripts detected in processes/extracellular space were not used for cell body classification, as these represent transcript localization in cellular projections rather than perisomatic expression. 7. Choroid plexus (cluster 20): FOLR1 (37%), IGFBP2 (55%).

### Statistical analysis

Differential gene expression analysis: Differentially expressed genes (DEGs) between CNS cell types were identified using FindAllMarkers in Seurat with the Wilcoxon rank-sum test (min.pct = 0.25, log₂FC threshold = 0.5, only positive markers, downsampled to a maximum of 5,000 cells per identity). For condition-specific DEGs within CNS cell types, FindMarkers was applied per donor comparing organoid-alone vs. co-culture conditions (min.pct = 0.1, log₂FC threshold = 0.1), requiring a minimum of 10 cells per group. A gene was considered significant at adjusted p-value < 0.05 (Bonferroni correction). To identify robust transcriptional changes reproducible across donors, a congruence analysis was performed: genes were retained only if they were significant (adjusted p < 0.05) in both donors (BB026 and BB028) with the same direction of effect. The reported fold change for congruent genes was the mean log₂FC across both donors.

DEGs between young and old CNS lineages at baseline: To identify transcriptomic differences between young (d200, BB007) and old (d750, BB026+BB028) organoids in the absence of immune stimulation, we performed differential expression analysis within each CNS lineage using cells from the CO alone condition only. For each of the 7 CNS lineages (Astrocyte, Reactive_Astrocyte, Radial_Glia, Mature_Neuron, Progenitors/OPCs, Oligodendrocyte, Choroid_Plexus), Seurat’s FindMarkers was applied comparing Young versus Old cells using the Wilcoxon rank-sum test on SCT-normalized data. Genes with adjusted p-value (Benjamini-Hochberg) < 0.05 were considered significant. BB007_R5 (flagged as an incomplete region) was excluded from all analyses.

Pseudotime analysis of astrocytes: To infer the transcriptional trajectory from resting astrocytes toward a reactive astrocyte state and compare its distribution between young and old organoids, we performed pseudotime analysis using Slingshot (v2.18.0). We subset the 3-donor annotated CNS object to cells classified as Astrocyte or Reactive_Astrocyte in the CO alone condition only (unstimulated). To ensure balanced representation across donors while maintaining computational tractability, cells were downsampled to a maximum of ∼16,666 cells per donor (50,000 total). Data were log-normalized, 2,000 variable features were identified (Seurat FindVariableFeatures), and PCA was computed (30 components). UMAP was generated from the first 20 PCs for visualization. Slingshot was run on the first 20 PCA dimensions using the lineage annotation (Astrocyte, Reactive_Astrocyte) as cluster labels, with Astrocyte set as the starting cluster and Reactive_Astrocyte as the end cluster. This produced a single trajectory representing the activation continuum. Pseudotime values were scaled to [0, 1] for cross-group comparability. The pseudotime distributions of Young (d200, BB007) and Old (d750, BB026+BB028) astrocytes were compared using the Kolmogorov-Smirnov test and Wilcoxon rank-sum test. Gene expression dynamics along pseudotime were visualized using loess smoothing (span=0.4) of log-normalized expression, stratified by age group.

Senescence marker analysis: To assess the senescence state of CNS lineages within cerebral organoids, we curated a panel of senescence-associated genes organized into functional categories: CDK inhibitors (CDKN1A, CDKN2A, CDKN2B, CDKN1B), DNA damage response/tumor suppressors (TP53, RB1, ATM, ATR, H2AFX, MDM2), lysosomal markers associated with SA-beta-galactosidase activity (GLB1, LAMP1, LAMP2, SQSTM1, CTSL), anti-apoptotic factors (BCL2, BCL2L1), stress response genes (GADD45A, HMOX1, SOD1), senescence-associated secretory phenotype (SASP) factors (CCL2, CXCL10, CXCL12, IL6, VEGFA, SERPINE1, IGFBP2, GDF15), and chromatin/nuclear lamina regulators (LMNB1, SIRT1, SIRT2, HDAC1, EZH2, HMGA1). For each gene and cell type, we computed the percentage of cells expressing the marker (detection rate) and mean normalized expression from the Xenium counts data. An extended panel of 50 senescence-associated genes with detection rate >=0.5% was also assessed. Results were visualized using DotPlots (Seurat) and FeaturePlots on UMAP embeddings.

Age-differential transcriptomic responses to activated PBMC: To identify genes whose response to activated PBMC co-culture differs between young and old organoids (i.e., age-by-treatment interaction effects), we implemented a two-step interaction analysis within each CNS lineage.

Step 1: Per-age-group response. For each lineage, we computed the transcriptomic response to activated PBMC separately in Old organoids (BB026+BB028 pooled) and Young organoids (BB007) using Seurat’s FindMarkers (Wilcoxon rank-sum test, SCT assay, min.pct=0.05, logfc.threshold=0). This yielded a response log2 fold-change for each gene in each age group.

Step 2: Interaction contrast. To formally test whether the age effect differed between stimulated and unstimulated conditions, we computed for each gene: (a) the Old-vs-Young difference under Activated PBMC stimulation (FindMarkers: Old_ActPBMC vs Young_ActPBMC), and (b) the Old-vs-Young difference at baseline (FindMarkers: Old_CO vs Young_CO). The interaction fold-change was defined as the difference between these two age effects: interaction_FC = FC(age_in_ActPBMC) - FC(age_in_CO).

Genes were classified as age-differential if they had a significant age effect under stimulation (adjusted p-value < 0.05, Benjamini-Hochberg correction) and an absolute interaction fold-change > 0.5. Genes with positive interaction FC were annotated as “Stronger in Old” (greater age-dependent change under stimulation), while genes with negative interaction FC were annotated as “Stronger in Young.” Results were visualized as scatter plots comparing the response magnitude (log2FC of ActPBMC vs CO) in Young (x-axis) versus Old (y-axis) for each lineage, with age-differential genes highlighted.

Detection of linear MBP structures: Putative myelinated axon segments were detected using extracellular MBP and PLP1 transcript localization. MBP and PLP1 transcripts not assigned to any cell (cell_id = “UNASSIGNED” in Xenium transcript output) were extracted from the transcripts.parquet files. Spatially proximal transcripts were grouped using DBSCAN clustering (epsilon = 5 μm, minimum points = 4). For each cluster, linearity was assessed by PCA of the transcript coordinates: structures with an eigenvalue ratio (PC1/PC2) ≥ 4 and a length (range along PC1) ≥ 10 μm were classified as linear structures consistent with oligodendrocyte processes. Structure lengths and counts were compared between conditions using the Wilcoxon rank-sum test.

## DATA AVAILABILITY

- Xenium spatial transcriptomics data have been deposited at GEO GSE293337 and are publicly available as of the date of publication.
- RNAseq data has been deposited at GEO GSE341831 are publicly available as of the date of publication.
- Source data for the main and supplementary figures are provided in Supplemantary Data 2.
- Microscopy data reported in this paper will be shared by the corresponding contact upon request.
- Any additional information required to reanalyze the data reported in this paper is available from the corresponding author upon request.

## CODE AVAILABILITY

- The original code used for the analysis of Xenium spatial transcriptomics data in this study has been deposited at https://github.com/cihangenome/xenium-cerebral-organoids/.
- The updated code for additional analyses of the Xenium spatial transcriptomics data adding second Xenium experiment (donor BB007, d200, 8 organoids and merging 3 donors, re-clustering, DEG analyses, Astrocyte pseudotime, Senescence, myelin transcripts) has been deposited at https://github.com/Bielekova-Lab/Code_CO-NPG_RegMed2026.

## ACKNOWLEDGMENTS

We thank the MS patients, healthy donors, caregivers, and clinical team at the Neuroimmunological Diseases Section for their invaluable support. We also thank the Biological Imaging Section at NIAID for assistance with live imaging; Dr. Katie Williams and Dr. James Carroll (Rocky Mountain Laboratories, NIH) for training in cerebral organoid generation and PIP-seq, respectively; Dr. Valentina Fossati (NYSCF) for guidance on the induction of myelinating oligodendrocytes; and Darwing Padilla for assistance with cerebral organoid maintenance. This work was supported by the Division of Intramural Research, National Institute of Allergy and Infectious Diseases (NIAID), National Institutes of Health (NIH).

## AUTHOR CONTRIBUTIONS

Conceptualization, B.B.; methodology: J.K., P.K., D.M, C.W., C.H, K.P., B.B.; investigation, J.K., S.H.P., D.M., P.K, G.W., B.B.; RNAseq data analysis: CO, TM, JL, J.K., S.H.P, P.K., B.B.; visualization: J.K., S.H.P., P.K., B.B.; writing—original draft: J.K., B.B., writing—review & editing: B.B., J.K., S.H.P, C.H., K.P.; funding acquisition, BB; supervision, B.B., K.P, C.H.; review & scientific editing: all authors.

## COMPETING INTERESTS

All authors declare no financial or non-financial competing interests.

## SUPPLEMENTARY INFORMATION

Document S1. Figures S1-S22 and Tabe S1

**Supplementary Data 1**. List of genes included in the single cell in situ spatial profiling experiment.

**Supplementary Data 2**. Source data for the main and supplementary figures.

**Video S1**. Cerebral organoid co-cultured with microglia progenitors. Organoid (red), DAPI (blue), microglia progenitors (green).

**Video S2.** Cerebral organoid co-cultured with activated PBMC. Organoid (red), DAPI (blue), PBMC (green).

## Notes

### Competing Interest Statement

The authors have declared no competing interest.

### Author Declarations

The study was approved by the NIH Institutional Review Board. Written informed consent was obtained from the study participants.

