## Supplementary material for "DEVELOPMENT OF ORGANOID BASED MODEL TO STUDY IMMUNE-NEURAL INTERACTIONS IN HUMAN CNS DISEASES": Document S1

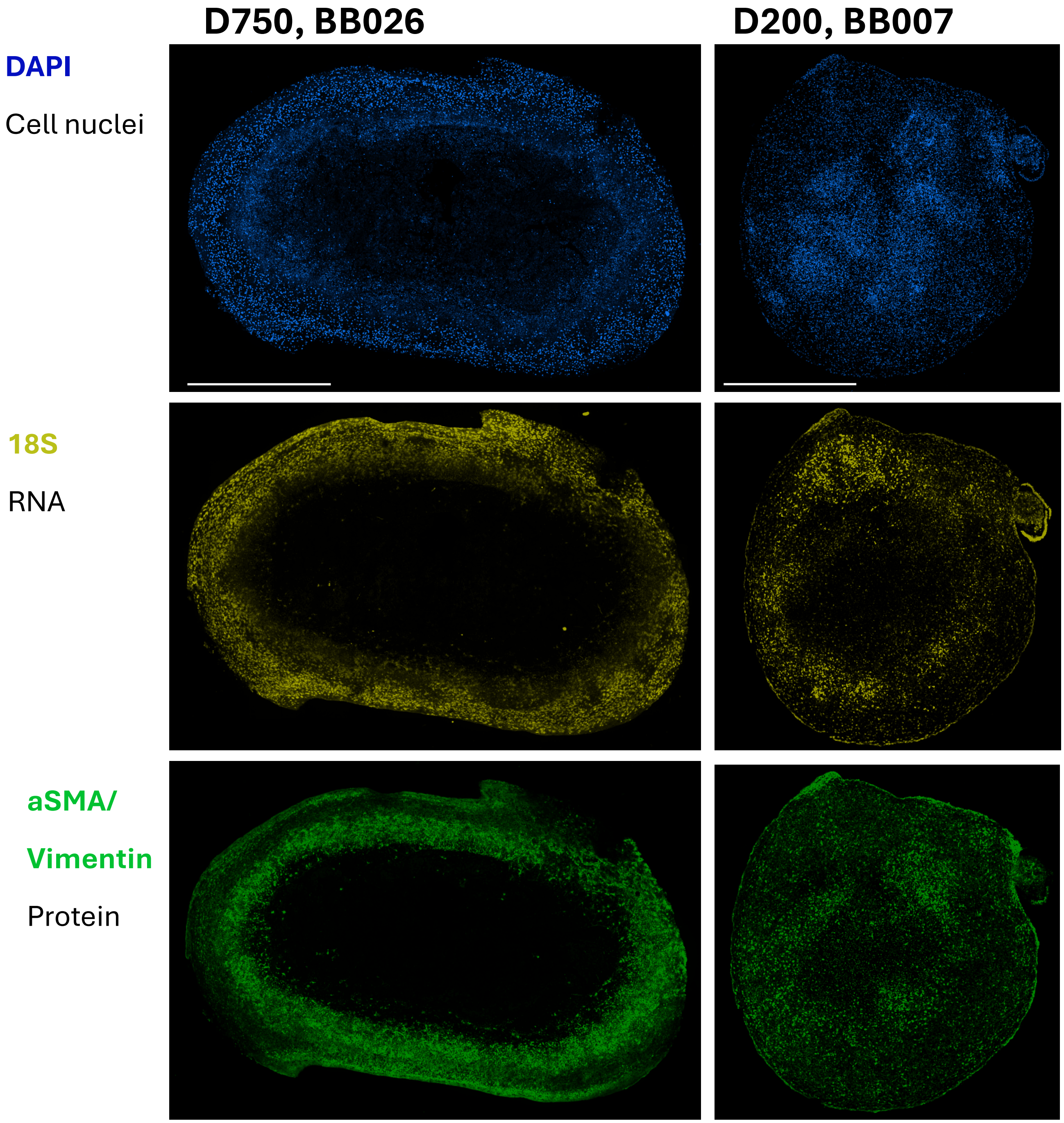


**Figure S1:** Structural changes in young and aged cerebral organoids.

Representative whole-section Xenium morphology images of young (day 200, BB007) and aged (day 750, BB0260) cerebral organoids. Sections were stained with DAPI (blue), the 18S ribosomal RNA probe (yellow), and αSMA/Vimentin morphology stain (green). Compared with young organoids, aged organoids exhibit an enlarged central hypocellular/necrotic core and a more prominent peripheral αSMA/Vimentin-positive layer, consistent with structural remodeling during long-term culture. DAPI and 18S signals delineate the overall tissue architecture. Representative images are shown. Scale bar, 1000 µm.

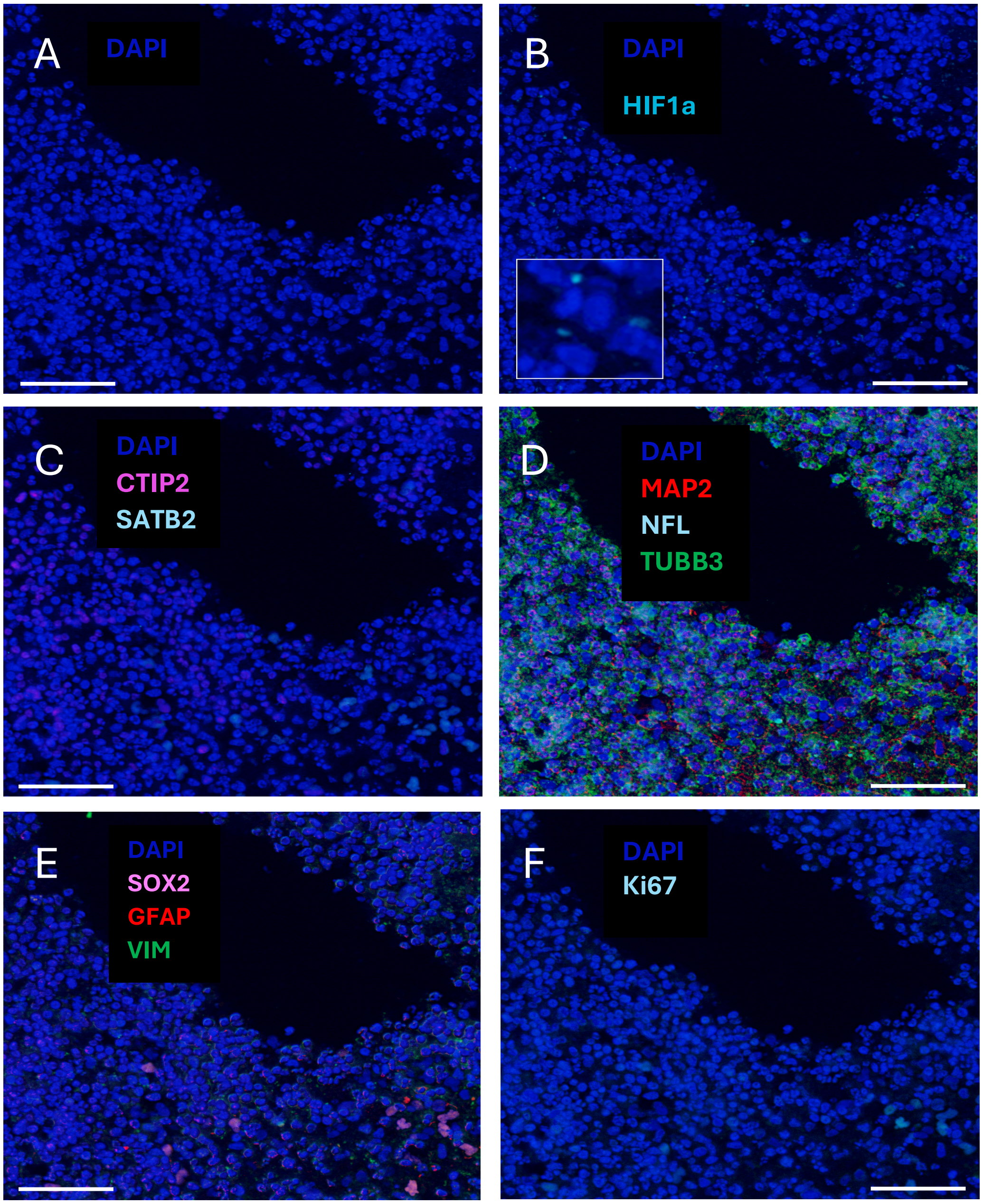


**Figure S2:** Tissue adjacent to post-processing cavities on day 98 cerebral organoid (donor BB026). To investigate if small cavities observed in pathology slides prepared for multicolor fluorescent microscopy show signs of tissue hypoxia or necrosis, we examined closely the surrounding tissue architecture.

A. DAPI staining does not show pyknotic nuclei. Scale bar, 50 μm.
B. Hypoxia-inducible factor 1 alpha (HIF1a) staining shows few cytoplasmic puncti of HIF1a consistent with normoxic conditions where continuously translated un-stabilized HIF1a is degraded in endosomal or autophagosomal compartments. Scale bar, 50 μm.
C. The neuronal lineage markers CTIP2 and SATB2 have correct nuclear localization. Scale bar, 50 μm.
D. Neuronal cytoarchitecture is preserved, with microtubule-associated protein (MAP2) and tubulin beta 3 class II (TUBB3) outlining intact cell bodies. Neurofilament light staining is sparse and punctate, consistent with staining in outermost layers of the organoid. Scale bar, 50 μm.
E. Astrocytes with SOX2+ nuclei are sparse and mostly proliferating. Scale bar, 50 μm.
F. Ki67 staining of proliferating cells), however, their intracytoplasmic levels of glial fibrillary acidic protein (GFAP) are modest and comparable to vimentin (VIM). Scale bar, 50 μm.

#### Cerebral organoid BB026 d40

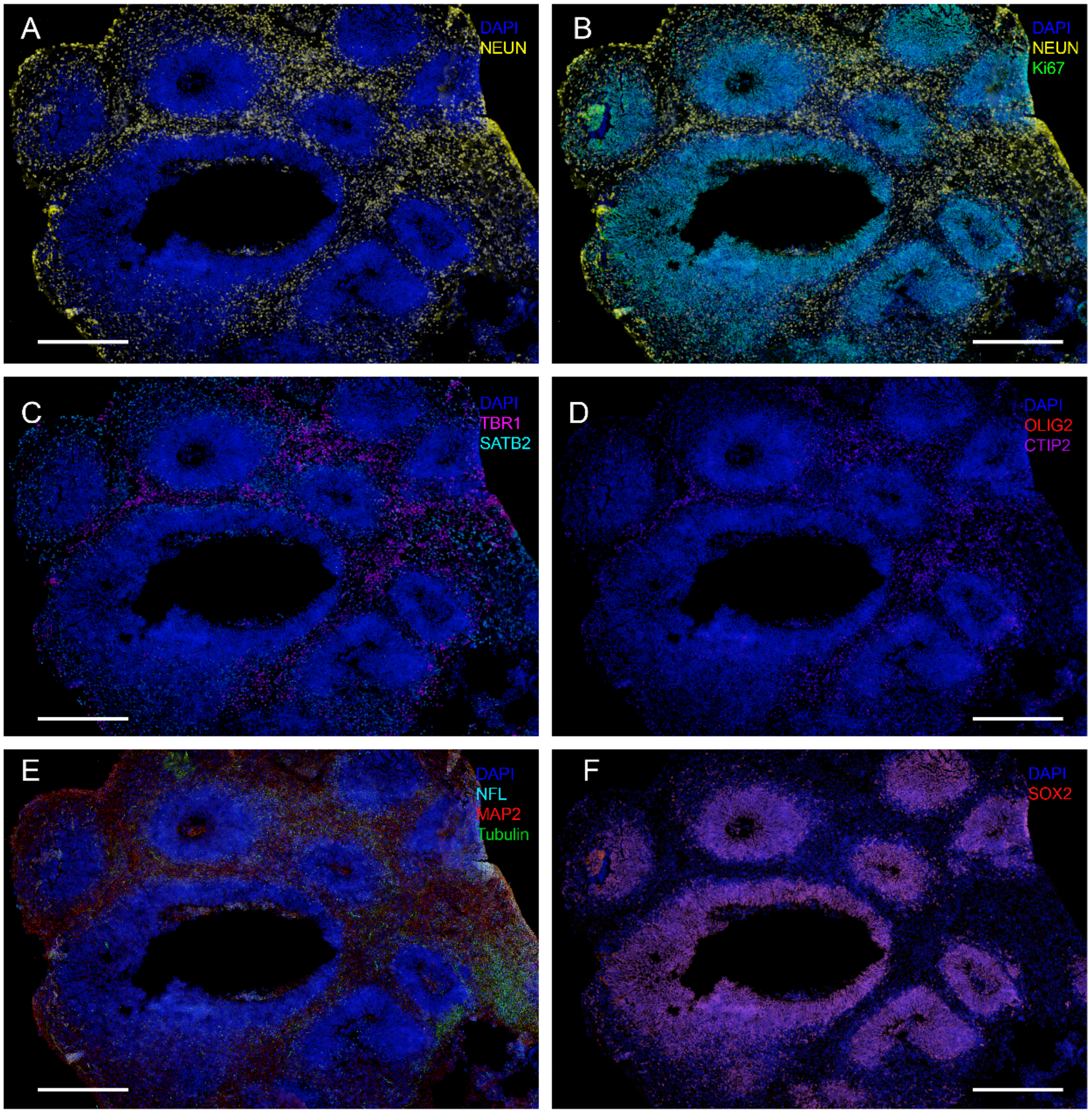


**Figure S3.** Organization of the neural architecture of cerebral organoid at day 40. DAPI (blue) and NEUN (yellow) stained regions indicate post-mitotic neural tissue. Scale bar, 300 μm.

A. Two separate populations of NEUN+ (yellow) post-mitotic neural tissue and the second Ki67+ (green) proliferating rosettes of immature neurons. Scale bar, 300 μm.
B. Deep layer projection neurons (cortical layer 2-6) stained with TBR1+ (pink) and SATB2+ (cyan) callosal projection neurons. Scale bar, 300 μm.
C. Deep layer projection neurons (cortical layer 5) stained with CTIP2+ (magenta) and OLIG2+ (red) oligodendrocyte progenitor cells diffused around rosettes. Scale bar, 300 μm.
D. MAP2+ (red) cells diffused through neural tissue, while Tubulin (TUBB3+, green) focused on region of deep-layer cells. Scale bar, 300 μm.
E. SOX2+ (red) co-localized in regions of Ki67+ rosettes. Scale bar, 300 μm.
Representative image of six organoids.

Cerebral organoid BB028 d140

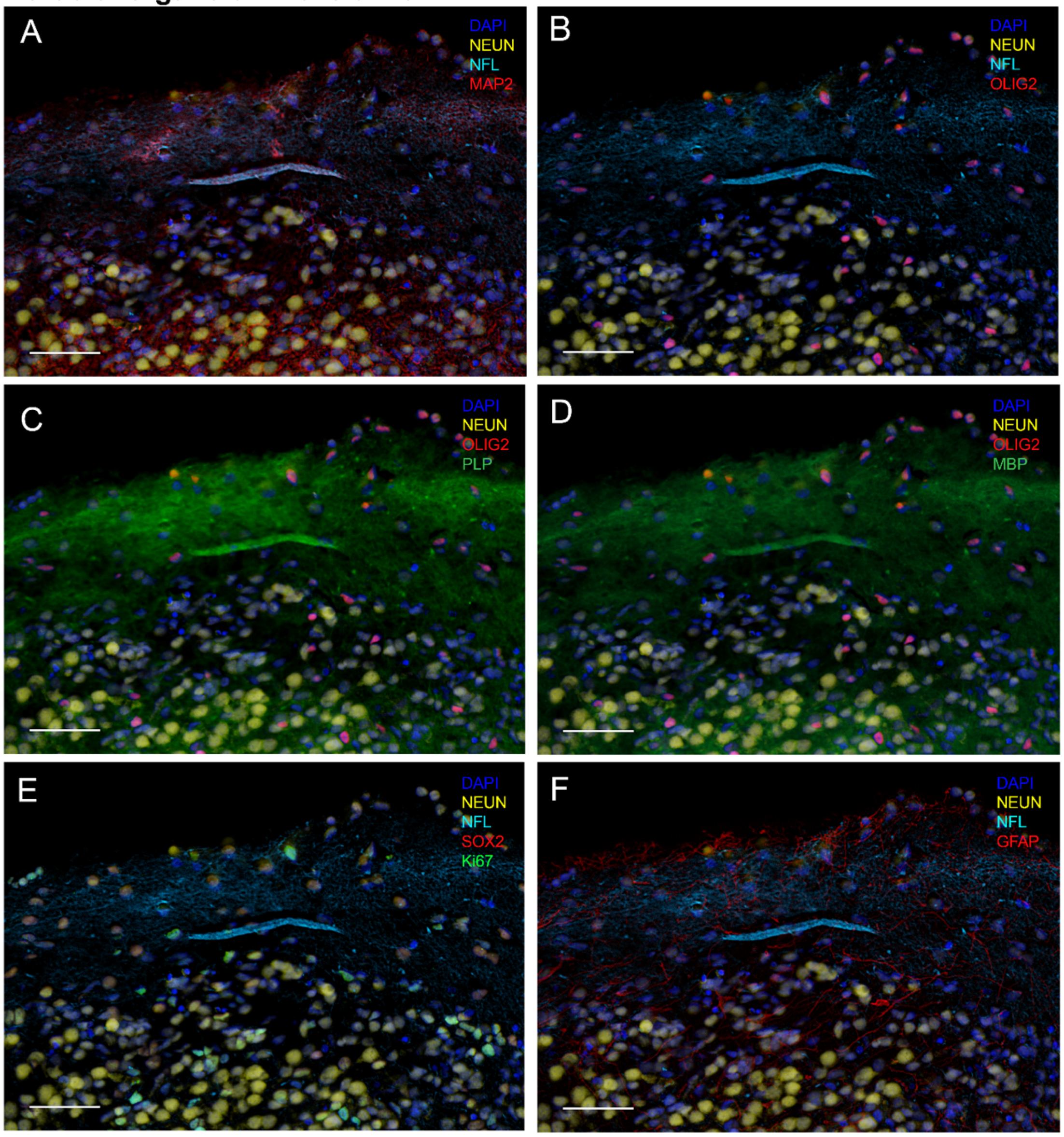


**Figure S4.** The increased presence of myelin proteins, MBP and PLP, co-expressed with neuronal axons indicates a larger and more mature population of myelinated axons at day 140.

A. Identified region of NEUN+ (yellow) neural tissue with NFL+ (cyan) axons, co-localized with MAP2 (red) extending throughout, indicating presence of mature neuronal architecture. Scale bar, 300 µm.
B. NFL (cyan)-rich neural tissue contains dispersed regions with OLIG2+ (red) oligodendrocyte and oligodendrocyte precursor cells. Scale bar, 300 µm.
C. OLIG2+ (red) cells abundant in PLP (green) expression, a myelin protein, extending across organoid outermost region. Scale bar, 300 µm.
D. OLIG2+ (red) regions abundant in MBP (green) expression, a myelin protein extending across organoid outermost region. Scale bar, 300 µm.
E. Outermost region of the organoid shows Ki67+ (green) proliferating cells. Scale bar, 300 µm.
F. Densely myelinated outer region (PLP+ and MBP+) co-expressing GFAP+ (red) astrocytic markers that boundary the edge of the organoid. Scale bar, 300 µm.

**Cerebral organoid BB026 d140**

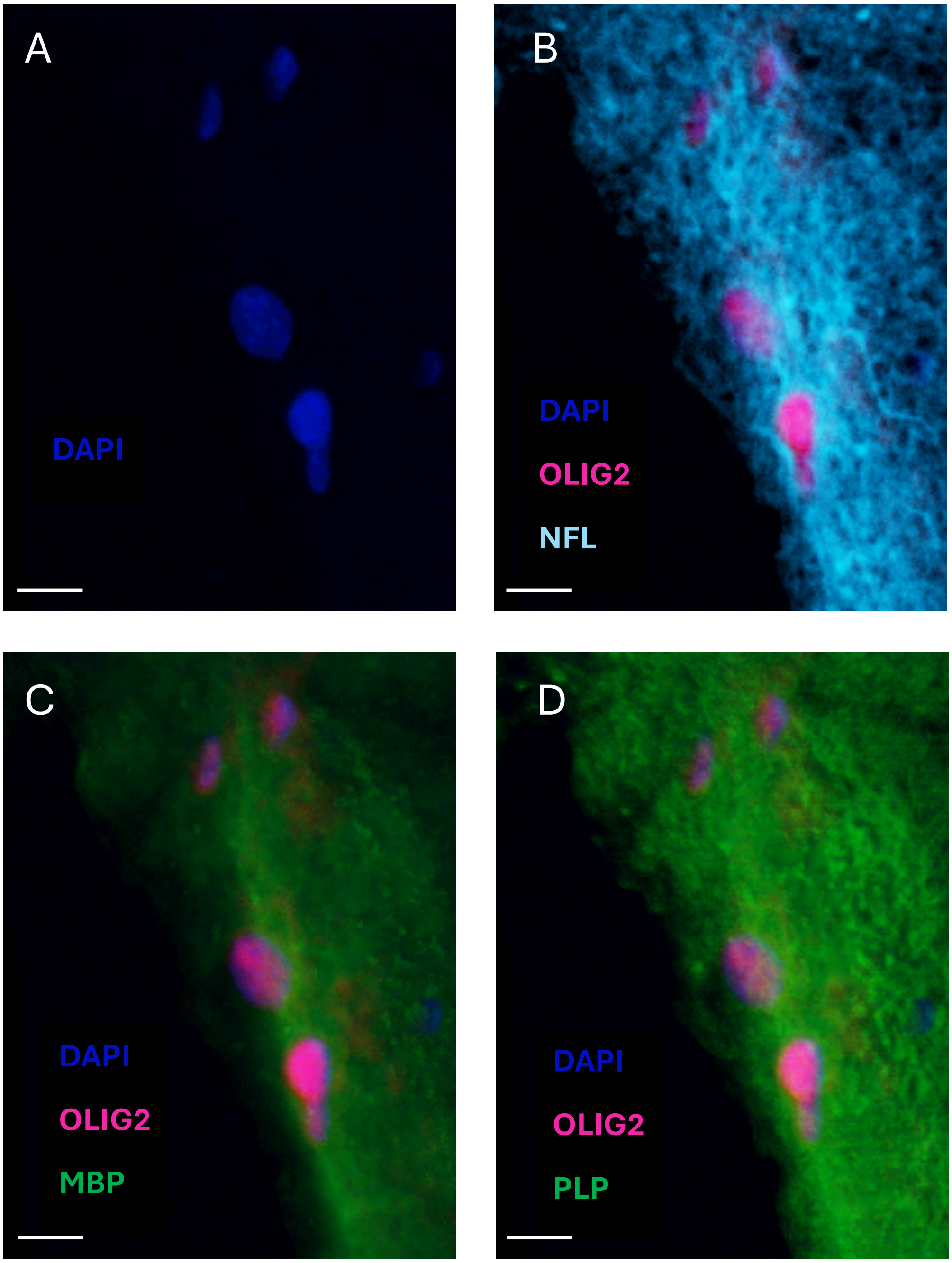


**Figure S5.** Representative high-magnification images from a day 140 cerebral organoid (donor BB026) showing OLIG2-positive oligodendrocyte lineage cells (magenta) in relation to NFL (cyan), MBP (green), and PLP (green).

A. DAPI. Scale bar, 10 µm.
B. DAPI, OLIG2, and NFL. Scale bar, 10 µm.
C. DAPI, OLIG2, and MBP. Scale bar, 10 µm.
D. DAPI, OLIG2, and PLP. Nuclei are counterstained with DAPI (blue). Scale bar, 10 µm.

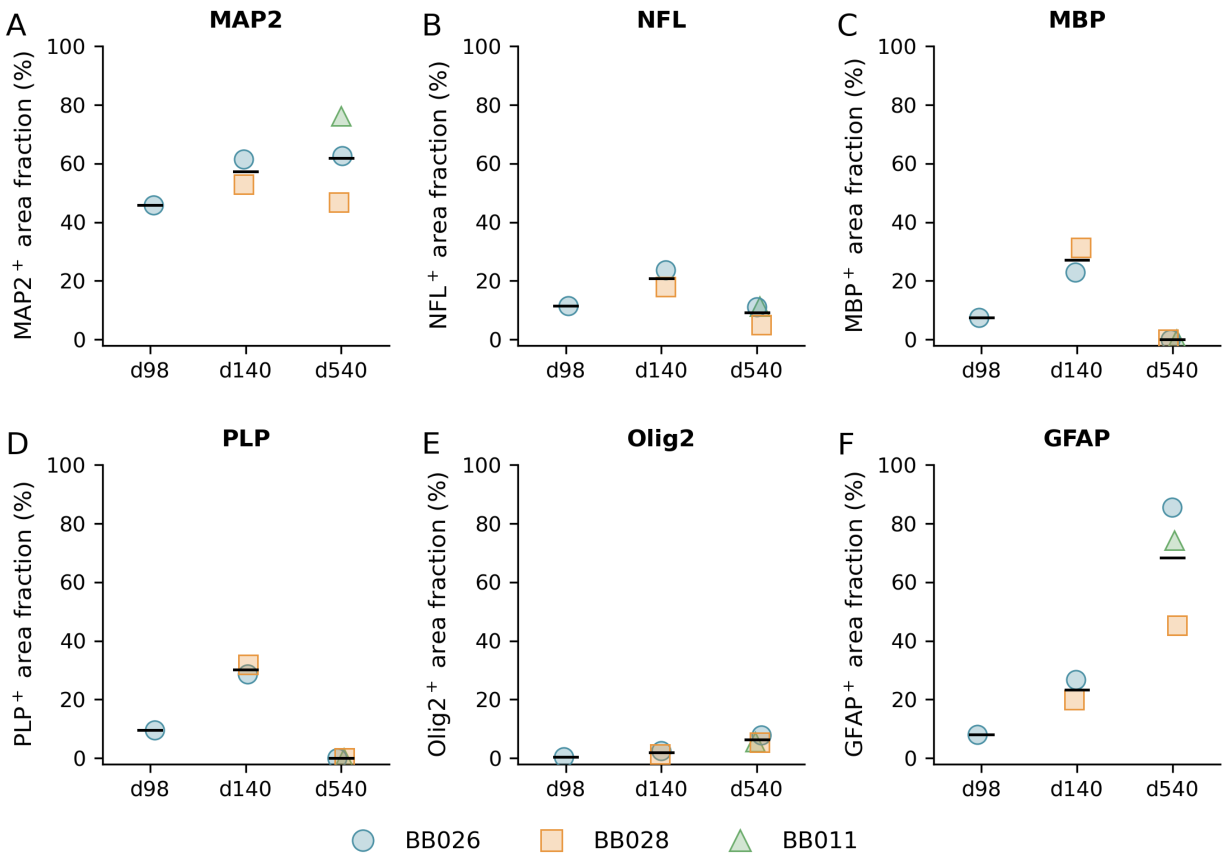


**Figure S6**. Immunofluorescence quantification of neuronal, oligodendroglial, and astrocytic markers during cerebral organoid maturation.

Marker-positive area, expressed as a percentage of the total organoid section area, for (A) MAP2, (B) neurofilament light chain (NFL), (C) myelin basic protein (MBP), (D) proteolipid protein (PLP), (E) Olig2, and (F) glial fibrillary acidic protein (GFAP) in cerebral organoids at days 98, 140, and 540 of differentiation. Each symbol represents an individual donor-derived organoid (BB026, BB028, or BB011). MAP2-positive area increased with organoid maturation, whereas NFL-positive area peaked at day 140 and declined by day 540. MBP- and PLP-positive areas were highest at day 140 and were not detected by immunofluorescence on day 540. In contrast, GFAP-positive area increased in aged organoids. Sample sizes were n = 1 (day 98), n = 2 (day 140), and n = 3 (day 540). One day 98 organoid (BB028) was excluded from the analysis due to tissue damage during multiplex immunofluorescence staining. Quantification was performed in Fiji (ImageJ) using threshold-based analysis.

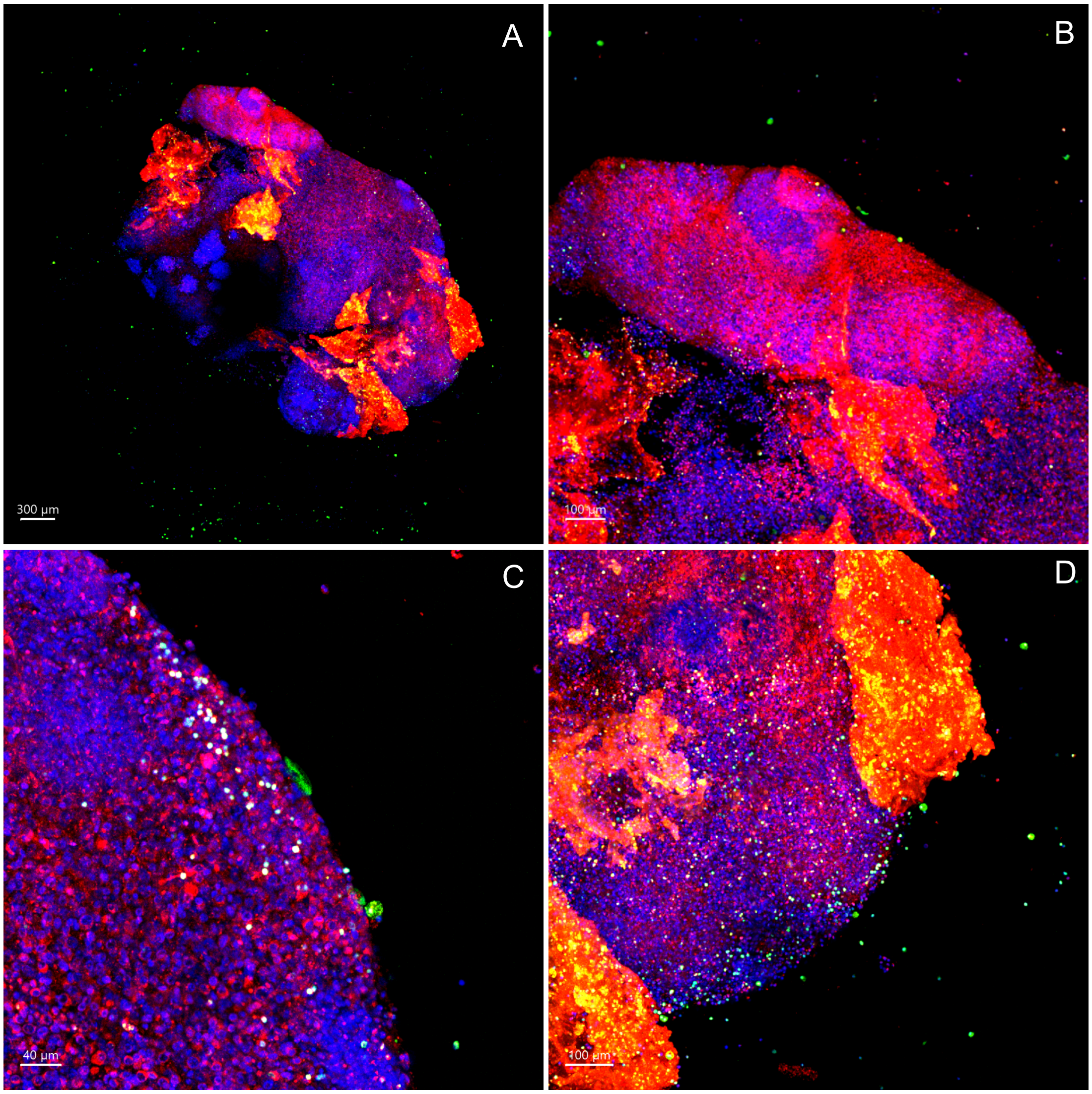


**Figure S7.** Incorporation of CX3CR1+/CD14+ microglia progenitors onto CO at d30.

A. Organoid (red) co-cultured with microglial progenitor cells (green). Scale bar, 300 µm.
B. Microglia progenitor cells (green) show minimal incorporation with organoid (red) on the outer layers unevenly around organoid. Scale bar, 100 µm.
C. Magnified images of microglia progenitor (green) incorporation with outer-layer of organoid (red). Regions of co-localization of DAPI, Red Tracker and Green Tracker reflect white. Scale bar, 40 µm.
D. Microglia progenitor cells (green) show minimal incorporation with organoid (red), There is uneven dispersion of microglia progenitors in the deeper layers of the organoid. Scale bar, 100 µm.

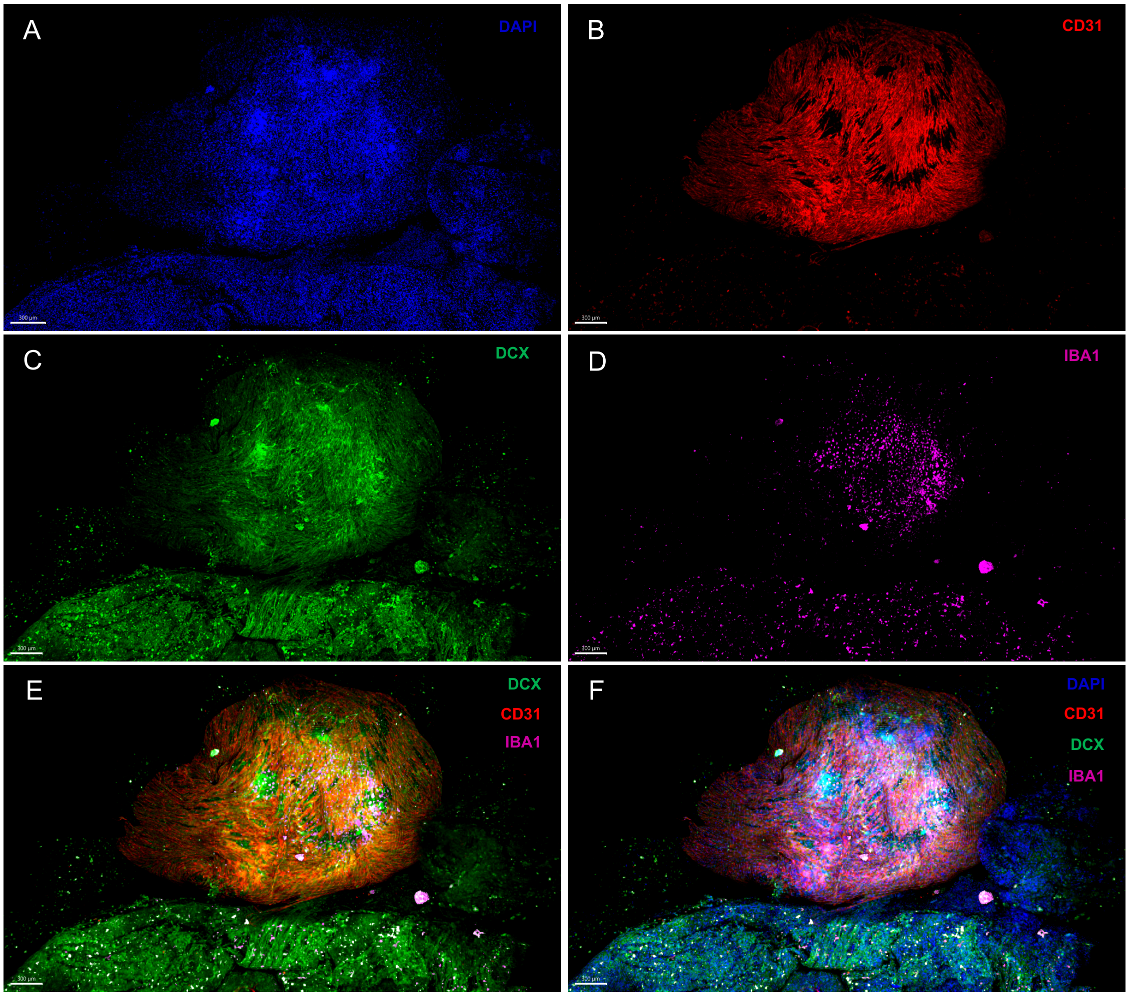


**Figure S8A.** Individual micrographs of assembloid to show CD31 and Iba1 engraftment onto neural tissue.

A–D. Individual micrographs showing nuclei (DAPI, blue), endothelial vascularization (CD31$^{+}$, red), immature neurons (DCX$^{+}$, green), and microglia-like cells (IBA1$^{+}$, magenta). Scale bar, 300 µm.
E. Fused assembloid showing infiltration of neural cells (DCX$^{+}$, green), vascular structures (CD31$^{+}$, red), and engrafted microglia-like cells (IBA1$^{+}$, magenta). Scale bar, 300 µm.
F. Composite overlay of DAPI with DCX, CD31, and IBA1 immunostaining in the fused assembloid. Scale bar, 300 µm.

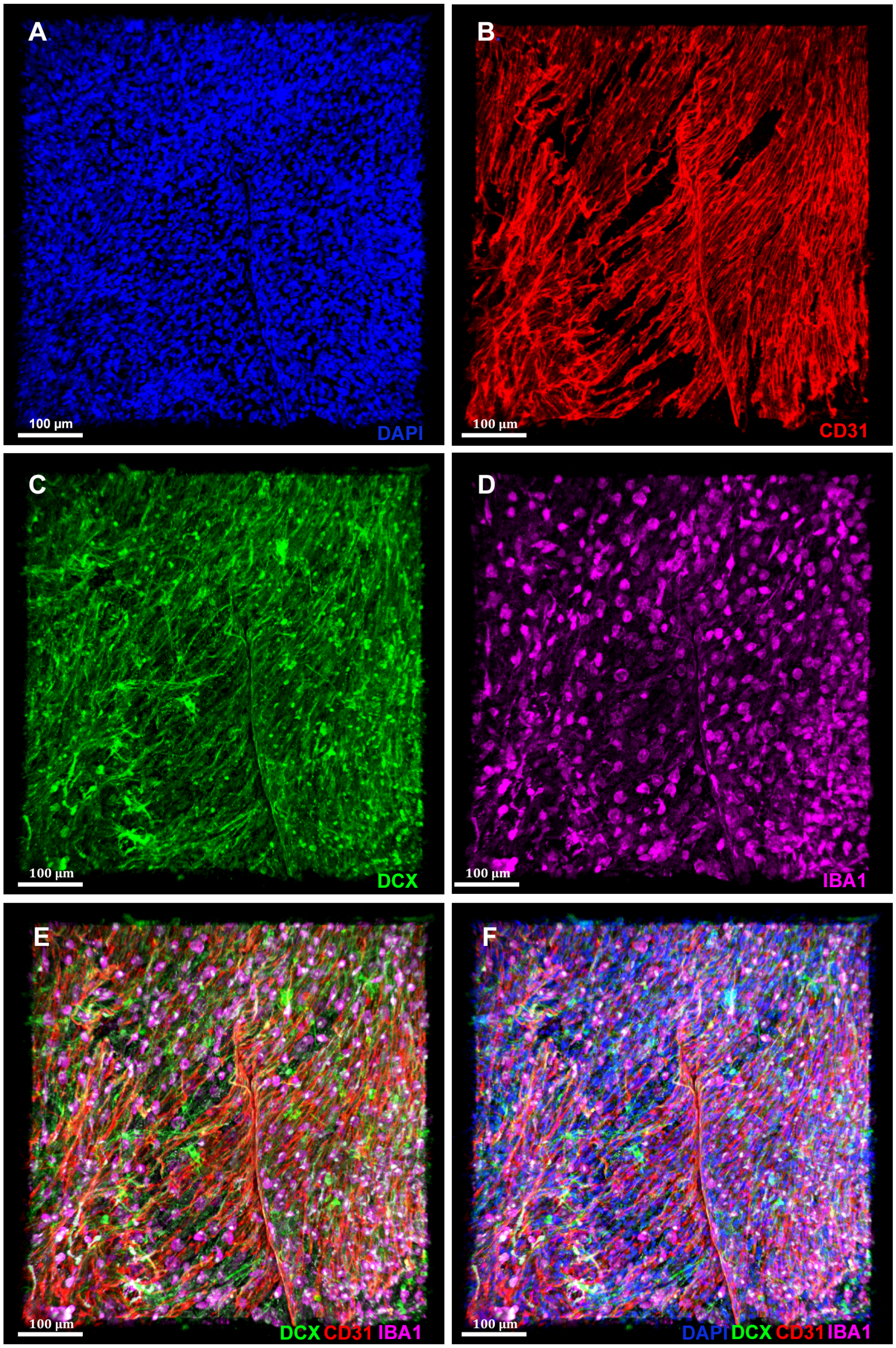


**Figure S8B.** Higher-magnification images of the assembloid (day 45) shown in Figure S8A, highlighting the engraftment of CD31$^{+}$ endothelial cells and IBA1$^{+}$ microglia within the neural tissue. Scale bar, 100 µm.

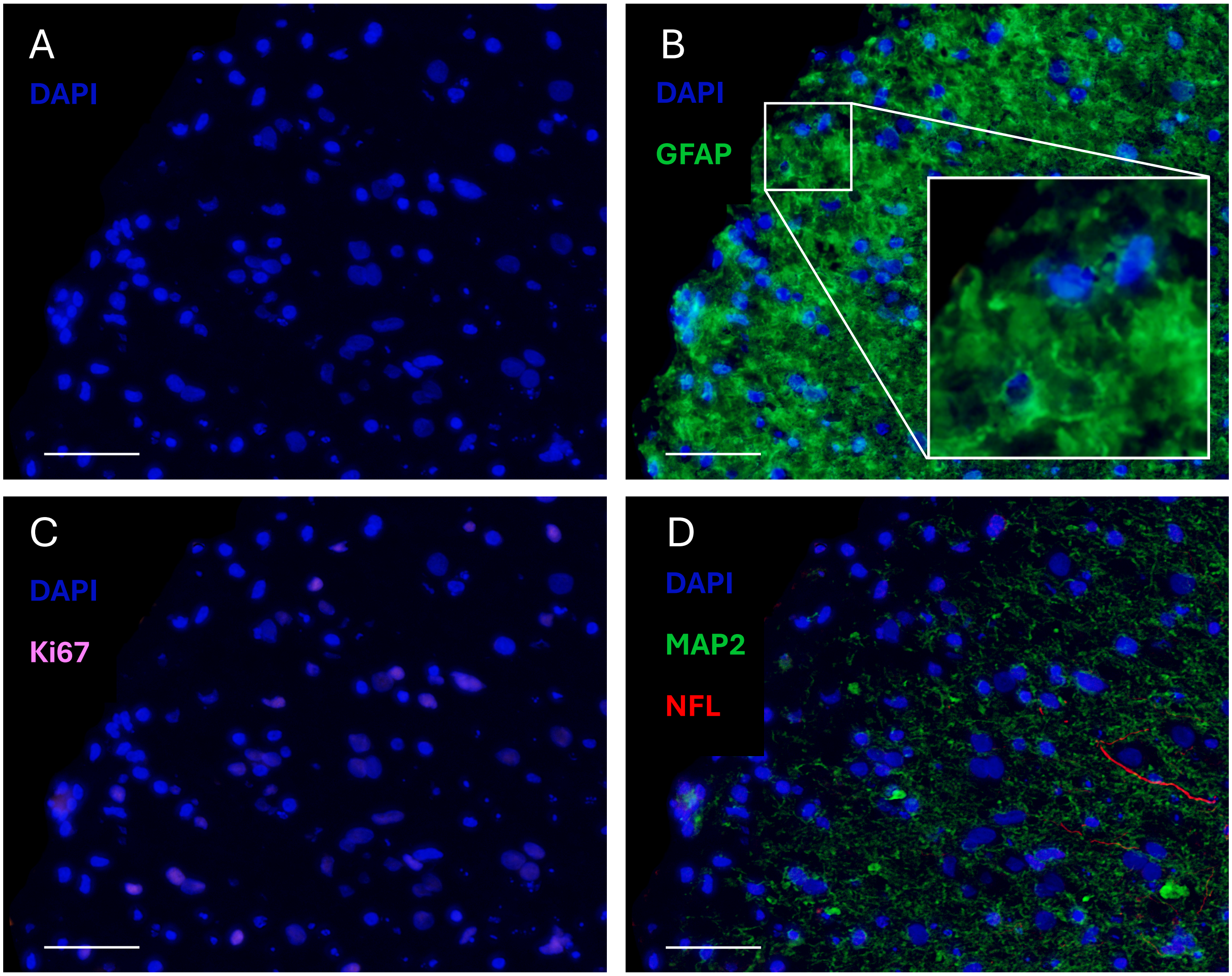


**Figure S9:** Astrocyte morphology and neuronal architecture in aged cerebral organoids**.** Representative high-magnification images from a day 540 cerebral organoid (donor BB011).

A. DAPI staining (blue). Scale bar, 50 μm.
B. GFAP-positive astrocytes (green) with a higher magnification inset showing astrocytic morphology.
C. Ki67-positive proliferating cells (magenta). Scale bar, 50 μm.
D. MAP2-positive neuronal processes (green) and NFL-positive neurites (red). Scale bar, 50 μm.

A-D. Nuclei are counterstained with DAPI (blue). Representative images are shown. Scale bar, 50 μm.

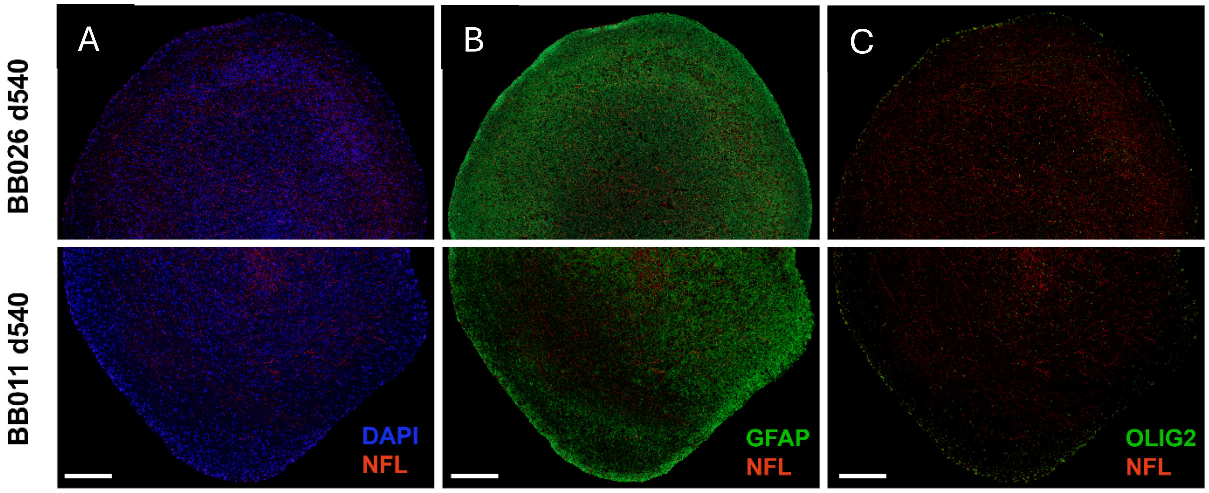


**Figure S10.** Neural architecture at day 540 (d540) reveals increased astrogliosis, characterized by an expansion of $GFAP^{+}$ cells, accompanied by marked loss of neuro-axonal markers and evidence of hypomyelination.

A. DAPI (blue) and NFL+ (red) axons stained in MS-derived CO (BB011) and healthy-donor derived (BB026) organoids. NFL+ axons localized to central regions of CO. Scale bar, 250 µm.
B. GFAP+ (green) and NFL+ (red) staining shows overwhelming astrogliosis in both organoids by d540. Scale bar, 250 µm.
C. OLIG2+ (green) cells only in peripheral of organoid where GFAP+ is predominantly present, while axonal staining (NFL+; red) is localized to the central regions of CO. Scale bar, 250 µm.

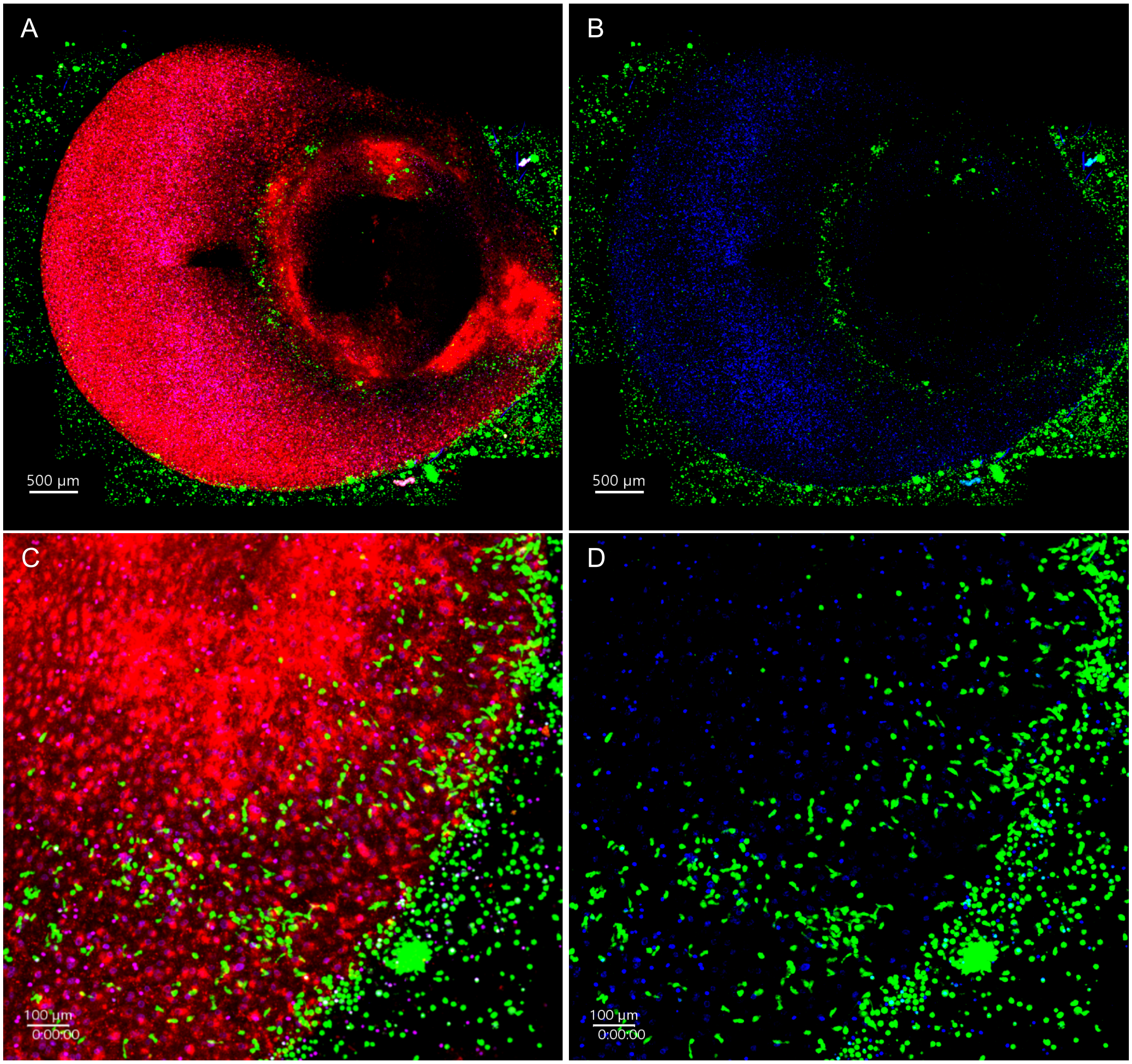


**Figure S11.** Micrographs from real-time fluorescence microscopy to monitor immune cell infiltration kinetics.

A. Neural tissue (red) from healthy donor-derived CO (BB028) 24h post- incubation with activated PBMC (green). Scale bar, 500 µm.
B. Removed neural (red) overlay to emphasize the number of PBMC infiltration onto and around the superficial tissue of the CO. Scale bar, 500µm.
C. Magnified region of neural tissue (red) shows uptake and incorporation of PBMC (green) into the CO after 24h of incubation. Scale bar, 100 µm at 0:00:00 of video 2.
D. Magnified region of neural (red) overlay removed from Figure S5C to emphasize uptake and incorporation of PBMC (green) into the CO after 24h of incubation. Scale bar, 100 µm at 0:00:00 of video 2.

   Representative time-lapse video of a cerebral organoid following 24 h incubation with activated PBMC. Three organoids were recorded with similar results.

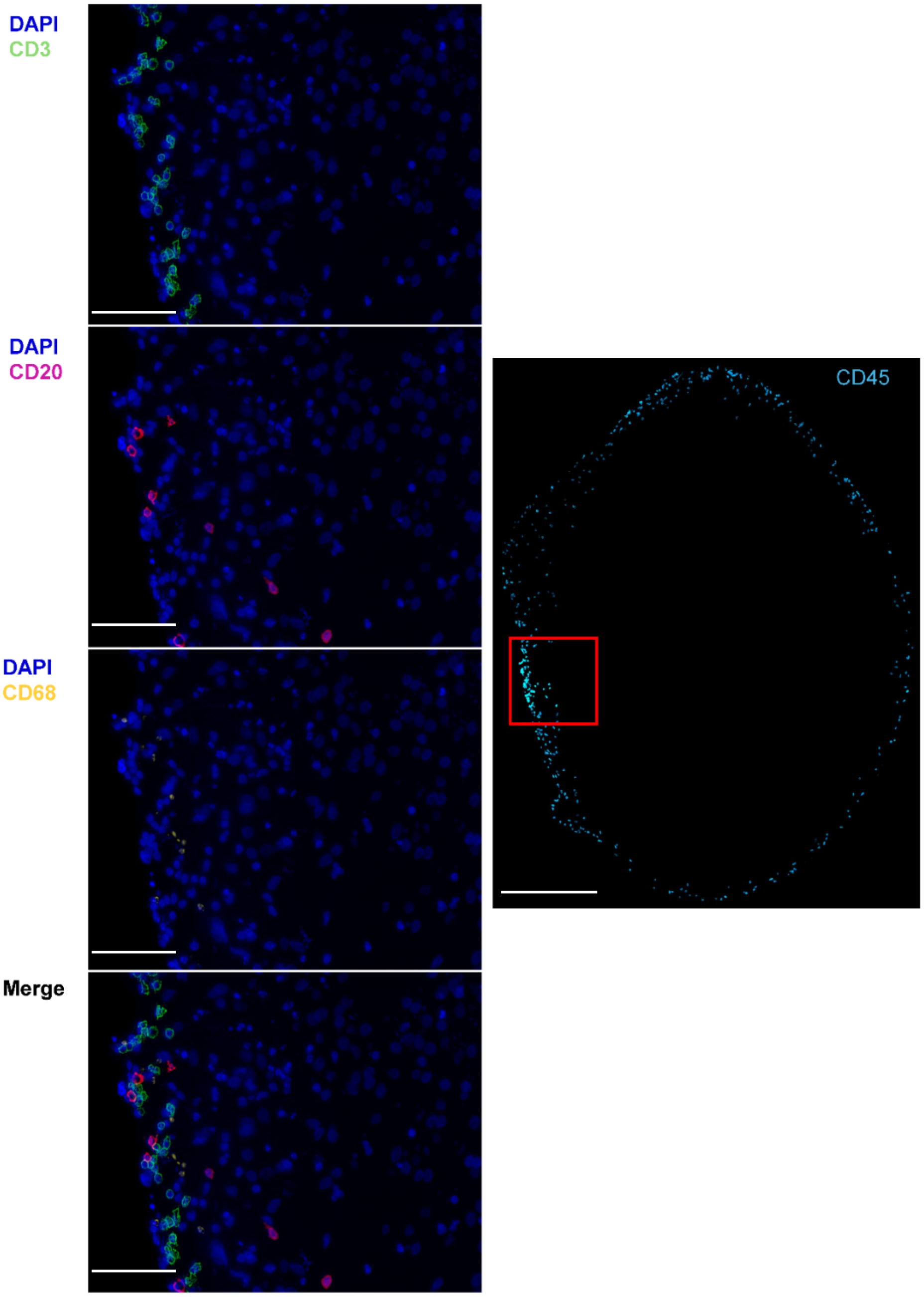


**Figure S12.** Immune cell infiltration into BB011 cerebral organoids following 24-hour co-culture with autologous activated PBMC.

Immune cell incorporation into MS-derived CO (BB011) 24h after co-culture with activated PBMC. Immune cell composition was determined via CD3+ (cyan) T-cells, CD20+ (magenta) B-cells, and CD68+ (yellow) macrophage (left panel). PBMC incorporation was determined via CD45+ cells (blue) (right panel). Scale bar, 100 μm left panel and 500 μm (right panel).

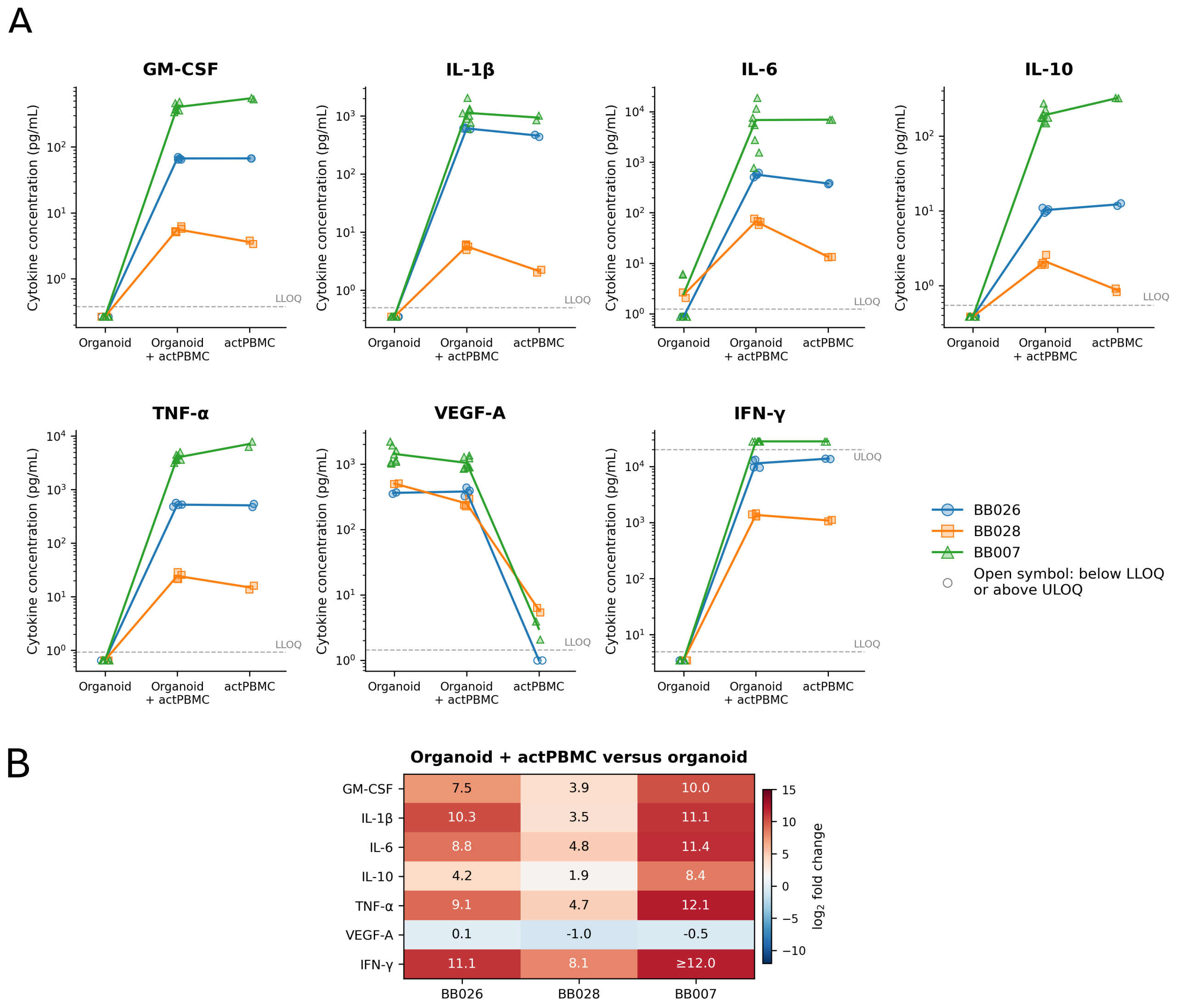


**Figure S13.** Activated PBMC establish a pro-inflammatory cytokine environment in cerebral organoid co-cultures.

A. Concentrations of GM-CSF, IL-1β, IL-6, IL-10, TNF-α, INF-γ and VEGF-A measured in conditioned medium from cerebral organoids cultured alone, organoids co-cultured with activated autologous PBMC (organoid+actPBMC), and activated PBMC cultured alone (actPBMC). Data are shown for individual donors (BB026, BB028, and BB007). Symbols represent biological replicates; open circles indicate values below (LLOQ) or above (ULOQ) the detection limit. Cytokine concentrations are plotted on a logarithmic scale and expressed as pg/mL).
B. Fold change in cytokine concentrations in organoid–actPBMC co-cultures relative to organoids cultured alone for each donor. Numbers within the heatmap indicate fold change, with color intensity corresponding to the magnitude of the increase or decrease. Please note that activated PBMC from bb028 were rested for 72h before co-culture with CO, as defined in Methods.
The primary repeated-measures ANOVA confirmed highly significant treatment effects across conditions for 6 of the 7 measured analytes (false discovery rate [FDR] $q < 0.05$). Subsequent post-hoc paired *t*-tests ($df = 2$) on donor geometric means revealed nominal statistical significance ($p < 0.05$) for key comparisons, including the upregulation of IL-6 ($p = 0.047$) and IFN-γ ($p = 0.013$) in co-culture versus organoid alone, as well as the robust modulation of VEGF-A ($p = 0.0085$ for actPBMC versus organoid). Although individual pairwise contrasts were underpowered due to the small sample size ($n$ = 3 donors) and yielded no statistically significant differences after FDR correction, we observed highly consistent, directional treatment effects across all three donors.

### BB007 CNS Composition – Per Organoid

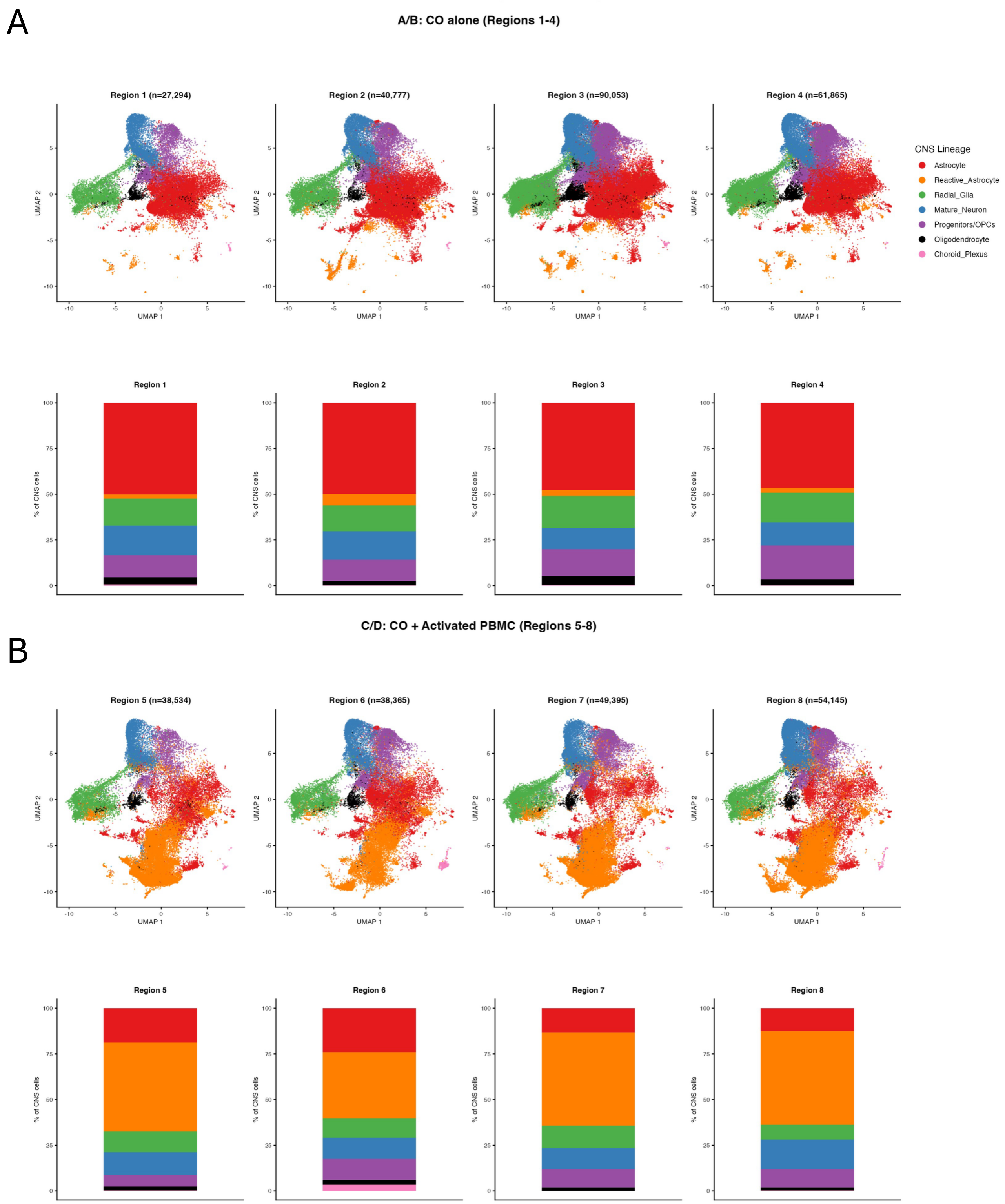

**Figure S14.** Cellular composition of day 200 cerebral organoids with and without activated PBMC co-culture.

A. UMAP visualization of major CNS cell populations identified in individual regions of a day 200 cerebral organoid derived from donor BB007 cultured alone (Regions 1–4; n=4).
B. UMAP visualization of individual regions from a day 200 cerebral organoid derived from donor BB007 following co-culture with activated PBMC (Regions 5–8; n=4).
   Each point represents a single cell colored by cell identity. Stacked bar plots below each UMAP show the relative proportion of the identified CNS cell populations in the corresponding region. Cell populations were annotated as astrocytes, reactive astrocytes, radial glia, mature neurons, progenitors/oligodendrocyte precursor cells (OPCs), oligodendrocytes, and choroid plexus cells. Numbers above each UMAP indicate the total number of cells analyzed in each region. Cell identities were assigned based on canonical marker gene expressions as described in the Methods.

A

| Feature | Young (d200) | Old (d750) |
|---|---|---|
| **Metabolic state** | Oxidative phosphorylation, lactate shuttle | Aerobic glycolysis (Warburg), HIF-driven |
| **Reactivity type** | Interferon/complement (CXCL10, C4B) | Senescent/neurotoxic (GFAP, CHI3L1, CD44, SERPINA3) |
| **ECM/structure** | Proteoglycan-rich (BCAN, NDRG2/4) | Fibrotic remodeling (TNC, SDC4, ITGB1) |
| **Neurosupport** | Active (APOE, ABCA1, PEA15, FGFR3) | Diminished |
| **Senescence** | Absent | Active (GADD45A, MDM2, CDKN1A) |
| **Paracrine signals** | Complement (C4B) | Myeloid recruitment (CSF1), adhesion (ICAM1) |

B

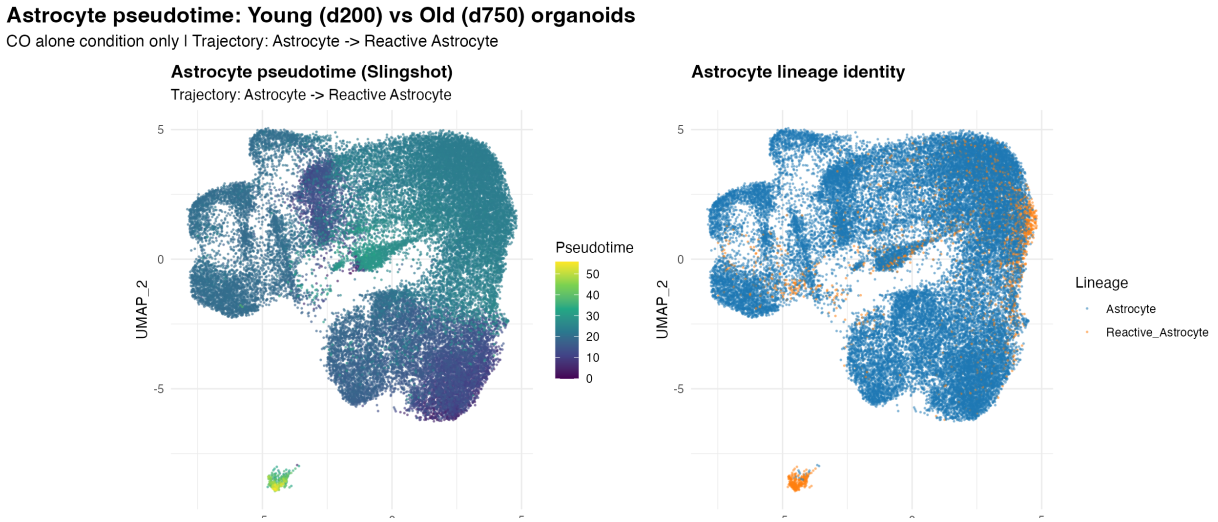


**Figure S15.** Comparison of transcriptional features and pseudotime trajectory of astrocytes from young and aged cerebral organoids.

A. Summary of the major transcriptional characteristics distinguishing astrocytes from young (day 200) and aged (day 750) cerebral organoids, including metabolic state, reactive phenotype, extracellular matrix (ECM) remodeling, neurosupportive functions, senescence-associated features, and paracrine signaling pathways.
B. Pseudotime analysis of astrocytes from day 200 and day 750 cerebral organoids under control conditions. Left, pseudotime trajectory inferred using Slingshot, illustrating the progression from astrocytes to reactive astrocytes. Right, the same trajectory colored by astrocyte lineage identity.

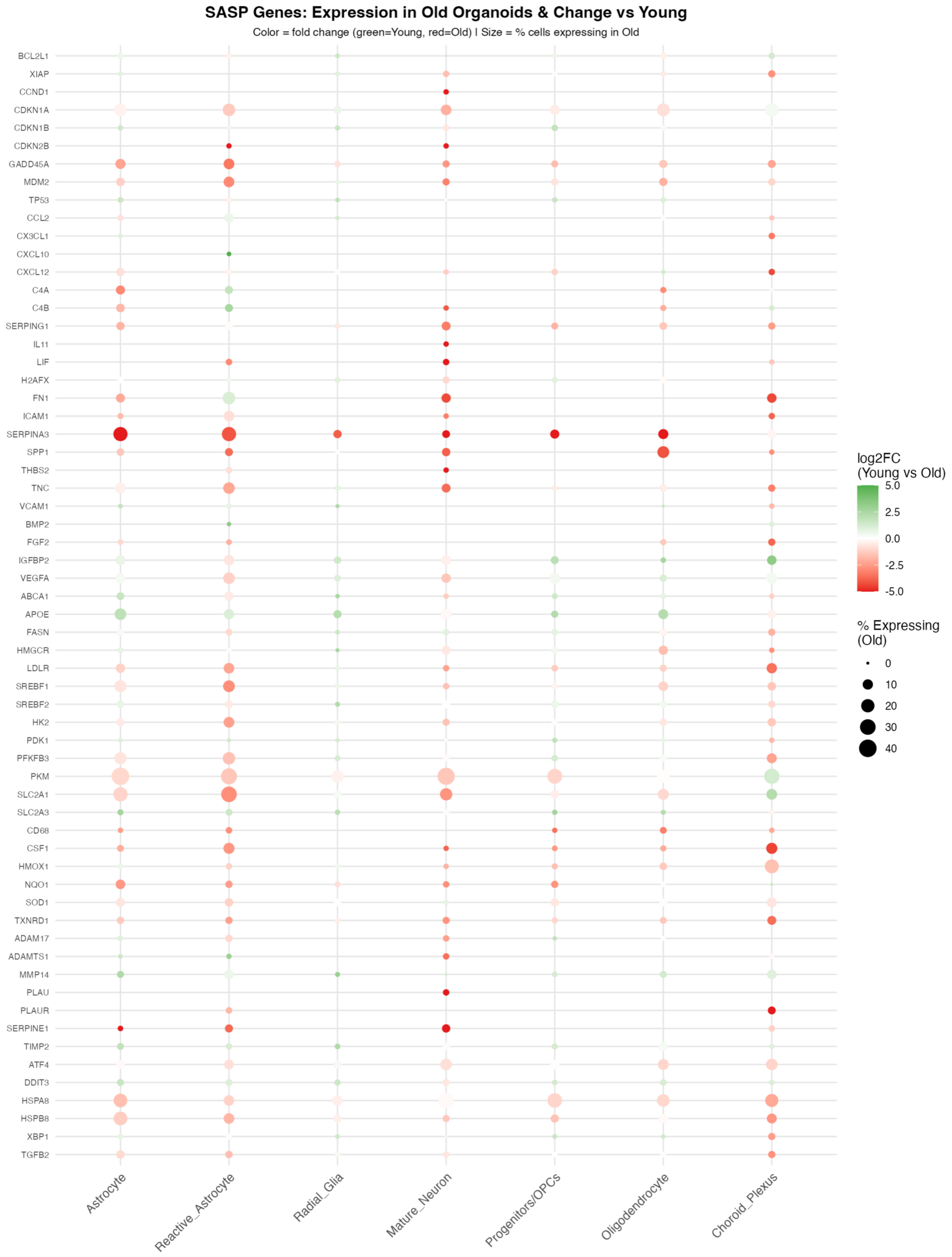

SASP Genes: Expression in Old Organoids & Change vs Young
Color = fold change (green=Young, red=Old) | Size = % cells expressing in Old
BCL2L1
XIAP
CCND1
CDKN1A
CDKN1B
CDKN2B
GADD45A
MDM2
TP53
CCL2
CX3CL1
CXCL10
CXCL12
C4A
C4B
SERPING1
IL11
LIF
H2AFX
FN1
ICAM1
SERPINA3
SPP1
THBS2
TNC
VCAM1
BMP2
FGF2
IGFBP2
VEGFA
ABCA1
APOE
FASN
HMGCR
LDLR
SREBF1
SREBF2
HK2
PDK1
PFKFB3
PKM
SLC2A1
SLC2A3
CD68
CSF1
HMOX1
NQO1
SOD1
TXNRD1
ADAM17
ADAMTS1
MMP14
PLAU
PLAUR
SERPINE1
TIMP2
ATF4
DDIT3
HSPA8
HSPB8
XBP1
TGFB2
Astrocyte
Reactive_Astrocyte
Radial_Glia
Mature_Neuron
Progenitors/OPCs
Oligodendrocyte
Choroid_Plexus
log2FC
(Young vs Old)
5.0
2.5
0.0
-2.5
-5.0
% Expressing
(Old)
0
10
20
30
40

**Figure S16.** Cell type-specific expression of senescence-associated secretory phenotype (SASP)-related genes in young and aged cerebral organoids.

Dot plot summarizing the expression of selected SASP-related genes across the major CNS cell populations identified by Xenium spatial transcriptomics. Dot color indicates the log2fold change (log2FC) between young (day 200) and aged (day 750) organoids (green, enriched in young; red, enriched in aged), while dot size represents the percentage of cells expressing each gene in the aged organoids. Cell populations include astrocytes, reactive astrocytes, radial glia, mature neurons, progenitor/oligodendrocyte precursor cells (OPCs), oligodendrocytes, and choroid plexus cells.

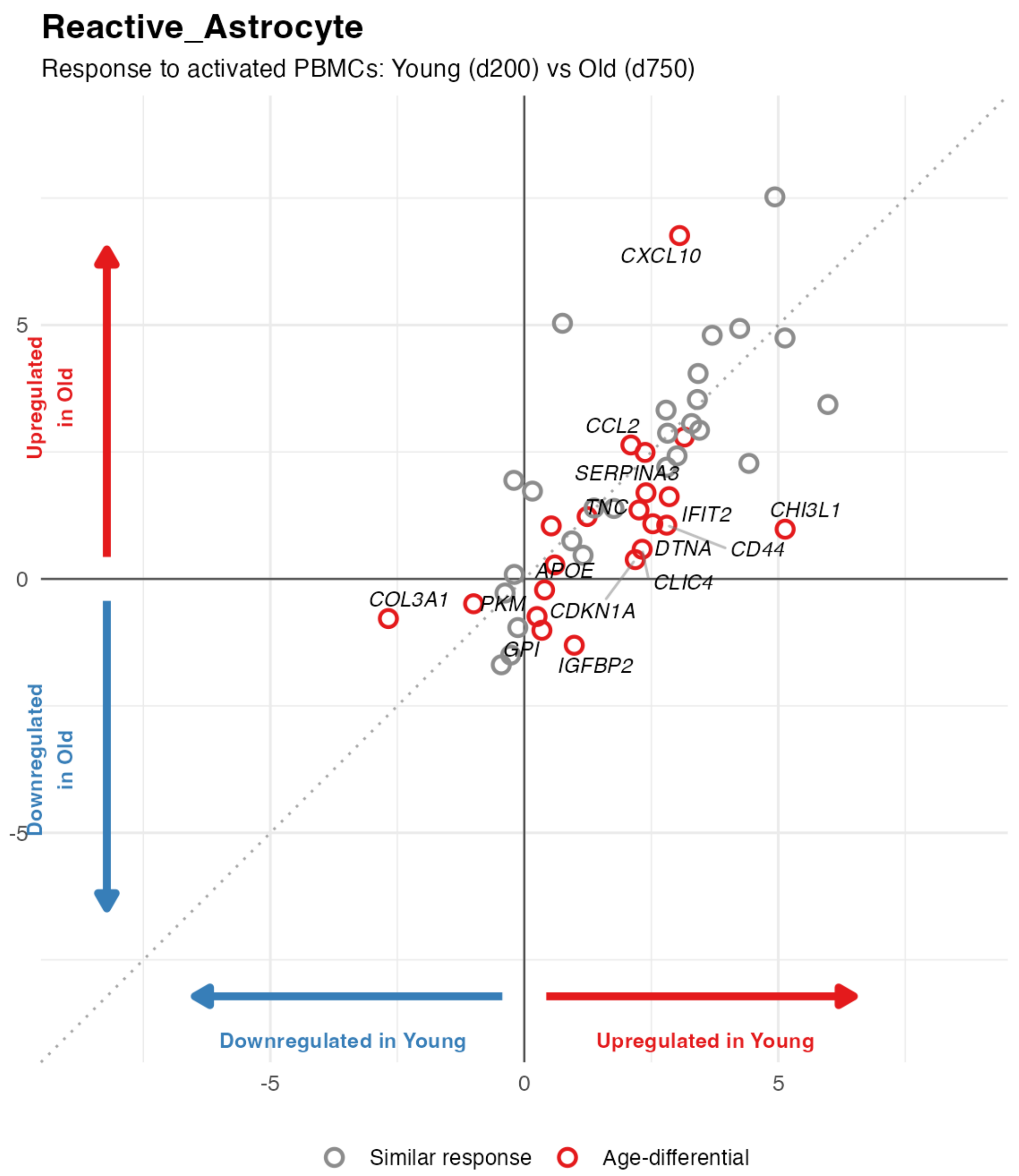

Reactive_Astrocyte
Response to activated PBMCs: Young (d200) vs Old (d750)
Upregulated in Old
Downregulated in Old
Downregulated in Young
Upregulated in Young
CXCL10
CCL2
SERPINA3
TNC
IFIT2
CHI3L1
DTNA
CD44
APOE
CLIC4
COL3A1
PKM
CDKN1A
GPI
IGFBP2
5
0
-5
-5
0
5
Similar response
Age-differential

**Figure S17.** Age-dependent transcriptional responses of reactive astrocytes to activated PBMC co-culture.

Scatter plot comparing gene expression responses to activated PBMC co-culture in reactive astrocytes from young cerebral organoids (day 200; donor BB007, CO alone: *n* = 4 organoids, CO + ActPBMC: *n* = 4 organoids) and aged cerebral organoids (day 750; donors BB026 and BB028, CO alone: *n* = 1 organoid each donor (2 organoids total), CO + ActPBMC: *n* = 2 organoids each, (4 organoids total)). Each point represents a differentially expressed gene in reactive astrocytes. The x-axis (log2FC) shows the transcriptional response in young organoids, whereas the y-axis shows the corresponding (log2FC) transcriptional response in aged organoids. Gray circles indicate genes exhibiting similar transcriptional responses in both age groups, whereas red circles highlight genes with age-dependent responses. Selected genes with pronounced age-associated differences are labeled. The diagonal dashed line denotes equal transcriptional responses in young and aged organoids.

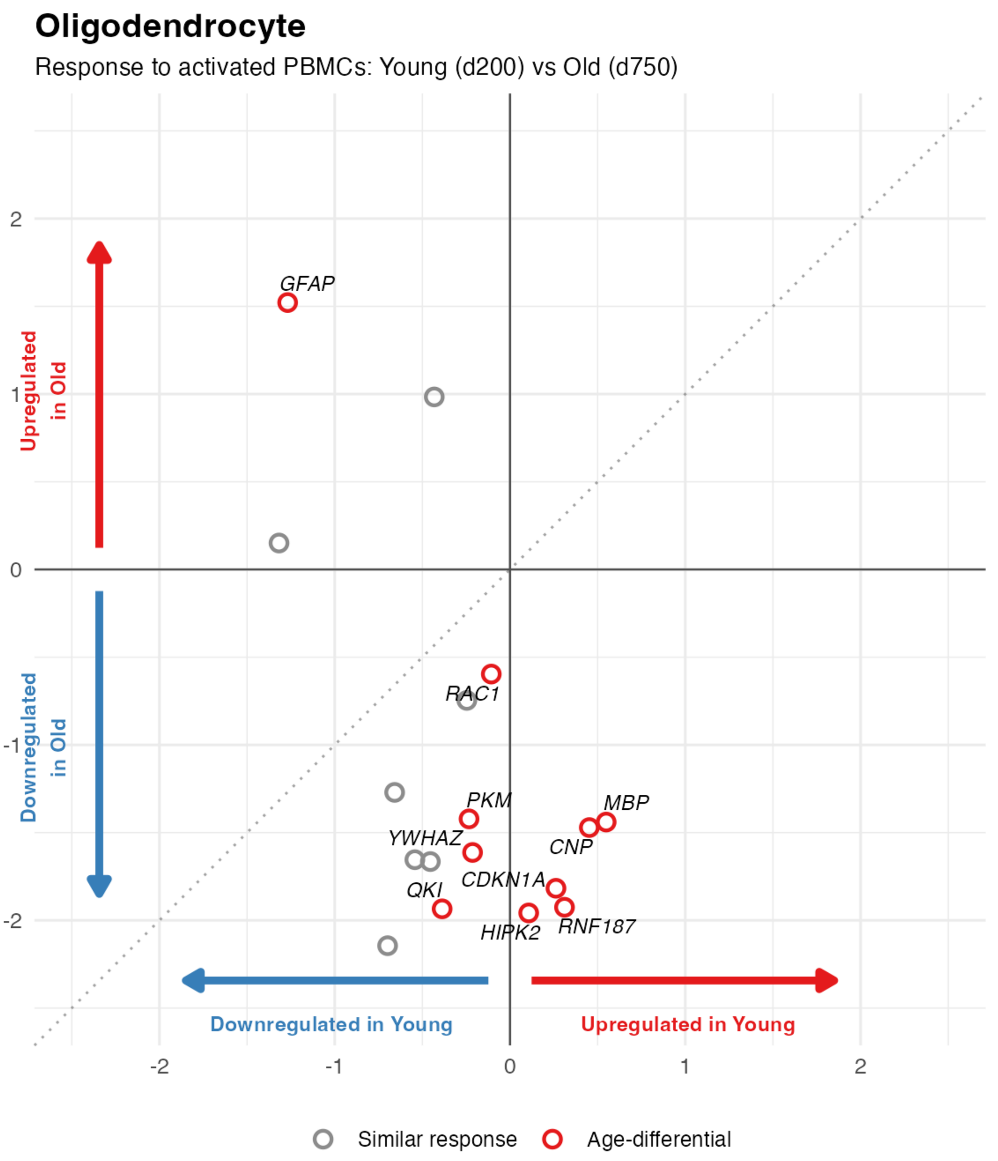


**Figure S18.** Age-dependent transcriptional responses of oligodendrocytes to activated PBMC co-culture.

Scatter plot comparing gene expression responses to activated PBMC co-culture in reactive oligodendrocytes from young cerebral organoids (day 200; donor BB007, CO alone: *n* = 4 organoids, CO + ActPBMC: *n* = 4 organoids) and aged cerebral organoids (day 750; donors BB026 and BB028, CO alone: *n* = 1 organoid each donor (2 organoids total), CO + ActPBMC: *n* = 2 organoids each donor (2 organoids total). Each point represents a differentially expressed gene in oligodendrocytes. The x-axis (log2FC) shows the transcriptional response in young organoids, whereas the y-axis shows the corresponding (log2FC) transcriptional response in aged organoids. Gray circles indicate genes exhibiting similar transcriptional responses in both age groups, whereas red circles highlight genes with age-dependent responses. Selected genes with pronounced age-associated differences are labeled. The diagonal dashed line denotes equal transcriptional responses in young and aged organoids.

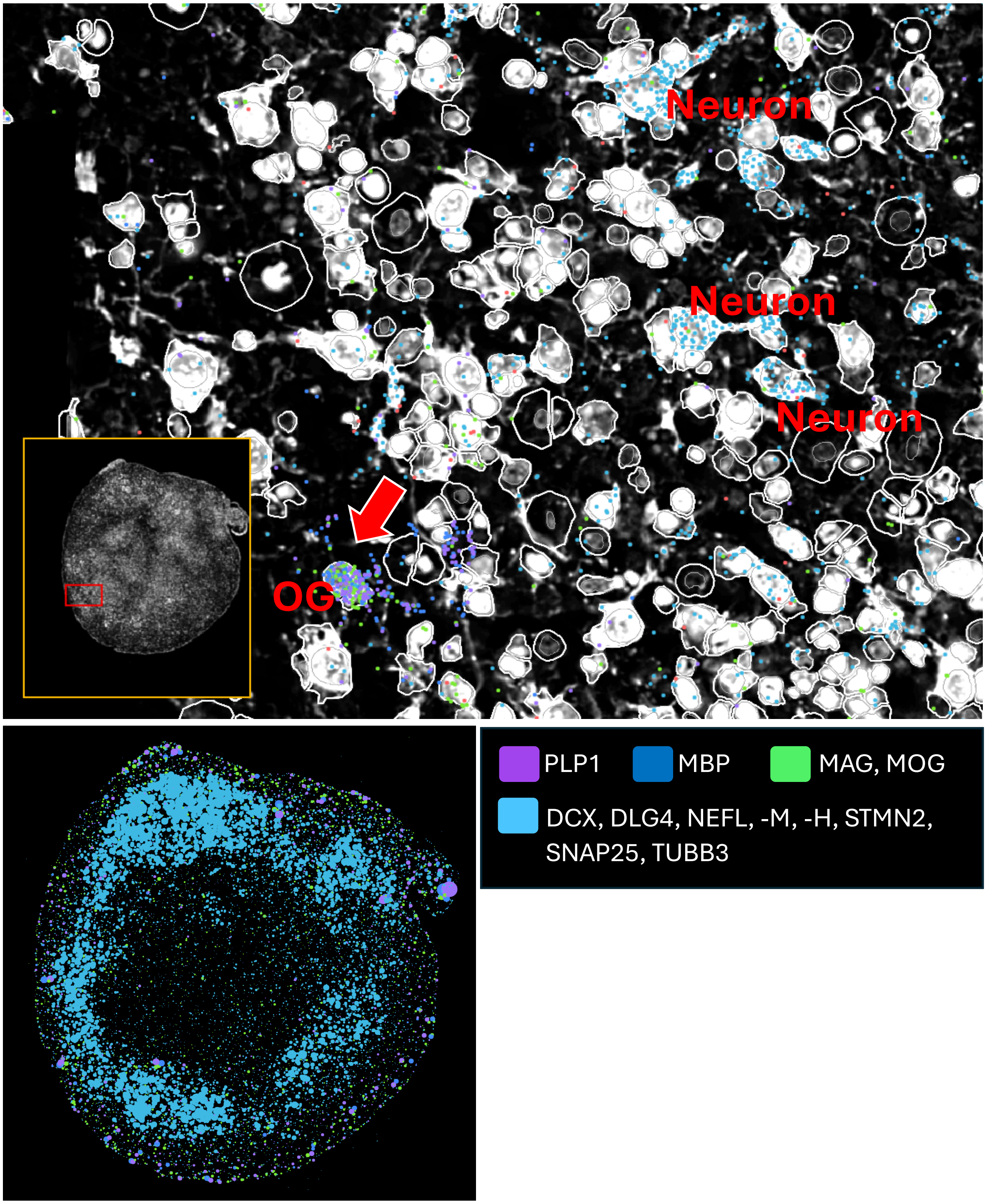


**Figure S19.** Representative Xenium spatial transcriptomic image of a day 200 cerebral organoid (donor BB007, CO alone) showing myelin-associated transcripts outlining oligodendrocyte (OG) processes. Myelin-associated transcripts were identified by the co-localization of *MBP*, *PLP1*, *MAG*, and *MOG* transcripts, revealing their linear organization along oligodendrocyte processes.

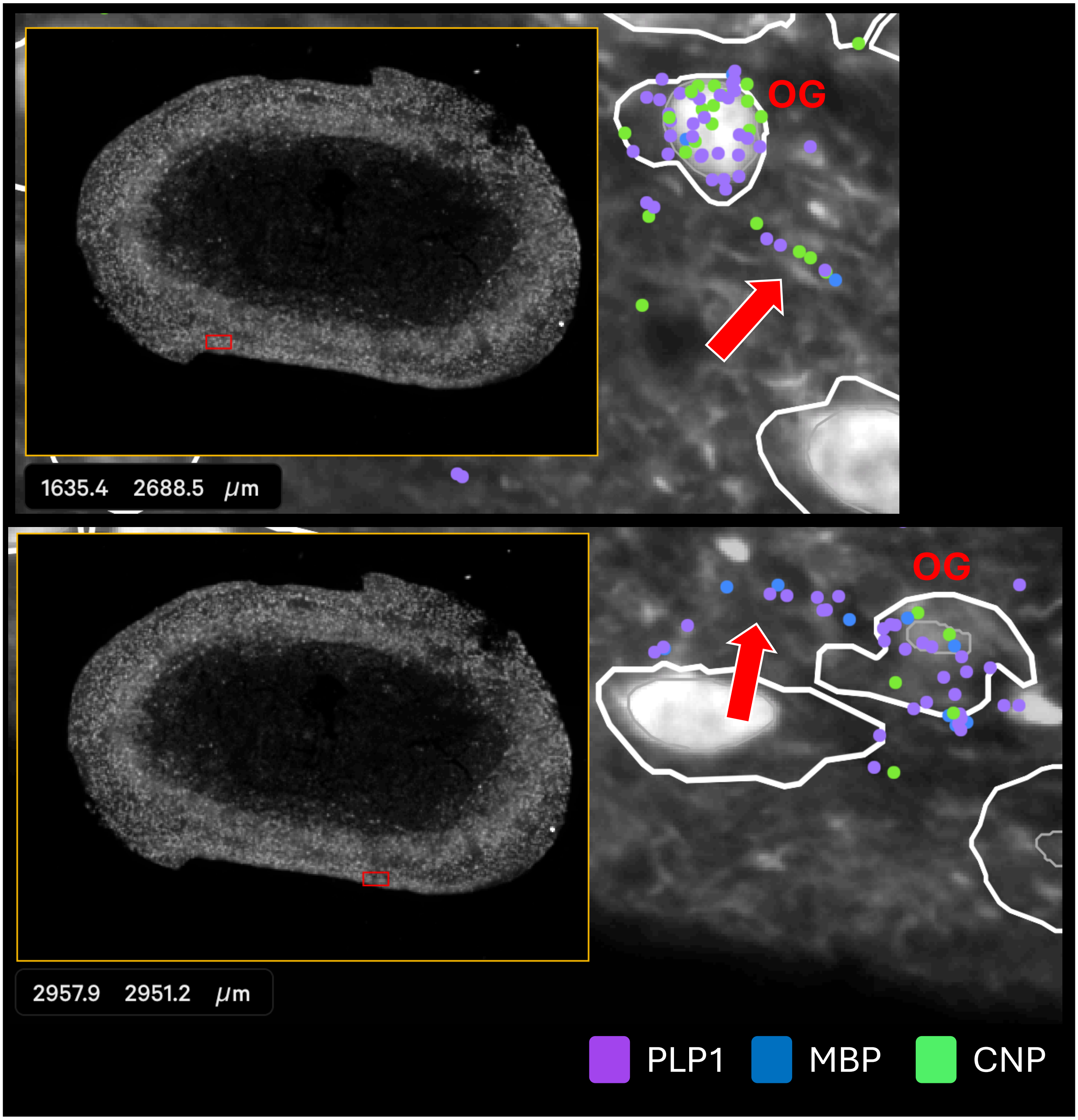


**Figure S20.** Representative Xenium spatial transcriptomic image of a day 750 cerebral organoid (donor BB026, CO alone) showing the linear assembly of myelin-associated transcripts. Myelin-associated transcripts are identified by the co-localization of *MBP, PLP1, CNP*, forming elongated extracellular myelin-like structures adjacent to oligodendrocytes.

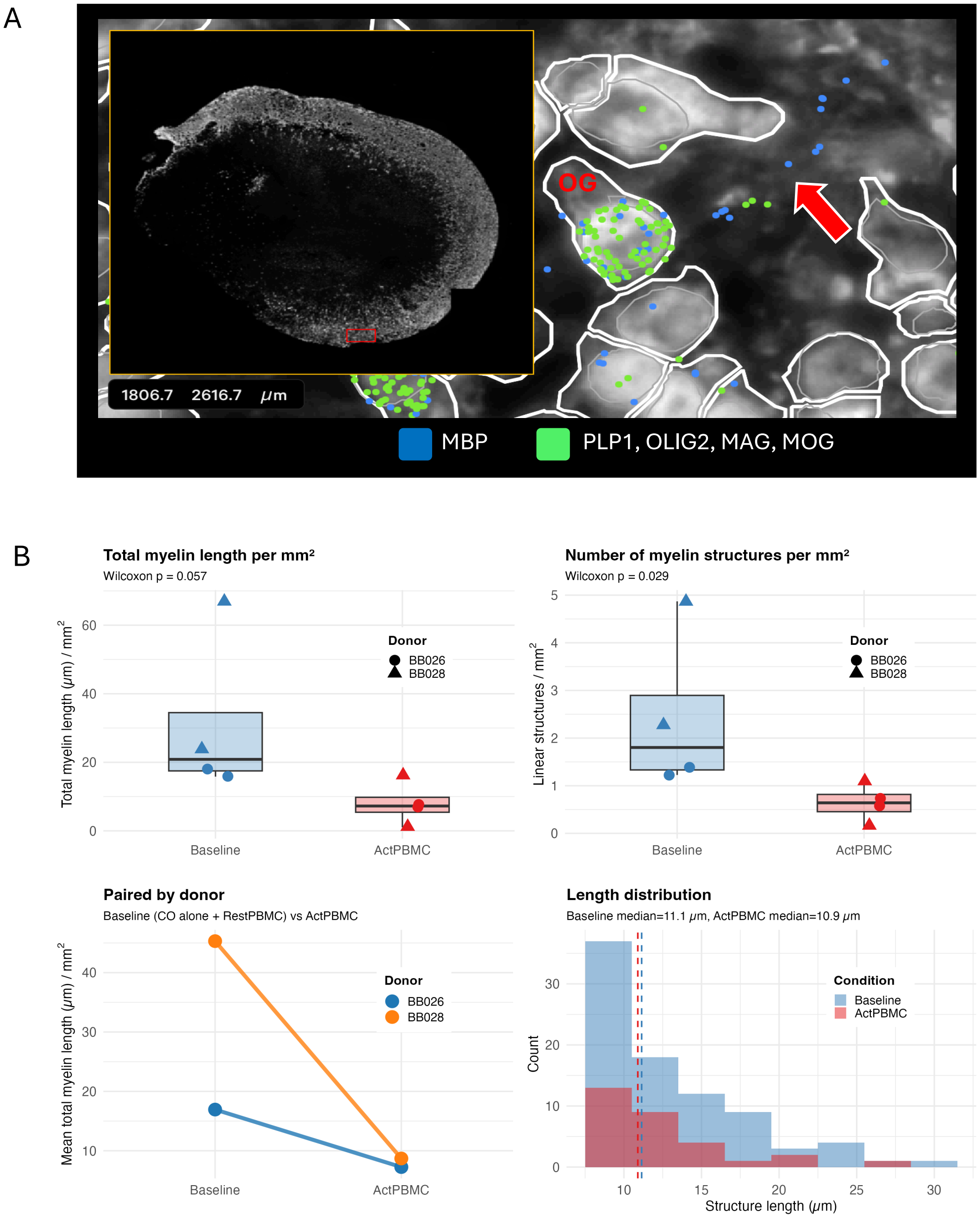


**Figure 21.** Activated PBMC reduce extracellular myelin structures in aged cerebral organoids.

A. Representative Xenium spatial transcriptomic image of a day 750 cerebral organoid showing extracellular myelin structures identified by the co-localization of *MBP* transcripts with oligodendrocyte/myelin-associated transcripts (*PLP1, OLIG2, MAG,* and *MOG*). The inset shows the analyzed organoid section, and the red box indicates the region displayed at higher magnification. The arrow indicates a representative extracellular myelin structure adjacent to an oligodendrocyte (OG).

B. Quantification of extracellular myelin structures (MBP+PLP) in aged cerebral organoids from donors BB026 and BB028 cultured under baseline conditions (CO alone + resting PBMC, n=4) or following exposure to activated PBMCs (ActPBMC, n=4). Total extracellular myelin length per $mm^2$ (top left), number of extracellular myelin structures per $mm^2$ (top right), paired donor analysis (bottom left), and length distribution of individual extracellular myelin structures (bottom right) are shown. Statistical significance was assessed using the Wilcoxon signed-rank test.

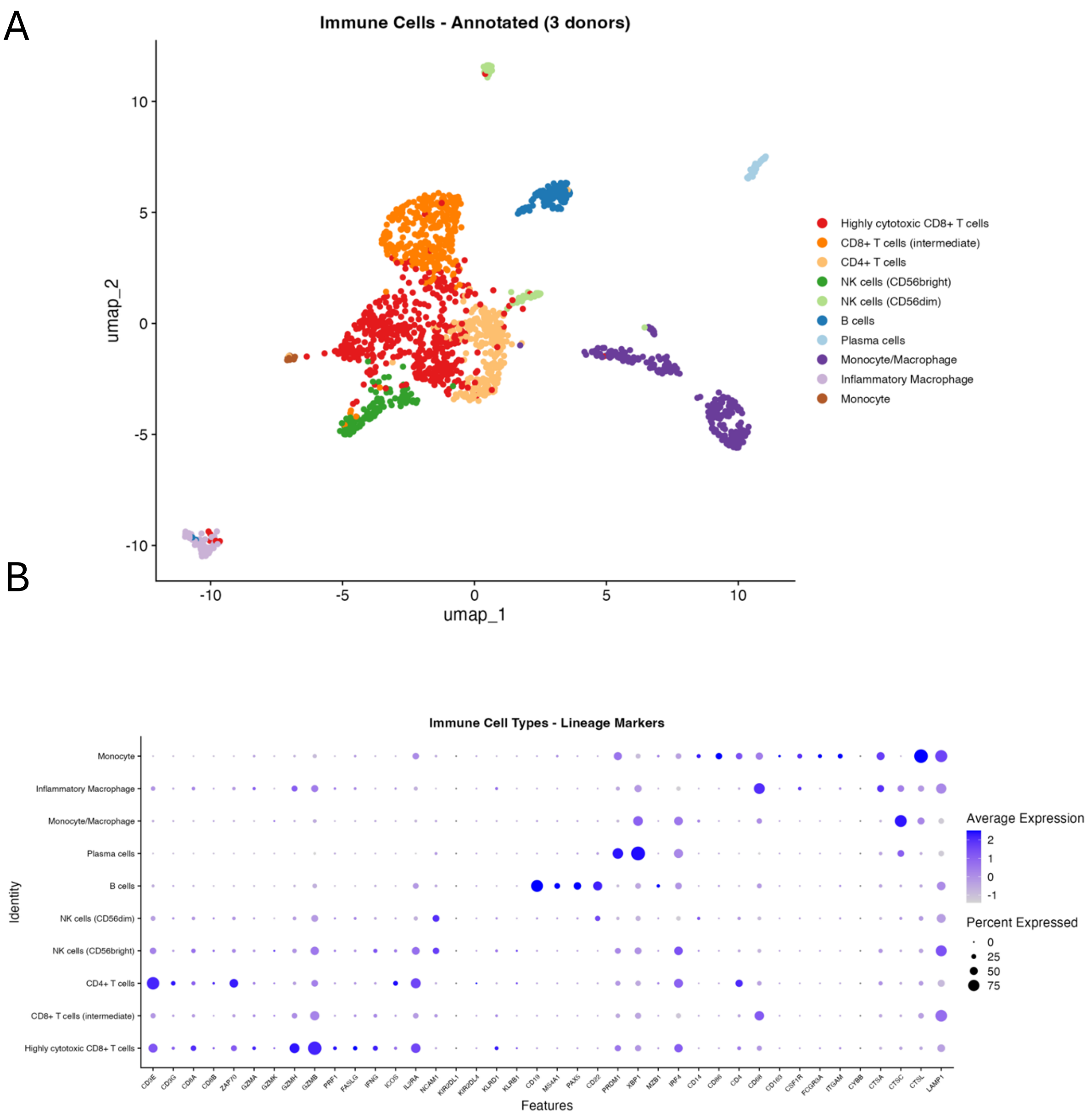


**Figure S22.** Immune cell populations incorporated into cerebral organoids following PBMC co-culture.

A. UMAP visualization of immune cells identified in cerebral organoids following PBMC co-culture, integrated across three donors (BB007, *n* = 4 organoids; BB026, *n* = 2 organoids; and BB028, *n* = 2 organoids). Cells are colored according to the major immune cell populations, including cytotoxic and helper T cells, NK cells, B cells, plasma cells, monocytes, and macrophage subsets. Cell identities were assigned based on canonical marker gene expression as described in the Methods**.**
B. Dot plot showing the expression of representative canonical marker genes used for immune cell annotation. Dot size represents the percentage of cells expressing each gene, and color intensity indicates the average normalized expression level.

**Table S1.** Summary of performed experiments

| | Multiplexed immunofluorescence staining | | | | Fusion with vessel organoid | Immunofluorescence staining of whole organoid (DCX, Iba and TMEM119) | Oligodendrocytes' induction | Co-culture with microglia progenitors | Immunofluorescence staining for Iba and TMEM119 | Xenium | Live staining | Xenium |
|---|---|---|---|---|---|---|---|---|---|---|---|---|
| **Day:** | **d. 40** | **d.98** | **d.140** | **d.540** | **d.8** | **d.45** | **d. 50-70** | **d.30** | **d. 92 and 180** | **d.200** | **d.240** | **d. 750** |
| **Experiment:** | | | | co-cultured with PBMC, 24h and 72h | | | | | co-cultured with microglia progenitors at d. 30 | | co-cultured with PBMC for 24h | co-cultured with PBMC for 24h |
| **Batch 1 bb026** | | v | v | | | | v | v | | | | v |
| **Batch 1 bb028** | | | v | | | | v | v | | | | v |
| **Batch 2 bb011** | v | | | v | | | v | v | v | | | |
| **Batch 2 bb026** | v | | | v | | | v | v | v | | v | |
| **Batch 2 bb028** | v | | | v | | | v | v | v | | v | |
| **Batch 3 bb007** | | | | | v | v | v | | | v | | |
